# Geographic and age-related variations in mutational processes in colorectal cancer

**DOI:** 10.1101/2025.02.13.25322219

**Authors:** Marcos Díaz-Gay, Wellington dos Santos, Sarah Moody, Mariya Kazachkova, Ammal Abbasi, Christopher D Steele, Raviteja Vangara, Sergey Senkin, Jingwei Wang, Stephen Fitzgerald, Erik N Bergstrom, Azhar Khandekar, Burçak Otlu, Behnoush Abedi-Ardekani, Ana Carolina de Carvalho, Thomas Cattiaux, Ricardo Cortez Cardoso Penha, Valérie Gaborieau, Priscilia Chopard, Christine Carreira, Saamin Cheema, Calli Latimer, Jon W Teague, Anush Mukeriya, David Zaridze, Riley Cox, Monique Albert, Larry Phouthavongsy, Steven Gallinger, Reza Malekzadeh, Ahmadreza Niavarani, Marko Miladinov, Katarina Erić, Sasa Milosavljevic, Suleeporn Sangrajrang, Maria Paula Curado, Samuel Aguiar, Rui Manuel Reis, Monise Tadin Reis, Luis Gustavo Romagnolo, Denise Peixoto Guimarães, Ivana Holcatova, Jaroslav Kalvach, Carlos Alberto Vaccaro, Tamara Alejandra Piñero, Beata Świątkowska, Jolanta Lissowska, Katarzyna Roszkowska-Purska, Antonio Huertas-Salgado, Tatsuhiro Shibata, Satoshi Shiba, Surasak Sangkhathat, Taned Chitapanarux, Gholamreza Roshandel, Patricia Ashton-Prolla, Daniel C Damin, Francine Hehn de Oliveira, Laura Humphreys, Trevor D. Lawley, Sandra Perdomo, Michael R Stratton, Paul Brennan, Ludmil B Alexandrov

**Affiliations:** Department of Cellular and Molecular Medicine, University of California San Diego, La Jolla, CA, USA; Department of Bioengineering, University of California San Diego, La Jolla, CA, USA; Moores Cancer Center, University of California San Diego, La Jolla, CA, USA; Digital Genomics Group, Structural Biology Program, Spanish National Cancer Research Center (CNIO), Madrid, Spain; Genomic Epidemiology Branch, International Agency for Research on Cancer (IARC/WHO), Lyon, France; Cancer, Ageing and Somatic Mutation, Wellcome Sanger Institute, Cambridge, UK; Biomedical Sciences Graduate Program, University of California San Diego, La Jolla, CA, USA; Division of Cancer Epidemiology and Genetics, National Cancer Institute, Bethesda, MD, USA; Department of Health Informatics, Graduate School of Informatics, Middle East Technical University, Ankara, Turkey; Evidence Synthesis and Classification Branch, International Agency for Research on Cancer (IARC/WHO), Lyon, France; Clinical Epidemiology, N.N. Blokhin National Medical Research Centre of Oncology, Moscow, Russia; Ontario Tumour Bank, Ontario Institute for Cancer Research, Toronto, ON, Canada; Centre for Biodiversity Genomics, University of Guelph, Guelph, ON, Canada; Lunenfeld-Tanenbaum Research Institute, Sinai Health System, Toronto, ON, Canada; Digestive Oncology Research Center, Digestive Disease Research Institute, Tehran University of Medical Sciences, Tehran, Iran; Clinic for Digestive Surgery - First Surgical Clinic, University Clinical Centre of Serbia, Belgrade, Serbia; Department of Pathology, University Clinical Centre of Serbia, Belgrade, Serbia; International Organization for Cancer Prevention and Research, Belgrade, Serbia; National Cancer Institute, Bangkok, Thailand; Department of Epidemiology, A.C. Camargo Cancer Center, Sao Paulo, Brazil; Colon Cancer Reference Center, A.C. Camargo Cancer Center, Sao Paulo, Brazil; Molecular Oncology Research Center, Barretos Cancer Hospital, Barretos, Brazil; Life and Health Sciences Research Institute (ICVS), School of Medicine, Minho University, Braga, Portugal; Department of Pathology, Barretos Cancer Hospital, Barretos, Brazil; Department of Colorectal Oncology Surgery, Barretos Cancer Hospital, Barretos, Brazil; Department of Endoscopy, Barretos Cancer Hospital, Barretos, Brazil; Institute of Public Health & Preventive Medicine, 2nd Faculty of Medicine, Charles University, Prague, Czech Republic; Department of Oncology, 2nd Faculty of Medicine, Charles University and Motol University Hospital, Prague, Czech Republic; Surgery Department, 2nd Faculty of Medicine, Charles University and Central Military Hospital, Prague, Czech Republic; 2nd Faculty of Medicine, Charles University and Motol University Hospital, Prague, Czech Republic; Institute of Animal Physiology and Genetics Czech Academy of Science, Libechov, Czech Republic; Clinical Center ISCARE, Prague, Czech Republic; Instituto de Medicina Traslacional e Ingeniería Biomédica (IMTIB)-CONICET-Universidad Hospital Italiano de Buenos Aires (UHIBA) y Hospital Italiano de Buenos Aires (HIBA), Buenos Aires, Argentina; Department of Environmental Epidemiology, Nofer Institute of Occupational Medicine, Łódź, Poland; The Maria Sklodowska-Cure National Research Institute of Oncology, Warsaw, Poland; Department of Pathology, The Maria Sklodowska-Cure National Research Institute of Oncology, Warsaw, Poland; Oncological pathology group, Terry Fox National Tumor Bank (Banco Nacional de Tumores Terry Fox), National Cancer Institute, Bogotá, Colombia; Laboratory of Molecular Medicine, The Institute of Medical Science, The University of Tokyo, Minato-ku, Japan; Division of Cancer Genomics, National Cancer Center Research Institute, Chuo-ku, Japan; Translational Medicine Research Center, Faculty of Medicine, Prince of Songkla University, Hat Yai, Thailand; Department of Biomedical Sciences and Biomedical Engineering, Faculty of Medicine, Prince of Songkla University, Hat Yai, Thailand; Department of Surgery, Faculty of Medicine, Prince of Songkla University, Hat Yai, Thailand; Department of Internal Medicine, Faculty of Medicine, Chiang Mai University, Chiang Mai, Thailand; Golestan Research Center of Gastroenterology and Hepatology, Golestan University of Medical Sciences, Gorgan, Iran; Department of Genetics, Universidade Federal do Rio Grande do Sul (UFRGS), Porto Alegre, Brazil; 46Medical Genetics Service, Hospital de Clínicas de Porto Alegre (HCPA), Porto Alegre, Rio Grande do Sul, Brazil; Department of Surgery, Division of Colorectal Surgery, Hospital de Clínicas de Porto Alegre (HCPA), Porto Alegre, Rio Grande do Sul, Brazil; Department of Pathology, Anatomic Pathology, Hospital de Clínicas de Porto Alegre (HCPA), Porto Alegre, Rio Grande do Sul, Brazil; Parasites and Microbes, Wellcome Sanger Institute, Cambridge, UK; Sanford Stem Cell Institute, University of California San Diego, La Jolla, CA, USA

## Abstract

Colorectal cancer incidence rates vary geographically and have changed over time. Notably, in the past two decades, the incidence of early-onset colorectal cancer, affecting individuals under the age of 50 years, has doubled in many countries. The reasons for this increase are unknown. Here, we investigate whether mutational processes contribute to geographic and age-related differences by examining 981 colorectal cancer genomes from 11 countries. No major differences were found in microsatellite unstable cancers, but variations in mutation burden and signatures were observed in the 802 microsatellite-stable cases. Multiple signatures, most with unknown etiologies, exhibited varying prevalence in Argentina, Brazil, Colombia, Russia, and Thailand, indicating geographically diverse levels of mutagenic exposure. Signatures SBS88 and ID18, caused by the bacteria-produced mutagen colibactin, had higher mutation loads in countries with higher colorectal cancer incidence rates. SBS88 and ID18 were also enriched in early-onset colorectal cancers, being 3.3 times more common in individuals diagnosed before age 40 than in those over 70, and were imprinted early during colorectal cancer development. Colibactin exposure was further linked to *APC* driver mutations, with ID18 responsible for about 25% of *APC* driver indels in colibactin-positive cases. This study reveals geographic and age-related variations in colorectal cancer mutational processes, and suggests that early-life mutagenic exposure to colibactin-producing bacteria may contribute to the rising incidence of early-onset colorectal cancer.

## INTRODUCTION

The age-standardized incidence rates (ASR) for most adult cancers vary across different geographic locations and can change over time^1^. Despite extensive epidemiological research, the underlying causes for many of these variations remain unclear. However, they are suspected to be due to exogenous environmental or lifestyle carcinogenic exposures, which are, in principle, preventable^2^. Many well-known exogenous carcinogens are also mutagens^3,4^, which can imprint characteristic patterns of somatic mutations in the genome, known as mutational signatures. Therefore, a complementary approach to conventional epidemiology for investigating unknown causes of cancer is the characterization of mutational signatures in the genomes of cancer and normal cells^5–7^. The *Mutographs* Cancer Grand Challenge project^8^ has implemented this strategy of “mutational epidemiology” by sequencing cancers from geographic areas of differing incidence rates, using mutational signature analysis to elucidate the mutational processes that have been operative, with results thus far from cancers of the esophagus^6^, kidney^5^, and head and neck^9^.

Colorectal cancer incidence rates differ markedly by geographic location and have changed substantially in some countries over the last 70 years^10^. For instance, the ASR for colorectal cancer in North America and in most European countries peaked in the 1980s and 1990s and have been declining since, whereas countries in East Asia such as Japan and South Korea have been steadily increasing over the past seven decades^1^. Moreover, in the past 20 years there has been a notable global increase in the incidence of early-onset colorectal cancer^10,11^, typically defined as colorectal cancer in adults under 50 years of age. This was first reported in the United States^12^ and subsequently observed in Australia, Canada, Japan^13^ and multiple European countries^14^. Although epidemiological studies have identified multiple risk factors for colorectal cancer, specific risk factors for early-onset colorectal cancer remain largely unidentified, with the exception of family history and hereditary predisposition. The latter is predominantly attributable to Lynch syndrome, which is characterized by DNA mismatch repair deficient cancers of the proximal colon^15,16^ and, therefore, is unlikely to be implicated in the recent increase in early-onset colorectal cancer, which is mainly enriched in sporadic, DNA mismatch repair proficient cancers affecting the distal colon and rectum^17,18^.

Previous colorectal cancer whole-genome sequencing studies have largely focused on cases from North America and Europe including USA^19,20^, UK^21,22^, Netherlands^23–26^, and Sweden^27^ and incorporated limited numbers of early-onset cases^19,21,22,26,27^. Here, we examine colorectal cancer genomes from 11 countries on four continents to investigate whether variation in mutational processes contributes to geographic and age-related differences in incidence rates.

## RESULTS

### Study design

981 colorectal cancers (45.7% female) were collected from intermediate-incidence countries with ASRs of 13-20/100,000 people (Iran, Thailand, Colombia, Brazil) and high-incidence countries with ASRs >24 (Argentina, Canada, Russia, Serbia, Czech Republic, Poland, Japan), including the highest ASR of 37 in Japan^1^ (**Fig. 1*a***; **Supplementary Table 1**). Of the 981 cases, 320 were from the proximal colon, 333 from the distal colon, 326 from the rectum, and 2 from unspecified subsites (**Fig. 1*b***). There were 132 early-onset cases, which were 1.88-fold enriched in the distal colon and rectum compared to the proximal colon (*p*=0.006). All cancers and their matched normal samples underwent whole genome sequencing, achieving a median coverage of 53-fold and 27-fold, respectively.

**Fig. 1.**
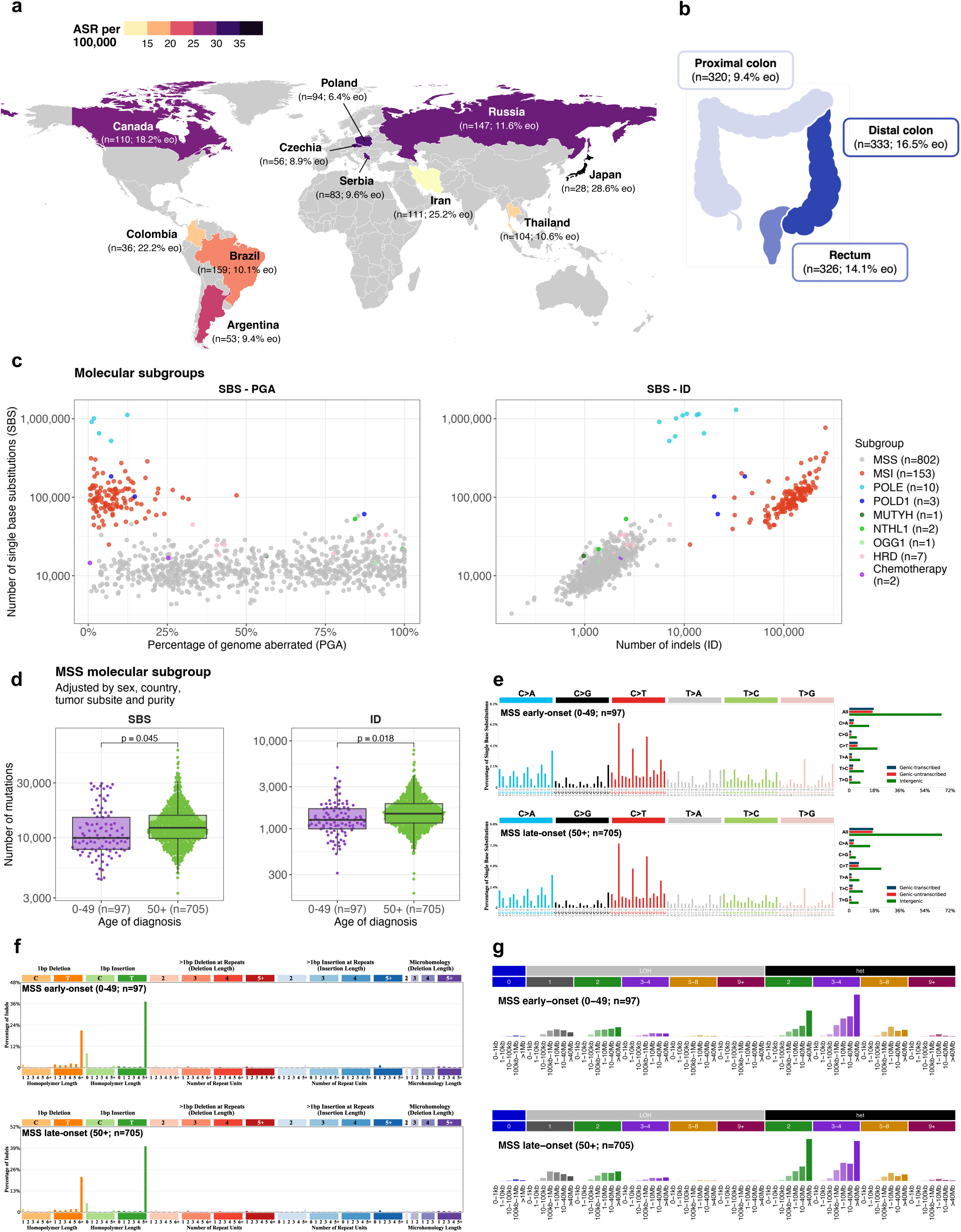
Geographic, clinical, and molecular characterization of the Mutographs colorectal cancer cohort. **a**, Geographic distribution of the 981 patients across four continents and 11 countries, with an indication of the total number of cases as well as the percentage of early-onset (eo) cases below 50 years of age. Countries were colored according to their age-standardized incidence rates (ASR) per 100,000 individuals. The designations employed and the presentation of the material in this publication do not imply the expression of any opinion whatsoever on the part of the authors or their institutions concerning the legal status of any country, territory, city or area or of its authorities, or concerning the delimitation of its frontiers or boundaries. **b**, Tumor subsite distribution of the cohort across the colorectum, with an indication of the total number of cases and the percentage of early-onset cases. Different subsites were colored according to the percentage of early-onset cases. Additional 2 cases had unspecified subsites. **c**, Scatter plots indicating the distribution of molecular subgroups across the sequenced tumors according to the total number of single base substitutions (SBS) and small insertions and deletions (indels; ID), as well as the percentage of genome aberrated (PGA). Cases for which tumor purity was insufficient to determine an accurate copy number profile or without large copy number alterations (65/981) were excluded from the SBS - PGA panel. **d**, Box plots indicating the distribution of SBS and ID across early-onset (under 50 years; purple) and late-onset (50 years or older; green) microsatellite stable (MSS) colorectal tumors. Statistically significant differences were evaluated using multivariable linear regression models adjusted by sex, country, tumor subsite, and tumor purity. The line within the box is plotted at the median, while the upper and lower ends indicate the 25^th^ and 75^th^ percentiles. Whiskers show 1.5 × interquartile range, and values outside it are shown as individual data points. **e-g**, Average mutational profiles of early-onset and late-onset MSS colorectal tumors for SBS (SBS-288 mutational context; **e**), ID (ID-83 mutational context; **f**), and copy number alterations (CN-68 mutational context; **g**).

### Mutation burden and molecular classification

The 981 colorectal cancers were divided into known molecular subtypes based on their somatic mutation burdens and profiles. Consistent with prior studies^20,28^, two main subtypes were identified: DNA mismatch repair proficient cancers, also known as microsatellite stable (MSS), and DNA mismatch repair deficient cancers, often referred to as tumors showing microsatellite instability (MSI). MSS samples (*n*=802, 81.8%; **Fig. 1*c***) were characterized by a lower burden of single base substitutions (SBS; median: 12,054) and small insertions and deletions (ID; median: 1,451), and a higher burden of large-scale genomic aberrations (median: 53.5% of genome altered). In contrast, MSI samples (*n*=153, 15.6%) exhibited higher SBS and ID burdens (median: 95,426 and 125,100, respectively) with limited genomic aberrations (median: 7.0%). As expected, the average mutational profiles of MSS and MSI colorectal tumors were different (**Extended Data Fig. 1a-b**).

MSI samples were predominantly found in the proximal colon (OR=12.2, *p*=3.8×10^-27^) and were more common in early-onset cases (OR=2.6, *p*=0.001). Notably, 31/153 MSI cases (20.3%), including 13/28 MSI early-onset cases (46.4%), carried germline pathogenic variants in DNA mismatch repair genes consistent with Lynch syndrome (**Supplementary Table 2**). After excluding all cases attributed to Lynch syndrome, there was no enrichment of MSI cancers in early-onset cases (*p*>0.05). Deficiencies of other DNA repair mechanisms were observed in 24/981 cancers (2.4%), including ultra-hypermutated cases with mutations in *POLE* (*n*=10, 1.0%) and *POLD1* polymerases (*n*=3, 0.3%), homologous recombination deficient (HRD) cases (*n*=7, 0.7%), and cases with mutations in the base excision repair genes *MUTYH* (*n*=1, 0.1%), *NTHL1* (*n*=2, 0.2%), and *OGG1* (*n*=1, 0.1%) (**Methods**; **Supplementary Table 3-4**; **Supplementary Fig. 1-3**).

The mutational catalogues of DNA repair deficient cancers are dominated by somatic mutations resulting from the failed repair process, rendering it difficult to characterize mutational processes unrelated to this failure^29^. To enable investigation of the latter, we therefore focused the main analyses on DNA repair proficient colorectal cancers, while reporting DNA repair deficient cases in the **Supplementary Note**. Two cases treated with chemotherapy for prior cancers were also excluded as their mutation profiles were dominated by the mutational signatures of chemotherapy agents^19,30^ (**Supplementary Fig. 4**). The remaining cohort consisted of 802 treatment-naïve DNA repair proficient colorectal cancers, including 97 early-onset cases.

After adjustment for sex, country, tumor subsite, and tumor purity (**Methods**), early-onset cancers showed reduced burdens of SBS (fold-change [FC]=0.92, *p*=0.045) and ID (FC=0.90, *p*=0.018; **Fig. 1*d***) but not of doublet base substitutions (DBS), copy number alterations (CN), or structural variants (SV) when compared to late-onset cases (*p*>0.05). Nevertheless, the average mutation spectra of early-onset and late-onset cancers were remarkably similar for all types of somatic mutations (cosine similarity>0.97; **Fig. 1*e*-*g***; **Extended Data Fig. 1c-d**). Mutation burden also varied substantially for specific countries when compared to all others, including Canada (lower SBS and ID burdens), Poland (higher SBS and DBS), Japan (lower SBS, ID, DBS), Iran (lower ID), and Brazil (higher ID and CN; **Extended Data Fig. 2**). However, mutation profiles were generally consistent across all countries (**Extended Data Fig. 3**).

### Repertoire of mutational signatures

A total of 16 SBS, 10 ID, 4 DBS, 6 CN, and 6 SV *de novo* mutational signatures were extracted from the 802 MSS colorectal cancers and subsequently decomposed into a combination of previously reported reference signatures and potential novel signatures (**Supplementary Note**; **Supplementary Tables 5-15**). The 16 *de novo* SBS signatures encompassed 15 COSMICv3.4 signatures (**Extended Data Fig. 4a**; **Supplementary Table 10**), including those previously associated with clock-like mutational processes (SBS1, SBS5)^31^, APOBEC deamination (SBS2, SBS13)^31^, deficient homologous recombination (SBS3)^31^, reactive oxygen species (SBS18)^32^, exposure to the mutagenic agent colibactin synthesized by *Escherichia coli* and other microbes carrying a ∼40kb polyketide synthase (*pks*) pathogenicity island (SBS88)^33,34^, and mutational processes of unknown causes (SBS8, SBS17a/b, SBS34, SBS40a, SBS89, SBS93, SBS94)^5,19,32,34,35^. Three previously described signatures of unknown origin^21^ (SBS_F, SBS_H, SBS_M; **Extended Data Fig. 4b**) and a novel signature (SBS_O; **Extended Data Fig. 4c**) were also detected. SBS_O corresponds to a refined version of a previously reported signature of unknown etiology (SBS41; **Methods**)^19^. With respect to ID, DBS, CN, and SV, most *de novo* extracted mutational signatures were highly similar to, or directly reconstructed by, COSMICv3.4 reference signatures (**Extended Data Fig. 4d-f** and **5**; **Supplementary Table 10**) with the exception of an ID signature (ID_J), characterized by deletions of isolated Ts and insertions of Ts in long repetitive regions resembling a previously reported signature^34^ (**Extended Data Fig. 4e**), and three novel signatures from large mutational events (CN_F, SV_B, SV_D; **Extended Data Fig. 5b*&d***), which were extracted due to the extended contexts used in our signature analysis (**Methods**).

### Geographic variation in mutational signatures

Despite the similar mutation profiles across countries (**Extended Data Fig. 3**), several signatures exhibited varying prevalence when comparing one country to all others (**Fig. 2*a***; **Supplementary Fig. 5**; **Supplementary Table 16**). Notably, SBS89 (OR=28.0, *q*=0.001), DBS8 (OR=8.9, *q*=3.2×10^-4^), and the novel ID_J (OR=9.6, *q*=6.2×10^-5^) were at elevated frequencies in Argentina when compared to all other countries (**Fig. 2*b***). Signatures SBS89, DBS8, and ID_J also showed a strong tendency to co-occur (*p*<1.7×10^-11^) suggesting they may arise from the same underlying mutational process. In Colombia (**Fig. 2*c***), higher frequencies were observed for SBS94 (OR=19.7, *q*=3.2×10^-5^), the novel SBS_F (OR=10.7, *q*=2.0×10^-4^), and DBS6 (OR=12.5, *q*=0.028) when compared to all other countries, with evidence of co-occurrence of SBS94 with SBS_F (*p*=0.017) and DBS6 (*p*=1.9×10^-4^). Enrichments were also found for SBS2 (OR=2.0, *q*=0.041) and SBS_H (OR=2.3, *q*=0.001) in Russia and CN_F (OR=3.5, *q*=3.9×10^-4^) in Brazil, whereas depletions were identified for DBS2 in Thailand (OR=0.38, *q*=0.008) and for DBS4 in Colombia (OR=0.06, *q*=0.034; **Fig. 2*a***). Overall, the results indicate international differences in the prevalence of certain mutational processes involved in colorectal cancer development.

**Fig. 2.**
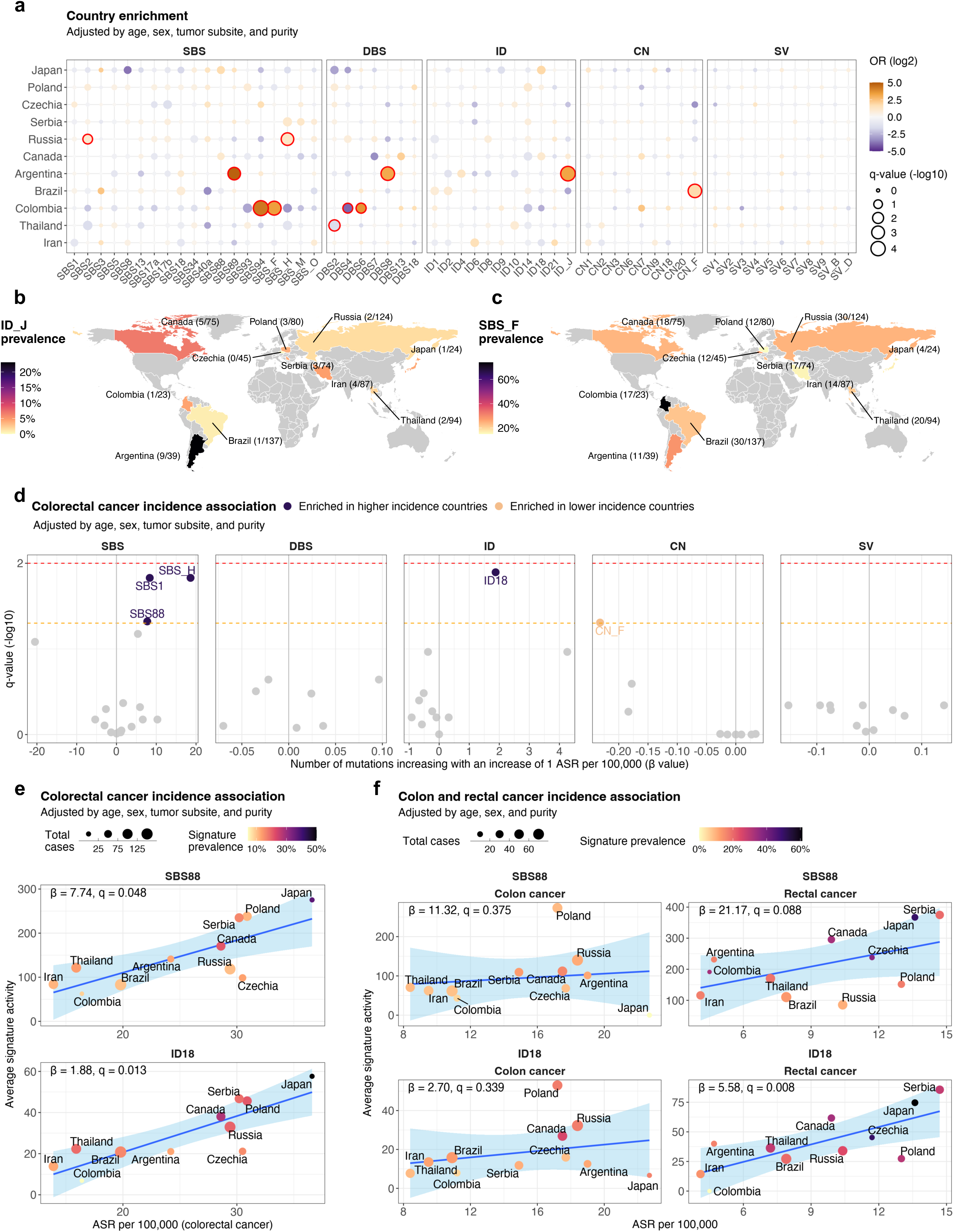
Geographic variation of mutational signatures in microsatellite stable colorectal cancers. **a**, Dot plot indicating the variation of mutational signature prevalence in specific countries compared to all others. Statistically significant enrichments were evaluated using multivariable logistic regression models adjusted by age of diagnosis, sex, tumor subsite, and tumor purity. Firth’s bias-reduced logistic regressions were used for regression presenting complete or quasi-complete separation. Data points were colored according to the odds ratio (OR) of the enrichment, with their size representing statistical significance. P-values were adjusted for multiple comparisons based on the total number of mutational signatures considered per variant type and the total number of countries assessed, and reported as q-values. Q-values<0.05 were considered statistically significant and marked in red. **b-c**, Geographic distribution of the ID_J (**b**) and SBS_F (**c**) mutational signatures. Countries were colored based on the signature prevalence. **d**, Volcano plots indicating the association of mutational signature activities with the age-standardized incidence rates. Statistically significant associations were evaluated using multivariable linear regression models adjusted by age of diagnosis, sex, tumor subsite, and tumor purity. P-values were adjusted for multiple comparisons based on the total number of mutational signatures considered per variant type and reported as q-values. Horizontal lines marking statistically significant thresholds were included at 0.05 (dashed orange line) and 0.01 q-values (dashed red line). **e-f**, Scatter plots indicating the association of the mutations attributed to the SBS88 and ID18 mutational signatures with the age-standardized incidence rates (ASR) across countries for colorectal cancer (**e**), and independently for colon and rectal cancers (**f**). Data points were colored based on signature prevalence, with their size indicating the total number of cases per country. Statistically significant associations were evaluated using the sample-level multivariable linear regression models used in **d** (**e**), and similar multivariable linear regression models adjusted by age of diagnosis, sex, and tumor purity (**f**).

To explore the broader epidemiological implications of international variation in mutational processes, as previously done for kidney cancer^5^, we evaluated the relationships between ASR and mutational signatures (**Fig. 2*d***; **Supplementary Table 17**). Independent of covariates, colibactin-induced mutational signatures, SBS88 and ID18, as well as clock-like signature SBS1 and novel signature SBS_H, associated with an increasing rate of ASR for colorectal cancer, whereas novel signature CN_F associated with a reduced ASR rate (*q*<0.05; **Fig. 2*d-e***; **Extended Data Fig. 6a**). For SBS88 and ID18, the association was linked with the ASR for rectal cancer (*q*=0.088 and *q*=0.008; **Fig. 2*f***; **Supplementary Table 18**). In contrast, for SBS1, SBS_H, and CN_F the association was particularly strong for the ASR of colon cancer (*q*=0.009, *q*=0.015, and *q*=0.057; **Extended Data Fig. 6b**). Colibactin-associated signatures were also found elevated in patients from countries with high ASR rates for early-onset colorectal cancer (**Extended Data Fig. 6c**).

### Colibactin induced mutational signatures are enriched in early-onset colorectal cancer

In addition to examining the global distribution of mutational signatures, the substantial number of early-onset colorectal cancer cases enabled evaluating the association between mutational signatures and age at diagnosis. Although the average mutation profiles of early-onset and late-onset colorectal cancer cases were similar (**Fig. 1*e-g***), the prevalence of some mutational signatures was associated with the age of diagnosis, independently of country of origin (**Fig. 3*a***; **Supplementary Table 19**), genetic ancestry or ethnicity (**Supplementary Fig. 6-8**). As expected, late-onset cases showed enrichment in signatures known to accumulate linearly with age in normal colorectal crypts^36^, including SBS1, SBS5, ID1, and ID2 (**Fig. 3*a-b***). Unknown etiology indel signatures ID4, ID9, and ID10 also showed associations with late-onset cases (**Fig. 3*a***-***b***).

**Fig. 3.**
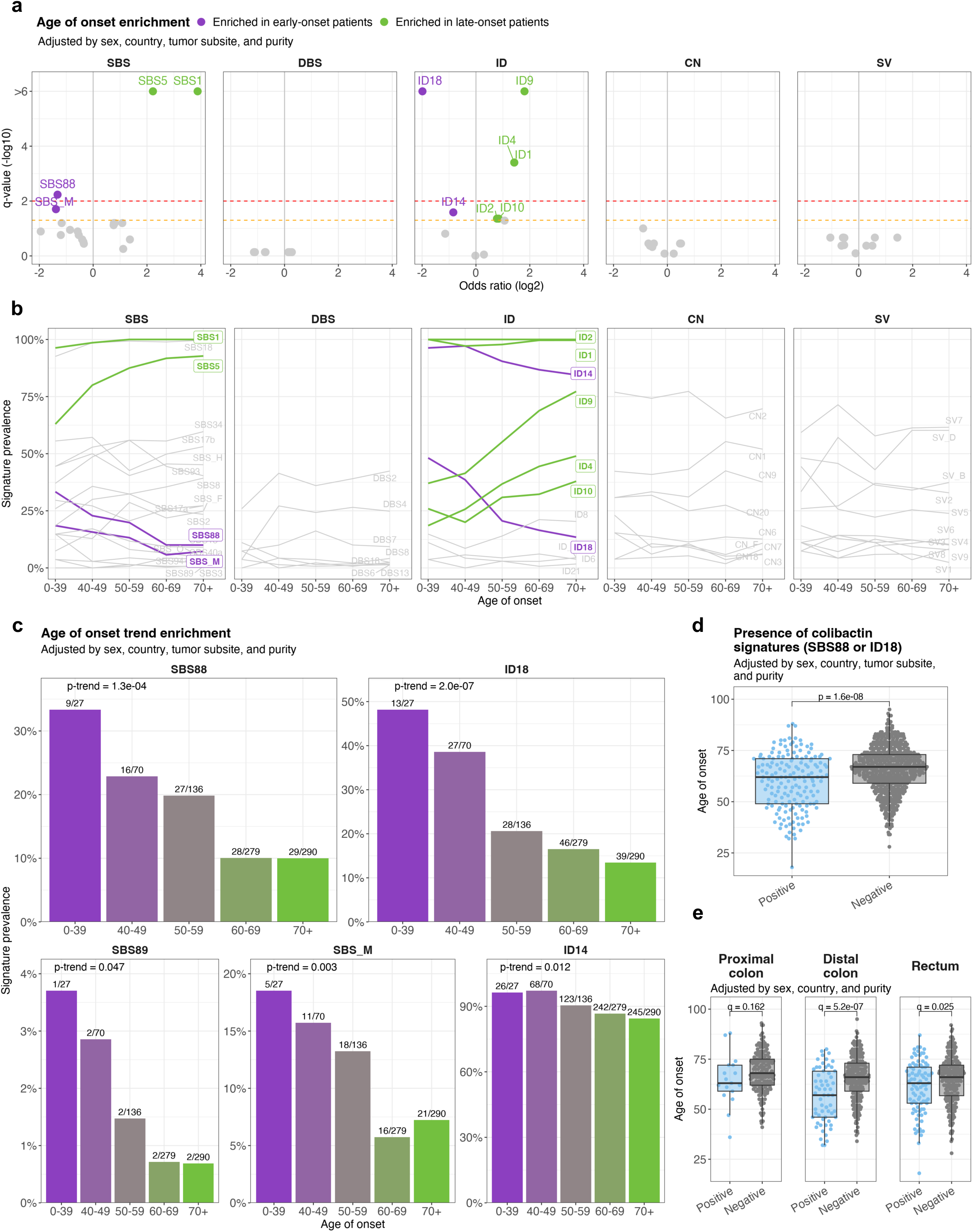
Variation of mutational signatures with age of onset in microsatellite stable colorectal cancers. **a**, Volcano plots indicating the enrichment of mutational signature prevalence in early-onset and late-onset cases. Statistically significant enrichments were evaluated using multivariable logistic regression models for age of onset categorized in two subgroups (early-onset, <50 years of age; and late-onset, ≥50) and adjusted by sex, country, tumor subsite, and tumor purity. Firth’s bias-reduced logistic regressions were used for regression presenting complete or quasi-complete separation. P-values were adjusted for multiple comparisons based on the total number of mutational signatures considered per variant type and reported as q-values. Horizontal lines marking statistically significant thresholds were included at 0.05 (dashed orange line) and 0.01 q-values (dashed red line). **b**, Line plots indicating mutational signature prevalence trend across ages of onset, using five different age groups. Signatures significantly enriched in early-onset or late-onset cases (as shown in **a**) were colored in purple and green, respective, whereas signatures not varying significantly with age were colored in grey. **c**, Bar plots indicating mutational signature prevalence across age groups, with indication of the total number of cases where signatures were detected. Statistically significant trends were evaluated using multivariable logistic regression models for age categorized in five subgroups (0-39, 40-49, 50-59, 60-69, ≥70) and adjusted by sex, country, tumor subsite, and tumor purity. Firth’s bias-reduced logistic regressions were used for regressions presenting complete or quasi-complete separation. **d-e**, Box plots indicating the variation in age of onset according to the presence of colibactin mutational signatures (either SBS88, ID18, or both) in all microsatellite stable cases (**d**) and across tumor subsites (**e**). Statistically significant differences were evaluated using multivariable linear regression models adjusted by sex, country, tumor purity, and tumor subsite (only for the analysis of all cases, **d**). The line within the box is plotted at the median, while the upper and lower ends indicate the 25^th^ and 75^th^ percentiles. Whiskers show 1.5 × interquartile range, and values outside it are shown as individual data points.

By contrast, enrichment in early-onset cancers was observed for colibactin-induced signatures. Signatures SBS88 and ID18 were 2.5 and 4 times more common, respectively, in colorectal cancers diagnosed below than above the age of 50 (*q*=0.006 and *q*=3.7×10^-7^, respectively; **Fig. 3*a-b***). The primary associations of early-onset cases with SBS88 and ID18 were further supported by the successive decline in the prevalence of these signatures with increasing age of diagnosis (*p-trend*=1.3×10^-4^ and *p-trend*=2.0×10^-7^, respectively; **Fig. 3*c***; **Supplementary Table 20)**. A similar effect was observed using a complementary motif enrichment analysis for detecting SBS88, similarly to a recent study^26^ (*p-trend*=1.0×10^-7^; **Extended Data Fig. 7a-b**). On the basis of the strong co-occurrence of SBS88 and ID18 (*p*=7.4×10^-63^), as well as previous functional^33^ and population studies^22,26,34^, we defined exposure to colibactin by the presence of either SBS88 or ID18. Colibactin exposure was found in 21.1% of all colorectal cancers (169/802) and was associated with earlier age of onset (median age: 62 vs. 67, *p*=1.6×10^-8^; **Fig. 3*d***), an effect more evident in the distal colon (median age: 57 vs. 66, *q*=5.2×10^-7^) and rectum (median age: 63 vs. 66, *q*=0.025; **Fig. 3*e***). Overall, colibactin exposure had a strong inverse correlation with age, being 3.3 times more common in colorectal cancers diagnosed in individuals younger than 40 compared to those over 70 (*p-trend*=2.7×10^-7^, **Extended Data Figure 7c**).

Signatures of unknown etiology SBS_M and ID14 (**Fig. 3*a-c***) were also enriched in early-onset cases, and SBS89 similarly exhibited a higher prevalence in younger individuals (5.8 times more prevalent in early-onset compared to late-onset patients with *p-trend*=0.047), albeit based on a very small number of cancers harboring the signature (9/802, 1.1%; **Fig. 3*c***). Interestingly, SBS_M showed an elevation in distal colon and rectum tumors compared to proximal colon similar to the one observed in colibactin-associated signatures SBS88 and ID18, previously reported^22^ (**Supplementary Fig. 9**).

### Colibactin mutagenesis is an early event in colorectal carcinogenesis

To time the imprinting of SBS88 and ID18, mutations were categorized as early clonal, late clonal, or subclonal during the development of each cancer and the contribution of each mutational signature to each category was determined (**Methods**). SBS88 and ID18 were both enriched in early clonal compared to late clonal mutations (*q*=4.2×10^-4^ and *q*=6.1×10^-5^; **Fig. 4*a***), as well as a similar trend in clonal compared to subclonal mutations (*q*=0.138 and *q*=0.058; **Extended Data Fig. 8a**), consistent with the presence of these mutational signatures in normal colorectal epithelium^34^. This enrichment in earlier evolutionary stages was similar to the one observed for other well-known clock-like signatures like SBS1, SBS5, or ID1 (**Fig. 4*a-b***), as previously shown in tumors^37,38^ and normal tissues^34^, and in contrast to signatures known to preferentially generate late clonal and subclonal mutations, such as SBS17a/b^38^. Interestingly, the enrichment of colibactin signatures in early clonal mutations was observed for both early-onset (*q*=0.004 for SBS88 and *q*=2.0×10^-4^ for ID18) and late-onset colorectal cancer cases (*q*=0.020 and *q*=0.024; **Extended Data Fig. 8b**).

**Fig. 4.**
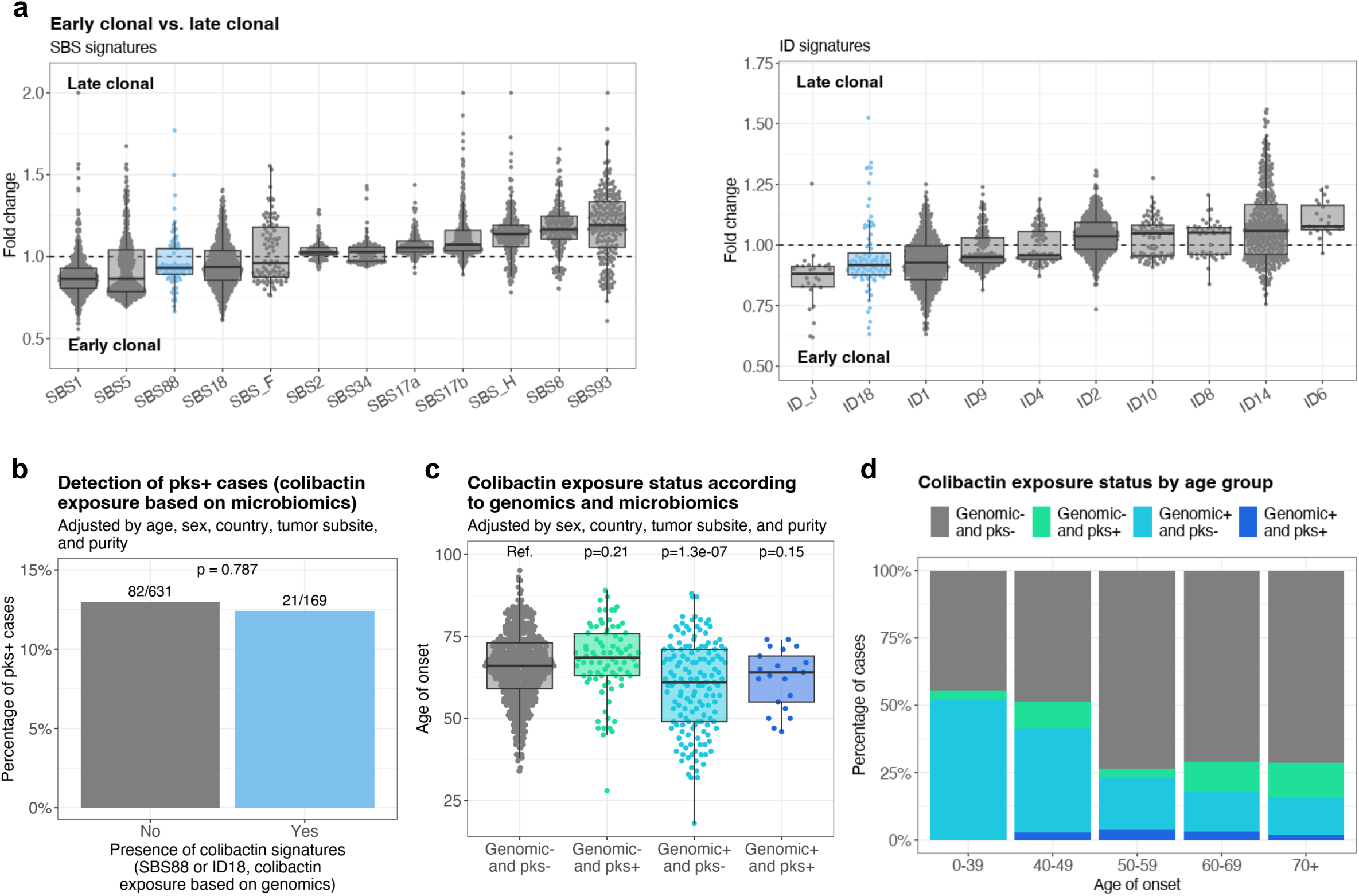
Colibactin mutagenesis as an early event in microsatellite stable colorectal cancer evolution. **a**, Box plots indicating the fold-change of the relative contribution per sample of each signature between early clonal and late clonal single base substitutions (SBS, left) and small insertions and deletions (ID, right). SBS signatures that generated early clonal SBSs in fewer than 50 samples and also generated late clonal SBSs in fewer than 50 samples were excluded from the analysis. Similarly, ID signatures that generated early clonal IDs in fewer than 20 samples and late clonal IDs in fewer than 20 samples were also excluded. Signatures were sorted by their median fold-change. The line within the box is plotted at the median, while the upper and lower ends indicate the 25^th^ and 75^th^ percentiles. Whiskers show 1.5 × interquartile range, and values outside it are shown as individual data points. **b**, Bar plot indicating the lack of concordance between colibactin exposure status determined by the presence of colibactin-induced mutational signatures SBS88 or ID18, and the microbiome *pks* status. Statistical significance was evaluated using a multivariable Firth’s bias-reduced logistic regression model (due to quasi-complete separation) adjusted by age of diagnosis, sex, country, tumor subsite, and tumor purity. **c-d**, Distribution of the age of onset (**c**) and cases across age groups (**d**) based on the detection of colibactin-positive samples using genomic and microbiome status. The genomic status is defined by the presence of SBS88 or ID18 signatures, while the microbiome status (*pks*) is determined by coverage of at least half of the *pks* island, and suggests ongoing or active *pks*+ bacterial infection. Statistical significance was evaluated using a multivariable linear regression model adjusted by sex, country, tumor subsite, and tumor purity.

Since colibactin is produced by bacteria carrying the *pks* pathogenicity island, we investigated whether colorectal cancer cases with SBS88 or ID18 harbored *pks+* bacteria based on sequencing reads from the cancer sample that did not map to the human genome but mapped to the *pks* locus (**Methods**). Consistent with a prior observation^39^, there was no association between the presence of SBS88 or ID18 and that of *pks+* bacteria (**Fig. 4*b***; **Extended Data Fig. 9**). Similarly, no microbiome association was observed for the other signatures enriched in early-onset colorectal cancers (**Supplementary Note**). Moreover, we observed a younger age of diagnosis for cases with SBS88 or ID18 but without an identified *pks+* bacteria (*p*=1.3×10^-7^; **Fig. 4*c-d***). While the reasons are unclear, one likely explanation is the imprinting of SBS88 and ID18 on the colorectal epithelium during an early period of life when *pks*+ bacteria were present, followed by the natural plasticity of the microbiome over subsequent decades, leading to the loss or gain of *pks*+ bacteria.

### Colibactin exposure and driver mutations

Using the IntOGen framework^40^, 46 genes under positive selection were identified, with eight mutated in more than 10% of cancers: *APC*, *TP53*, *KRAS*, *FBXW7*, *SMAD4*, *PIK3CA*, *TCFL2*, and *SOX9* (**Fig. 5*a***; **Supplementary Table 21**). Forty-three of the 46 genes have been previously reported as colorectal cancer driver genes^22,40^, two in other cancer types (*MED12*, *NCOR1*)^40^, and a putative novel colorectal cancer driver gene, *CCR4*, was identified with mutations indicating inactivation of the encoded protein. Mutations affecting these 46 cancer driver genes were annotated as driver mutations using a multi-step process based on the mutation type and the mode of action of the gene (**Methods**). An elevation in the total number of driver mutations was observed in late-onset compared to early-onset cases (FC=1.21, *p*=5.4×10^-5^; **Fig. 5*b***). In addition, an enrichment in *APC* driver mutation carriers was also found for late-onset cases (OR=2.7, *q*=0.027; **Fig. 5*c-d***; **Supplementary Table 22**), as previously reported^41^, whereas no hotspot driver mutation (defined as those affecting the same genomic position in at least 10 cases) was associated with age of onset (*q*>0.05; **Supplementary Table 23**). No statistically significant differences across countries were found for driver mutations within cancer driver genes or for hotspot driver mutations (*q*>0.05; **Supplementary Tables 24-25**).

**Fig. 5.**
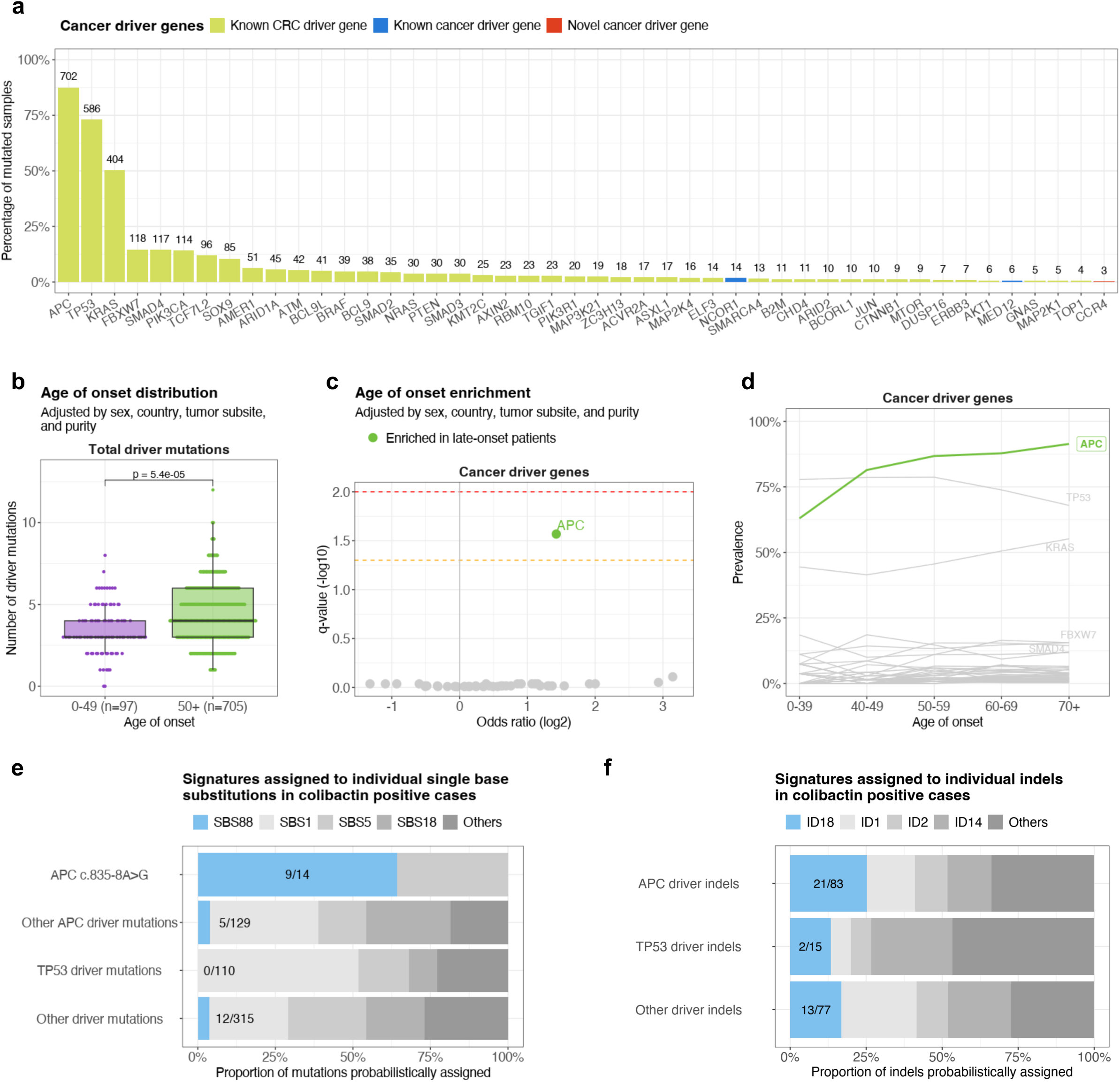
Variation of driver mutations with age of onset and association with colibactin mutagenesis in microsatellite stable colorectal cancers. **a**, Bar plot indicating the prevalence of driver mutations affecting the 48 bioinformatically detected driver genes in microsatellite stable colorectal cancers. Genes were colored according to their status as known cancer driver genes for colorectal cancer, known cancer driver genes for other cancer types, or newly detected cancer driver genes. **b**, Box plots indicating the distribution of total driver mutations across early-onset (under 50 years of age; purple) and late-onset (50 or over; green) tumors. Statistical significance was evaluated using a multivariable linear regression model adjusted by sex, country, tumor subsite, and tumor purity. The line within the box is plotted at the median, while the upper and lower ends indicate the 25^th^ and 75^th^ percentiles. Whiskers show 1.5 × interquartile range, and values outside it are shown as individual data points. **c**, Volcano plot indicating the enrichment of driver mutations in cancer driver genes in early-onset and late-onset cases. Statistically significant enrichments were evaluated using multivariable logistic regression models adjusted by sex, country, tumor subsite, and tumor purity. Firth’s bias-reduced logistic regressions were used for regressions presenting complete or quasi-complete separation. P-values were adjusted for multiple comparisons based on the total number of cancer driver genes considered and reported as q-values. Horizontal lines marking statistically significant thresholds were included at 0.05 (dashed orange line) and 0.01 q-values (dashed red line). **d**, Line plot indicating the prevalence of driver mutations in cancer driver genes across ages of onset, using five different age groups. Cancer driver genes significantly enriched in late-onset cases (as shown in **c**) were colored in green, whereas genes not varying significantly with age of onset were colored in grey. **e**, Bar plots indicating the proportion and number of driver mutations probabilistically assigned to colibactin-induced and other mutational signatures, including single base substitutions (**e**) and small insertions and deletions (indels; **f**). Driver mutations were divided into different groups, including the *APC* c.835-8A>G splicing-associated driver mutation, other *APC* driver mutations, *TP53* driver mutations, and driver mutations affecting other cancer driver genes.

The contributions of SBS88 and ID18 to driver mutations were assessed using probabilistic assignment of signatures to individual mutations^42^. SBS88 accounted for 64.3% of the colibactin-induced^43^ *APC* splicing variant c.835-8A>G in colibactin-exposed samples, compared to only 3.9% and 3.8% of driver substitutions in *APC* or other cancer genes (**Fig. 5*e***). Similarly, ID18 accounted for 25.3% of *APC* driver indels and 16.9% of other driver indels in colibactin-exposed cases (**Fig. 5*f***). Overall, SBS88 and ID18 accounted for 8.3% of all SBS and ID driver mutations, and 15.5% of all *APC* driver mutations in colibactin positive cancers. Nevertheless, no differences were observed between early-onset and late-onset colibactin positive colorectal cancer in the proportion of driver mutations assigned to specific mutational signatures (**Extended Data Fig. 10a-b**). In addition, a prior study observed that SBS88 is also responsible for mutations in chromatin modifier genes^39^, and we were able to validate this as well as show a similar effect for the colibactin-associated indel signature, ID18 (**Extended Data Fig. 10c-d**). Interestingly, using a similar methodology, we observed an elevated number of driver mutations assigned to SBS94 and SBS_F in Colombia, as well as SBS89 and ID_J in Argentina, compared to other countries (**Extended Data Fig. 10e-g**).

## DISCUSSION

Over the last seven decades, colorectal cancer incidence rates have shown complex changes with marked international variation. Notably, while many high-income countries have seen decreases in overall incidence rates, there has been an increase amongst adults under the age of 50. If these trends continue into older age groups, they could reverse the currently overall positive trajectory for colorectal cancer incidence. In this study, whole-genome sequences of 981 colorectal cancers from 11 countries revealed evidence of geographic and age-related variation in their landscapes of somatic mutation, which may contribute to explaining these global trends. These variations were almost exclusively found in the 802 microsatellite-stable colorectal cancers. For colorectal cancers with MSI, limited geographic or age-related differences were observed, possibly due to the smaller sample size and the predominance of somatic mutations resulting from defective DNA repair mechanisms. Similarly, no differences were noted in colorectal cancers harboring other DNA repair deficiencies.

The prevalence of certain mutational signatures, notably SBS89/DBS8/ID_J in Argentina and SBS94/SBS_F/DBS6 in Colombia, was higher in these countries compared to all others. Although such geographic variation could, in principle, be due to differences in population-specific inheritance, it is more plausible that these are due to differences in exogenous environmental or lifestyle mutagenic exposures. Indeed, apart from country of origin, we also assessed the variability with genetic ancestry and self-reported ethnicity (**Methods**), although the homogenous distribution of these characteristics within countries (**Supplementary Fig. 10**) precluded us from clarifying if the varying prevalence of signatures in different countries was related to genetic or environmental factors. The natures of the putative exposures underlying SBS89/DBS8/ID_J and SBS94/SBS_F/DBS6 are currently unknown. However, SBS89 shares several features with colibactin-induced signatures SBS88 and ID18. SBS89 has been previously found in normal colorectal crypts^34^ but not in other normal cells. In individuals with SBS89, some crypts have these mutations while others do not. SBS89 appears to be imprinted on the normal colorectal epithelium early in life, with mutagenesis ceasing thereafter^34^. Moreover, SBS89 mutations show transcriptional strand bias^34^, a common trait of mutations caused by exogenous mutagenic exposures that form bulky covalent DNA adducts. Thus, SBS89 may also be caused by a mutagen originating from the colorectal microbiome and it is conceivable that multiple microbiome-derived mutagens may contribute to the mutation burden of the colorectal epithelium. Although the impact of country-specific microbiome-derived exposures on geographic differences in colorectal cancer incidence remains unclear, the correlations between colorectal cancer ASR and signatures SBS88 and ID18 suggest that microbiome-derived colibactin exposure may influence colorectal cancer incidence rates. Nonetheless, further studies are necessary to thoroughly investigate this hypothesis.

The evidence for enrichment of SBS88 and ID18 mutation burdens in early-onset colorectal cancers may indicate a role for colibactin exposure in the increase in early-onset colorectal cancer incidence over the last 20 years. Prior studies have indicated that mutagenesis due to colibactin exposure can occur within the first decade of life and then ceases^34^. In some instances, the mutation burden caused by this early-life mutation burst can endow affected colorectal crypts with the equivalent of decades of mutation accumulation and, thus, this ‘head start’ could plausibly result in an increased risk of early-onset cancers. One mechanism by which colibactin-induced mutagenesis might contribute to colorectal neoplastic change is by somatically inactivating one copy of *APC* through the generation of protein-truncating driver mutations. Since *APC* mutations usually occur early in the sequence of driver mutations leading to colorectal cancer^38,44^, a first-hit inactivating mutation in *APC* during early life could put an individual several decades ahead for developing colorectal cancer and resulting in a higher likelihood of early-onset colorectal cancer. The mutation profile of SBS88, with its preponderance of T>C substitutions, is intrinsically ineffective in generating translation termination codons and SBS88 accounts for only a small proportion of *APC* driver base substitutions. However, colibactin mutagenesis entails a relatively high proportion of ID mutations, with the characteristic profile of ID18, almost all of which will introduce translational frameshifts in coding sequences. ID18 accounts for approximately one quarter of *APC* indel drivers in colibactin positive cancers and is elevated amongst *APC* indel drivers compared to indel drivers in other cancer genes such as *TP53*, which occur later in the multistep process of colorectal carcinogenesis^45^. Thus colibactin-induced indel driver mutations in *APC* may account for a substantial proportion of any putative impact colibactin exposure has on colorectal carcinogenesis. Conversely, the unexpected increase in driver mutations observed in late-onset colorectal cancers might suggest that we failed to identify all driver mutational events in early-onset cases, possibly overlooking additional effects of colibactin or other mutagenic exposures, and potentially related to alterations beyond *APC*, as early-onset cases are enriched in *APC* wild-type tumors^41^. In this context, BMI, diet, lifestyle, and other exposomal factors— particularly in early life—may play an important mutagenic role, with the lack of analyses on these factors being a limitation of the current study.

Although our results show for the first time an association between the presence of colibactin-induced mutational signatures and early-onset colorectal cancer, complementing the prior finding that tumors harboring colibactin mutagenesis have a younger average age at diagnosis^26^, further research is required to establish causality. Future studies should examine the SBS88 and ID18 mutation burdens of normal colorectal crypts from individuals with early-onset colorectal cancer (cases) and age-matched healthy individuals (controls) with the expectation of an enrichment in cancer cases if colibactin mutagenesis is causally implicated. If so, the increase in early-onset colorectal cancer over the last 30 years would indicate that an increased exposure to colibactin in affected populations occurred during the second half of the 20^th^ century, perhaps due to increasing prevalence of *pks*+ bacteria, and genome sequences of appropriately selected colorectal cancers and normal colorectal tissues would inform on this historical flux. These studies could be supported by international and, if possible, retrospective studies of the prevalence of colibactin-producing *pks+* bacteria in the colorectal microbiome, which should include paired stool samples or other methods for robust microbiome analysis, not available for the current study. Finally, definitive evidence of a causal role for colibactin in early-onset colorectal carcinogenesis would be provided by prevention of early-life exposure to colibactin-producing bacteria reducing cancer incidence.

In summary, mutational epidemiology reveals country-specific and age-specific variations in the prevalence of certain mutational signatures. The results also highlight the potential role of the large intestine microbiome as an early-life mutagenic factor in the development of colorectal cancer.

## EXTENDED DATA FIGURE LEGENDS

**Extended Data Fig. 1. Mutational profiles across molecular subtypes and ages of onset. a-b**, Average mutational profiles of microsatellite stable (MSS; **a**) and microsatellite unstable (MSI; **b**) colorectal tumors for single base substitutions (SBS-288 mutational context), small insertions and deletions (ID-83 mutational context), doublet base substitutions (DBS-78 mutational context), copy number alterations (CN-68 mutational context), and structural variants (SV-38 mutational context). **c-d**, Average mutational profiles of early-onset and late-onset MSS colorectal tumors for doublet base substitutions (**c**) and structural variants (**d**).

**Extended Data Fig. 2. Geographic distribution of mutation burden.** Box plots indicating the distribution of single base substitutions (SBS), small insertions and deletions (ID), doublet base substitutions (DBS), copy number alterations (CN), and structural variants (SV) across countries for microsatellite stable (MSS) colorectal tumors. Box plots and data points representing total number of mutations for each variant type were colored according to each country’s colorectal cancer age-standardized incidence rates (ASR) per 100,000 individuals. A horizontal blue line indicates the median mutation burden for each variant type. Statistically significant differences were evaluated using multivariable linear regression models comparing each country to all others and adjusted by age of diagnosis, sex, tumor subsite, and tumor purity. P-values were adjusted for multiple comparisons based on the total number of countries assessed and reported as q-values. The line within the box is plotted at the median, while the upper and lower ends indicate the 25^th^ and 75^th^ percentiles. Whiskers show 1.5 × interquartile range, and values outside it are shown as individual data points.

**Extended Data Fig. 3. Geographic distribution of mutational profiles. a-e**, Average mutational profiles of microsatellite stable (MSS) colorectal tumors for single base substitutions (SBS-288 mutational context; **a**), small insertions and deletions (ID-83 mutational context; **b**), doublet base substitutions (DBS-78 mutational context; **c**), copy number alterations (CN-68 mutational context; **d**), and structural variants (SV-38 mutational context; **e**).

**Extended Data Fig. 4. Mutational signatures of small mutational events identified in microsatellite stable colorectal cancers. a-c**, Mutational profiles of single base substitution (SBS) signatures, including COSMICv3.4 reference signatures (**a**), previously reported signatures not present in COSMIC (**b**), and novel signature SBS_O (**c**). **d-e**, Mutational profiles of small insertions and deletions (ID) signatures, including COSMICv3.4 signatures (**d**) and novel signature ID_J (**e**). **f**, Mutational profiles of doublet base substitution (DBS) signatures, all previously reported in COSMIC.

**Extended Data Fig. 5. Mutational signatures of large mutational events identified in microsatellite stable colorectal cancers. a-b**, Mutational profiles of copy number (CN) signatures, including COSMICv3.4 reference signatures (**a**) and novel signature CN_F (**b**). **c-d**, Mutational profiles of structural variant (SV) signatures, including COSMIC signatures (**c**) and novel signatures SV_B and SV_D (**d**).

**Extended Data Fig. 6. Association of mutational signatures with colorectal, colon, and rectal cancer incidence rates. a-b**, Scatter plots indicating the association of the mutations attributed to signatures SBS1, SBS_H, and CN_F with the age-standardized incidence rates across countries for colorectal cancers (**a**), and independently for colon and rectal cancer (**b**). Data points were colored based on signature prevalence, with their size indicating the total number of cases per country. Statistically significant associations were evaluated using the sample-level multivariable linear regression models used in **Fig. 2*d*** (**a**), and similar multivariable linear regression models adjusted by age of diagnosis, sex, and tumor purity (**b**). **c**, Bar plots indicating mutational signature prevalence enrichment between low and high ASR countries (defined as those below or above an ASR of 7 per 100,000 people, for early-onset colorectal cancer, diagnosed between 20 and 49 years old). Statistically significant associations were evaluated using multivariable logistic regression models for early-onset colorectal cancer ASR adjusted by age of diagnosis, sex, tumor subsite, and tumor purity.

**Extended Data Fig. 7. Enrichment of colibactin mutagenesis in early-onset colorectal cancers based on motif analysis. a**, Box plots indicating the percentage of total W[T>N]W mutations with the WAWW[T>N]W motif across different age groups. Statistically significant trend was evaluated using a multivariable linear regression model adjusted by sex, country, tumor subsite, and tumor purity. The line within the box is plotted at the median, while the upper and lower ends indicate the 25^th^ and 75^th^ percentiles. Whiskers show 1.5 × interquartile range, and values outside it are shown as individual data points. **b**, Box plots indicating the percentage of total W[T>N]W mutations with the WAWW[T>N]W motif across samples grouped by colibactin exposure status, determined by the presence of signatures SBS88 or ID18. Statistical significance was evaluated using a multivariable linear regression model adjusted by age, sex, country, tumor subsite, and tumor purity. **c**, Bar plots indicating the prevalence of colibactin exposure across age groups, with indication of the total number of cases where colibactin signatures were detected. Statistically significant trend was evaluated using a multivariable logistic regression model adjusted by sex, country, tumor subsite, and tumor purity.

**Extended Data Fig. 8. Enrichment of colibactin mutagenesis as an early clonal event in early-onset and late-onset colorectal cancers. a**, Box plots indicating the fold-change of the relative contribution per sample of each signature between clonal and subclonal single base substitutions (SBS, left) and small insertions and deletions (ID, right). Signatures that generated clonal somatic mutations in fewer than 10 samples and also generated subclonal somatic mutations in fewer than 10 samples were excluded from the analysis. The line within the box is plotted at the median, while the upper and lower ends indicate the 25^th^ and 75^th^ percentiles. Whiskers show 1.5 × interquartile range, and values outside it are shown as individual data points. **b**, Boxplots indicating the fold-change of the relative contribution per sample of each signature between early clonal and late clonal SBS (left) and ID (right) with samples separated by age of diagnosis in early-onset (under 50 years of age; purple) and late-onset (50 or over; green). As in **Fig. 4*a***, SBS signatures that generated early clonal SBSs in fewer than 50 samples and late clonal SBSs in fewer than 50 samples, as well as ID signatures generating early clonal IDs in fewer than 20 samples and late clonal IDs in fewer than 20 samples, were excluded from the analysis.

**Extended Data Fig. 9. Representative microbiome and genomic profiles of colibactin-exposed samples. a-d**, Microbiome and genomic profiles of representative samples corresponding to the four different sample types according to colibactin exposure: genomic+ and *pks+* (**a**), genomic+ and *pks-* (**b**), genomic- and *pks+* (**c**), and genomic- and *pks-*. The genomic status is defined by the presence of SBS88 or ID18 signatures, while the microbiome status (*pks*) is determined by the coverage of at least half of the *pks* island, and suggests ongoing and/or active *pks*+ bacterial infection. Circos plots display Reads Per Kilobase of transcript per Million (RPKM) values across *clb* genes within the *pks* island (left). Bar plots represent the proportion of mutations attributed to SBS88 and ID18 colibactin signatures compared to others (center), and are displayed next to mutational profiles of single base substitutions (SBS-288 mutational context) and small insertions and deletions (ID-83 mutational context) for each sample (right).

**Extended Data Fig. 10. Driver mutations associated with colibactin mutagenesis in early-onset and late-onset colibactin positive cases and with country-enriched mutational signatures in microsatellite stable colorectal cancers. a-b**, Bar plots indicating the proportion and number of driver mutations probabilistically assigned to colibactin-induced and other mutational signatures, including single base substitutions (**a**) and small insertions and deletions (indels; **b**), with samples separated by age of diagnosis in early-onset (under 50 years of age; left) and late-onset (50 or over; right). Driver mutations were divided into different groups, including the *APC* c.835-8A>G splicing-associated driver mutation, other *APC* driver mutations, *TP53* driver mutations, and driver mutations affecting other cancer driver genes. **c-d**, Bar plots indicating the proportion and number of mutations in chromatin modifier genes probabilistically assigned to colibactin-induced and other mutational signatures, including single base substitutions (**c**) and indels (**d**), in the 169 colibactin positive cases. **e-g**, Bar plots indicating the proportion and number of driver single base substitutions (**e** and **f**) and indels (**g**) in cancer driver genes probabilistically assigned to specific mutational signatures in cases from Colombia (**e**) or Argentina (**f** and **g**) compared to other countries.

## SUPPLEMENTARY FIGURE LEGENDS

**Supplementary Fig. 1. Mutational profiles of homologous recombination deficient colorectal cancers. a**, Mutational profiles of individual samples identified as homologous recombination deficient for single base substitutions (SBS-288 mutational context) and small insertions and deletions (ID-83 mutational context). **b**, Mutational signatures previously associated with homologous recombination deficiency in COSMICv3.4 (SBS3 and ID6).

**Supplementary Fig. 2. Mutational profiles of base excision repair deficient and *POLD1* mutated colorectal cancers. a**, Mutational profile of an individual sample identified as base excision repair deficient due to mutations in *MUTYH* for single base substitutions (SBS-288 mutational context). **b**, Mutational signature previously associated with base excision repair deficiency due to mutations in *MUTYH* in COSMICv3.4 (SBS36). **c**, Mutational profiles of individual samples identified as base excision repair deficient due to mutations in *NTHL1* for single base substitutions (SBS-288 mutational context). **d**, Mutational signature previously associated with base excision repair deficiency due to mutations in *NTHL1* in COSMICv3.4 (SBS30). **e**, Mutational profile of an individual sample identified as base excision repair deficient due to mutations in *OGG1* for single base substitutions (SBS-288 mutational context). **f**, Mutational signature previously associated with base excision repair deficiency due to mutations in *OGG1* (Signal signature SBS108). **g**, Mutational profiles of individual samples harboring mutations in *POLD1* for single base substitutions (SBS-288 mutational context). **h**, Mutational signature previously associated with mutations in *POLD1* in COSMICv3.4 (SBS10c).

**Supplementary Fig. 3. Mutational profiles of *POLE* mutated colorectal cancers. a**, Mutational profiles of individual samples harboring mutations in *POLE* for single base substitutions (SBS-288 mutational context). **b**, Mutational signatures previously associated with mutations in *POLE* in COSMICv3.4 (SBS10a, SBS10b, and SBS28).

**Supplementary Fig. 4. Mutational profiles of cases treated with chemotherapy for prior cancers. a**, Mutational profiles of individual samples treated with chemotherapy for prior cancers for single base substitutions (SBS-288 mutational context) and doublet base substitutions (DBS-78 mutational context). **b**, Mutational signatures previously associated with chemotherapy in COSMICv3.4 (SBS25, SBS31, SBS35, and DBS5).

**Supplementary Fig. 5. Prevalence of mutational signatures in microsatellite stable colorectal cancers by country. a-e**, SBS (**a**), ID (**b**), DBS (**c**), CN (**d**), and SV (**e**) signatures.

**Supplementary Fig. 6. Main age association analyses adjusted by self-reported ethnicity instead of country of origin. a-e**, Replicates of Fig. 1d (**a**), Fig. 3a (**b**), and Fig. 3c-e (**c-e**).

**Supplementary Fig. 7. Main age association analyses adjusted by the first five principal components of the genetic ancestry analysis instead of country of origin. a-e**, Replicates of Fig. 1d (**a**), Fig. 3a (**b**), and Fig. 3c-e (**c-e**).

**Supplementary Fig. 8. Main age association analyses adjusted by genetic ancestry groups (ADMIX, AFR, EAS, EUR) instead of country of origin. a-e**, Replicates of Fig. 1d (**a**), Fig. 3a (**b**), and Fig. 3c-e (**c-e**).

**Supplementary Fig. 9. Variation of mutational signatures associated with earlier age of onset with tumor subsite in microsatellite stable colorectal cancers.**

**Supplementary Fig. 10. Genetic ancestry (a) and self-reported ethnicity (b) distribution by country**.

**Supplementary Fig. 11. Comparison of clinicopathological characteristics at baseline by country, comparing Mutographs samples vs. GLOBOCAN-based expectations.**

**Supplementary Fig. 12. Mutational signature reconstruction of an individual microsatellite unstable tumor not validated by droplet digital PCR.** Multiple mutational signatures were assigned to this case, as indicated in **Supplementary Note Table 8**, including microsatellite instability-associated COSMICv3.4 signature SBS15 and SBS26, suggesting the presence of microsatellite instability.

**Supplementary Fig. 13. Novel mutational signatures identified in microsatellite unstable colorectal cancers. a-b**, Mutational profiles of single base substitution (SBS) signatures not matching any previous COSMICv3.4 signature, including three novel signatures (**a**) and a previously reported signature (**b**). **c**, Mutational profile of a novel doublet base substitution (DBS) signature not previously reported in COSMIC. **d-e**, Exemplar mutational profiles of individual colorectal cancers supporting the three novel signatures indicated in **a** (**d**) and the previously reported signature in **b** (**e**).

**Supplementary Fig. 14. Variation of germline pathogenic variants with age of onset. a-b**, Volcano plots indicating the enrichment of pathogenic or likely pathogenic germline variants in early-onset and late-onset cases in microsatellite stable (MSS; **a**) and unstable colorectal cancers (MSI; **b**). Separate analyses were performed for all individual genes (left), and for genes grouped in colorectal cancer predisposition syndromes (Lynch syndrome and Cowden syndrome), homologous recombination, or DNA damage repair-associated genes (right). Statistically significant enrichments were evaluated using multivariable logistic regression models for age of onset categorized in two subgroups (early-onset, <50 years of age; and late-onset, ≥50) and adjusted by sex, country, and tumor subsite, and tumor purity. Firth’s bias-reduced logistic regressions were used for regressions presenting complete or quasi-complete separation. P-values were adjusted for multiple comparisons based on the total number of germline variants considered and reported as q-values. Horizontal lines marking statistically significant thresholds were included at 0.05 (dashed orange line) and 0.01 q-values (dashed red line).

**Supplementary Fig. 15. Dendrogram of the relative abundances of the considered bacterial genus based on the Bray-Curtis distance.**

**Supplementary Fig. 16. Single base substitution mutational signatures extracted by SigProfilerExtractor using the SBS-288 and SBS-1536 mutational contexts in microsatellite**

**stable colorectal cancers.** All single base substitution (SBS) *de novo* signatures extracted using the SBS-288 (16 signatures) and SBS-1536 (14 signatures) mutational contexts, shown side by side for comparison. Equivalent signatures were not extracted in SBS-1536 format for SBS288E and SBS288K. For clarity, the signature context is retained in the signature names in this figure. The extended context for SBS-1536 signatures is omitted from the figure. Instead, the SBS-96 down-sampled version of the SBS-1536 *de novo* extracted signatures was used to display the signatures.

**Supplementary Fig. 17. Single base substitution mutational signatures extracted by mSigHdp in microsatellite stable colorectal cancers.** Seventeen single base substitution (SBS) *de novo* signatures were extracted by mSigHdp, using the SBS-96 mutational context.

**Supplementary Fig. 18. Small insertion and deletion mutational signatures extracted by SigProfilerExtractor and mSigHdp in microsatellite stable colorectal cancers. a**, Ten small insertions and deletions (ID) *de novo* signatures were extracted by SigProfilerExtractor using the ID-83 mutational context. **b**, Eight ID *de novo* signatures were extracted by mSigHdp.

**Supplementary Fig. 19. Sensitivity analysis for the detection of colibactin signatures in microsatellite stable colorectal cancers. a-b**, Sensitivity analysis for SBS88 (**a**) and ID18 (**b**) detection, including average relative activity of the signature detected across all colibactin negative samples and simulations (top), activity of the signature per sample across all colibactin negative samples and simulations (middle), and activity of the signature per sample across all colibactin positive samples compared to the median detection of the signature in the simulated data (bottom).

## SUPPLEMENTARY TABLES

**Supplementary Table 1. Summary of incidence rates, sex, age, tumor subsite, and molecular subgroups across countries included in the Mutographs colorectal cancer cohort.**

**Supplementary Table 2. Germline mutations in mismatch repair genes in MSI Lynch syndrome cases.**

**Supplementary Table 3. Germline and somatic mutations in DNA repair deficient cases.**

**Supplementary Table 4. Detection results of homologous recombination deficiency with CHORD.**

**Supplementary Table 5. Mutational profiles of *de novo* SBS signatures extracted in MSS colorectal cancer cases.**

**Supplementary Table 6. Mutational profiles of *de novo* ID signatures extracted in MSS colorectal cancer cases.**

**Supplementary Table 7. Mutational profiles of *de novo* DBS signatures extracted in MSS colorectal cancer cases.**

**Supplementary Table 8. Mutational profiles of *de novo* CN signatures extracted in MSS colorectal cancer cases.**

**Supplementary Table 9. Mutational profiles of *de novo* SV signatures extracted in MSS colorectal cancer cases.**

**Supplementary Table 10. Decomposition of *de novo* MSS colorectal cancer signatures into previously reported signatures.**

**Supplementary Table 11. Sample attributions of decomposed SBS signatures in MSS colorectal cancers.**

**Supplementary Table 12. Sample attributions of decomposed ID signatures in MSS colorectal cancers.**

**Supplementary Table 13. Sample attributions of decomposed DBS signatures in MSS colorectal cancers.**

**Supplementary Table 14. Sample attributions of decomposed CN signatures in MSS colorectal cancers.**

**Supplementary Table 15. Sample attributions of decomposed SBS signatures in SV colorectal cancers.**

**Supplementary Table 16. Enrichment of mutational signature prevalence in specific countries compared to all others in MSS colorectal cancers.**

**Supplementary Table 17. Enrichment of mutational signature activities with colorectal cancer incidence in MSS colorectal cancers.**

**Supplementary Table 18. Enrichment of mutational signature activities with colon and rectal cancer incidence in MSS colon and rectal cancers.**

**Supplementary Table 19. Enrichment of mutational signature prevalence in late-onset compared to early-onset MSS colorectal cancers.**

**Supplementary Table 20. Trend enrichment of mutational signature prevalence with age of diagnosis in MSS colorectal cancers.**

**Supplementary Table 21. Driver genes detected in MSS colorectal cancers.**

**Supplementary Table 22. Enrichment of driver mutations in cancer driver genes in late-onset compared to early-onset MSS colorectal cancers.**

**Supplementary Table 23. Enrichment of hotspot driver mutations in late-onset compared to early-onset MSS colorectal cancers.**

**Supplementary Table 24. Enrichment of driver mutations in cancer driver genes in specific countries compared to all others in MSS colorectal cancers.**

**Supplementary Table 25. Enrichment of hotspot driver mutations in specific countries compared to all others in MSS colorectal cancers.**

**Supplementary Table 26. Details of individual case collections.**

**Supplementary Table 27. Clinicopathological characteristics of included and excluded cases.**

## ONLINE METHODS

### Recruitment of patients and informed consent

The International Agency for Research on Cancer (IARC/WHO) coordinated case recruitment through an international network of 17 collaborators from 11 participating countries in North America, South America, Asia, and Europe (**Supplementary Table 26**). The inclusion criteria for patients were ≥18 years of age (ranging from 18 to 95, with a mean of 64 and a standard deviation of 12), confirmed diagnosis of primary colorectal cancer, and no prior treatment for colorectal cancer. Informed consent was obtained for all participants. Patients were excluded if they had any condition that could interfere with their ability to provide informed consent or if there were no means of obtaining adequate tissues or associated data as per the protocol requirements. Ethical approvals were first obtained from each Local Research Ethics Committee and Federal Ethics Committee when applicable, as well as from the IARC/WHO Ethics Committee.

### Bio-samples and data collection

Dedicated standard operating procedures, following guidelines from the International Cancer Genome Consortium (ICGC), were designed by IARC/WHO to select appropriate case series with complete biological samples and exposure information^46^, as described previously^5,6,8^ (**Supplementary Table 26**). In brief, for all case series included, anthropometric measures were taken, together with relevant information regarding medical and familial history. All biological samples from retrospective cohorts were collected using rigorous, standardized protocols and fulfilled the required standards of sample collection defined by the IARC/WHO for sequencing and analysis. Potential limitations of using retrospective clinical data collected using different protocols from different populations were addressed by a central data harmonization to ensure a comparable group of exposure variables (**Supplementary Table 26**). All patient-related data were pseudonymized locally through the use of a dedicated alpha-numerical identifier system before being transferred to the IARC/WHO central database.

### Expert pathology review

Original diagnostic pathology departments provided diagnostic histological details of contributing cases through standard abstract forms, together with a representative hematoxylin-eosin-stained slide of formalin-fixed paraffin-embedded tumor tissues whenever possible. IARC/WHO centralized the entire pathology workflow and coordinated a centralized digital pathology examination of the frozen tumor tissues collected for the study as well as formalin-fixed paraffin-embedded sections when available, via a web-based approach and dedicated expert panel following standardized procedures as described previously^5,6^. A minimum of 50% viable tumor cells was required for eligibility for whole-genome sequencing. In summary, frozen tumor tissues were first examined to confirm the morphological type and the percentage of viable tumor cells. A random selection of tumor tissues was independently evaluated by a second pathologist. Enrichment of tumor component was performed by dissection of the non-tumoral part, if necessary.

### DNA extraction

A total of 1,977 primary colorectal cancer patients were enrolled into the study, including biological samples for 1,946 cases and sequencing data (FASTQ) for 31 cases from Japan. Of these, 906 samples (45.8%) were excluded due to insufficient viable tumor cells (pathology level) or inadequate DNA (tumor or germline). Extraction of DNA from fresh frozen primary tumor and matched blood/normal tissue samples was centrally conducted at IARC/WHO (except for samples from Japan) following a similarly standardized DNA extraction procedure. Germline DNA was extracted from whole blood (*n*=1,015), except for a small subset of Canadian cases (*n*=25) where only adjacent normal tissue was available, following previously described protocols and methods^5,6^. As a result, DNA from 1,040 cases was sent to the Wellcome Sanger Institute for whole-genome sequencing.

### Whole-genome sequencing

Fluidigm SNP genotyping with a custom panel was performed to ensure that each pair of tumor and matched normal samples originated from the same individual. Whole-genome sequencing (150bp paired-end) was performed on the Illumina NovaSeq 6000 platform with a target coverage of 40x for tumors and 20x for matched normal tissues. All sequencing reads were aligned to the GRCh38 human reference genome using the Burrows-Wheeler Aligner MEM (BWA-MEM; v0.7.16a and v0.7.17)^47^. Post-sequencing quality control metrics were applied for total coverage, evenness of coverage, contamination, and total number of somatic single base substitutions (SBSs). Cases were excluded if coverage was below 30x for tumor or 15x for normal tissue. For evenness of coverage, the median over mean coverage (MoM) score was calculated. Tumors with MoM scores outside the range of values determined by previous studies^48^ to be appropriate for whole-genome sequencing (0.92-1.09) were excluded. Conpair^49^ was used to detect contamination, cases were excluded if the result was greater than 3%^48^. Lastly, samples with <1,000 total somatic SBSs were also excluded. A total of 981 pairs of colorectal cancer and matched-normal tissue passed all criteria. Comparing the clinicopathological characteristics between the included and excluded patients revealed very similar traits (**Supplementary Table 27**), and comparable to those expected for each country according to GLOBOCAN metrics (obtained from https://gco.iarc.who.int/today/en/dataviz/; **Supplementary Fig. 11**).

### Germline variant calling

Germline SNVs and indels were derived from whole-genome sequencing from the normal paired material for each individual using Strelka2 with appropriate quality-control criteria^50^. Variant calls were then derived into genotypes for each individual and annotated using ANNOVAR^51^.

### Somatic variant calling

Variant calling was performed using the standard Sanger bioinformatics analysis pipeline (https://github.com/cancerit). Copy number profiles were determined using ASCAT^52^ and BATTENBERG^53^ when tumor purity allowed. SNVs were called with cgpCaVEMan^54^, indels were called with cgpPINDEL^55^, and structural rearrangements were called using BRASS (https://github.com/cancerit/BRASS). CaVEMan and BRASS were run using the copy number profile and purity values determined from ASCAT when possible (complete pipeline, *n*=916). When tumor purity was insufficient to determine an accurate copy number profile (partial pipeline, *n*=31) CaVEMan and BRASS were run using copy number defaults and an estimate of purity obtained from ASCAT. Finally, for a subset of cases which had no large copy number alterations (copy number normal pipeline, *n=34*), CaVEMan and BRASS were run using copy number defaults and an estimate of purity calculated by the median variant allele frequency (VAF) of indels multiplied by two. For SNVs, additional filters (ASRD≥140 and CLPM=0) were applied in addition to the standard PASS filter to remove potential false positive calls. To further exclude the possibility of caller-specific artifacts being included in the analysis, a second variant caller, Strelka2^50^, was run for SNVs and indels. Only variants called by both the Sanger variant calling pipeline and Strelka2 were included in subsequent analysis.

### Generation of mutational matrices

Mutational matrices for single base substitutions (SBS), indels (ID), doublet base substitutions (DBS), copy number alterations (CN), and structural variants (SV) were generated using SigProfilerMatrixGenerator with default options (v1.2.0)^56,57^.

### Microsatellite instability validation

The presence of microsatellite instability (MSI) in colorectal cancers was validated using the QX200 Droplet Digital PCR System (Bio-Rad, Hercules, CA, USA) for the detection of five microsatellite markers (BAT25, BAT26, NR21, NR24, and Mono27) commercially pooled in three primer–probe mix assays, as previously described^58^. Briefly, samples were tested in duplicate, and each reaction comprised 1× ddPCR Multiplex Supermix for probes (Bio-Rad), 1X primer–probe mix, and 10 ng of extracted tumor DNA, in a total volume of 22 μl. MSI-positive, negative, and no-template (nuclease-free water) controls were included in each experiment. Droplet generation and plate preparation for thermal cycling amplification were performed using the QX200 AutoDG Droplet Digital PCR System (Bio-Rad). The following PCR protocol was applied on a C1000 Touch Thermal Cycler (Bio-Rad): 37 °C for 30 min, 95 °C for 10 min, followed by 40 cycles of denaturation at 94 °C for 30 s, annealing at 55 °C for 1 min, with a final extension at 98 °C for 10 min. Following PCR amplification, fluorescence signals were quantified using the QX200 Droplet Reader (Bio-Rad), and data were analyzed with QuantaSoft Analysis Pro v.1.0.596.0525 (Bio-Rad) software. Positive and negative controls served as guides to call markers and delineate clusters. For each assay, the cluster at the bottom left of the x–y plot was designated as the negative population. Clusters located vertically and horizontally from the negative cluster were identified as the mutant population, while clusters located diagonally from the negative cluster represented the wild-type population. Tumors were characterized for the MSI phenotype by analyzing the results for all five markers using the following criteria: MSI positive if two or more mutant microsatellite markers were observed, and microsatellite stable (MSS; *i.e.,* MSI negative) when none or only one of the microsatellite markers was altered (**Supplementary Note**).

### Extraction and decomposition of mutational signatures

Mutational signatures were primarily extracted using SigProfilerExtractor^35^, based on nonnegative matrix factorization, and validated by mSigHdp^59^, based on hierarchical Dirichlet process mixture models. For SigProfilerExtractor (v1.1.21), *de novo* mutational signatures were extracted from SBS, DBS, and ID mutational matrices using 500 NMF replicates (nmf_replicates=500), nndsvd_min initialization (nmf_init=”nndvsd_min”), and default parameters. Extractions were performed separately on the subsets of 802 MSS and 153 MSI cases (**Supplementary Note**). *De novo* SBS mutational signatures were extracted for both SBS-288 and SBS-1536 contexts, which, beyond the common SBS-96 trinucleotide context using the mutated base and the 5’ and 3’ adjacent nucleotides^57,60^, also consider the transcriptional strand bias and the pentanucleotide context (two 5’ and 3’ adjacent nucleotides), respectively^57^. The results were largely concordant, with the SBS-288 *de novo* signatures allowing additional separation of mutational processes. The SBS-1536 results can be found in the **Supplementary Note**. Therefore, the SBS-288 *de novo* signatures were taken forward for further analysis (**Supplementary Table 5**). Previously established mutational contexts DBS-78 and ID-83^19,57^ were used for the extraction of DBS and ID signatures (**Supplementary Tables 6-7**).

Copy number signatures were extracted *de novo* using SigProfilerExtractor with default parameters and following an updated context definition benefitting from WGS data (CN-68) (**Supplementary Table 8**), which allowed to further characterize CN segments below 100kbp in length (in contrast to current COSMICv3.4 reference signatures using the CN-48 context, which were based on SNP6 microarray data and therefore without the resolution to characterize short CN segments)^61^. SV signatures were extracted using a similarly refined context, with an in-depth characterization of short SV alterations below 1kbp (SV-38 context, in contrast to current COSMICv3.4 signatures based on the SV-32 context^62^; **Supplementary Table 9**).

After *de novo* extraction was completed, SigProfilerAssignment^42^ v0.0.29 was used to decompose the *de novo* extracted SBS, ID, DBS, CN, and SV mutational signatures into COSMICv3.4 reference signatures based on the GRCh38 reference genome^63^ (**Supplementary Table 10**). When possible, SigProfilerAssignment matched each *de novo* extracted mutational signature to a set of previously identified COSMICv3.4 signatures (**Supplementary Note**). For the SBS-288, CN-68, and SV-38 signatures, this required collapsing the high-definition classifications into the standard SBS-96, CN-48, and SV-32 mutational classifications, respectively. Four of the *de novo* extracted MSS SBS signatures did not match any previous COSMICv3.4 signatures, with three of them (SBS_F, SBS_H, and SBS_M) showing a strong similarity with previously reported signatures in the UK population^21^ (cosine similarity > 0.93), and one (SBS_O) reflecting a cleaner version of a previously reported COSMICv3.4 signature (SBS41). To validate the latter, we performed a decomposition of the current mutational profile of signature SBS41 using the decomposed signatures from our analysis, obtaining a confirmation that SBS41 can be reconstructed by a linear combination of SBS_O (contributing 19.00% of the mutational profile), SBS93 (62.54%), and SBS34 (12.60%), and SBS5 (5.86%) with a cosine similarity of 0.91. Notably, SBS93, first identified in gastric tumors^35^, was unknown at the time SBS41 was first reported^19^. For the MSS cohort, one ID (ID_J), one CN (CN_F), and two SV signatures (SV_B and SV_D) were additionally not decomposed into previously known signatures, and therefore considered as novel (**Supplementary Table 10**). The novel SV signature SV_D, identified in the MSS cohort, was also considered for the decomposition of *de novo* SV signatures extracted in the MSI cohort. In the MSI cohort, four of the *de novo* extracted SBS signatures (SBS_I_MSI, SBS_M_MSI, M, SBS_N_MSI, and SBS_O_MSI) as well as one *de novo* DBS signature (DBS_B_MSI) did not match COSMICv3.4 signatures, with SBS_M_MSI showing a strong similarity with a previously reported signature in the UK population^21^ (cosine similarity=0.89), and the other four signatures considered as novel (**Supplementary Note**).

mSigHdp^59^ extraction of SBS-96 and ID-83 signatures was performed on the 802 MSS subset using the suggested parameters and using the country of origin to construct the hierarchy. SigProfilerAssignment was subsequently used to match mSigHdp *de novo* signatures to previously identified COSMIC signatures. A comparison of the signatures extracted from mSigHdp and SigProfilerExtractor can be found in the **Supplementary Note**.

### Attribution of mutational signatures to individual samples

Known COSMIC signatures and *de novo* signatures that were not decomposed into COSMIC signatures (**Supplementary Table 10**; **Supplementary Note**) were attributed for each sample using MSA^64^ (v2.0) for SBS, ID, and DBS, whereas SigProfilerAssignment^42^ was used for CN and SV. A conservative approach was used for MSA attributions utilizing the (params.no_CI_for_penalties=False) option for the calculation of optimum penalties. Pruned attributions were used for the final analysis, where confidence intervals were applied to each attributed mutational signature and any signature activity with a lower confidence limit equal to 0 was removed.

### Attribution of mutational signatures to individual somatic mutations

SBS and ID mutational signatures were probabilistically attributed to individual somatic mutations using the MSA activities per sample, based on Bayes’ rule and the specific mutational context for the mutation, as previously described^42^. Briefly, to calculate the probability of a specific mutational signature being responsible for a mutation in a given mutational context and in a particular sample, we multiplied the general probability of the signature causing mutations in a specific mutational context (obtained from the mutational signature profile) by the activity of the signature in the sample (obtained from the signature activities), and then normalized this value dividing by the total number of mutations corresponding to the specific mutational context (obtained from the reconstructed mutational profile of the sample). The signature with the maximum likelihood estimation was assigned to each individual somatic mutation.

### Driver gene analysis

Consensus *de novo* driver gene identification was performed by IntOGen^40^, which combines seven state-of-the-art computational methods to detect signals of positive selection across the cohort. The genes identified as drivers with a combination q-value<0.10 were classified according to their mode of action in tumorigenesis (*i.e.*, tumor suppressor genes or oncogenes) based on the relationship between the excess of observed nonsynonymous and truncating mutations computed by dNdScv^65^ and their annotations in the Cancer Gene Census^66^.

To identify potential driver mutations, we selected SBS or ID mutations that fulfilled any of the following criteria: mutations classified as “Oncogenic” or “Likely Oncogenic” by OncoKB^67^; mutations classified as drivers in the TCGA MC3 drivers study^68^; truncating mutations in driver genes annotated as tumor suppressors; recurrent missense mutations (seen in at least three cases); mutations classified as “Likely Drivers” by boostDM (score >0.50)^69^; or missense mutations classified as “Likely Pathogenic” by AlphaMissense^70^ in driver genes annotated as tumor suppressors. Six of the IntOGen-identified driver genes did not carry any potential driver mutations according to our strict criteria and were therefore excluded from subsequent analysis. In summary, 60 driver genes were identified (46 and 31 for MSS and MSI cases, respectively; **Supplementary Table 21**; **Supplementary Note**).

### Evolutionary analysis

DPClust^53^ was run on all complete pipeline MSS samples with Battenberg data (*n*=774) to identify clonal structure in each sample. The DPClust output was used in running MutationTimeR^38^ to annotate somatic mutations as early clonal, late clonal, subclonal, or NA clonal. Samples with at least 256 early clonal and late clonal SBSs or 100 early clonal and late clonal IDs were retained and split into separate VCF files (*n*=574 for SBS; *n*=430 for ID). MSA^64^ was run on the resulting VCF files to identify the active mutational signatures in the early clonal and late clonal mutations. SBS signatures that were found to generate early clonal SBSs in fewer than 50 samples and also generated late clonal SBSs in fewer than 50 samples were excluded from the analysis. Similarly, ID signatures generating early clonal IDs in fewer than 20 samples and late clonal IDs in fewer than 20 samples were also excluded. Wilcoxon signed-rank tests were used to assess the differences in the relative activity of each signature between the early clonal and late clonal mutations. P-values were adjusted across signatures using the Benjamini-Hochberg method^71^, and adjusted p-values were reported as q-values. This process was repeated with the same thresholds for SBSs and IDs to also assess the difference in the relative activity of each signature between clonal and subclonal mutations (*n*=133 for SBS; *n*=64 for ID). Due to the lower numbers, signatures that were found to generate clonal somatic mutations in fewer than 10 samples and also generated subclonal somatic mutations in fewer than 10 samples were excluded from the analysis.

### Motif analysis

MutaGene^72^ was used to find the number of mutations with the WAWW[T>N]W motif, previously associated with colibactin mutagenesis^33^, in each sample, regardless of the DNA strand. This value was then divided by the total number of W[T>N]W mutations per sample to identify the percentage of W[T>N]W mutations with the colibactin mutational motif.

### Microbiome analysis

To identify microbial reads that map to the pks island (*pks*), non-human reads were aligned to the IHE3034 genome (RefSeq assembly: GCF_000025745.1) using Bowtie2^73^. IHE3034 is a *pks E. coli* strain that contains the *pks* island with all 19 *clb* genes in the *clbA*-*clbS* gene cluster. Prior to alignment, poor quality reads were filtered using fastp^74^, and the remaining human reads were removed by excluding those that mapped to GRCh38, T2T-CHM13v2.0, and the 47 pangenomes^75^. A sample was considered *pks+* if it had at least one read across at least 8 out of the 19 genes in the *clbA*-*clbS* gene cluster. Genome coverage circos plots were generated using Reads Per Kilobase per Million (RPKM) values and visualized with the *circlize* R package^76^.

### Regressions

To compare the mutation burden of different variant types, a linear regression of the mutation burden logarithm (base 10) was considered, using age, sex, tumor subsite, country, and tumor purity as independent variables. For mutational signature-based analyses, signature attributions were dichotomized into presence and absence using confidence intervals, with presence defined as both lower and upper limits being positive and absence as the lower limit being zero. If a signature was present in at least 70% of cases (SBS1, SBS5, SBS18, ID1, ID2, ID14 and CN2 for MSS cases; ID1, ID2, DBS_B_MSI, CN1, and SV_D for MSI cases), it was dichotomized into above and below the median of attributed mutation counts. The binary attributions served as dependent variables in logistic regressions. Regressions with variables presenting complete or quasi-complete separation^77^ were performed using Firth’s bias-reduced logistic regressions based on the *logistf* R package. To adjust for confounding factors, sex, age of diagnosis, tumor subsite, country, and tumor purity were added as covariates in all regressions, serving as independent variables for the regressions. The tumor subsite variable was categorized as proximal colon (ICD-10-CM codes C18.0, C18.2, C18.3, and C18.4), distal colon (C18.5, C18.6, and C18.7), or rectum (C19 and C20), unless otherwise specified. One MSI tumor from an unspecified subsite was removed for the multivariable regression models in MSI cases. The age of diagnosis variable was generally considered as a numerical variable, or categorized into two (early-onset, <50 years old; and late-onset, ≥50) or five subgroups (0-39, 40-49, 50-59, 60-69, ≥70), depending on the analysis performed, with specific indications in the corresponding figure legends. Similarly, regressions for driver mutations in cancer driver genes and hotspot driver mutations (present in at least 10 cases) were done using the same logistic regression models but replacing signature by driver mutation prevalence across samples.

Regressions with colorectal cancer incidence were performed as linear regressions with signature attributions with confidence intervals not consistent with zero as dependent variables, and age-standardized rates (ASR) of colorectal cancer (and independent ASR of colon and rectal cancer) obtained from the Global Cancer Observatory (GLOBOCAN)^1^, sex, age of diagnosis, tumor subsite, and tumor purity as independent variables. Regressions were performed on a sample basis.

Regressions with colibactin presence (based on genomic and/or microbiome-derived detection) were performed as linear regressions with age of diagnosis as the dependent variable, and sex, tumor subsite, country, and tumor purity as independent variables.

### Additional statistical analyses

For regressions of signatures, driver mutations in cancer driver genes, and hotspot driver mutations, p-values were adjusted for multiple comparisons based on the total number of decomposed reference mutational signatures considered per variant type (*i.e.*, 19 SBS, 7 DBS, 11 ID, 9 CN, and 11 SV signatures for MSS cases; 18 SBS, 10 DBS, 2 ID, 4 CN, and 4 SV for MSI cases), cancer genes (46 for MSS; 31 for MSI), or hotspot driver mutations (38 for MSS; 14 for MSI) using the Benjamini-Hochberg method^71^. For country enrichment analyses, the mutation burdens and binary attributions of mutational signatures were compared for each country against all others. Therefore, p-values were also adjusted for multiple comparisons based on the total number of countries assessed (a total of 11 countries). Adjusted p-values were reported as q-values, with q-values<0.05 considered statistically significant. For age of diagnosis-based regressions of colibactin presence across tumor subsites, p-values were adjusted and reported as q-values based on the total number of tumor subsites assessed (a total of 3 tumor subsites). For the age of diagnosis trend enrichment analysis of signatures, p-trends were reported, with p-trends<0.05 considered statistically significant. For evidence of co-occurrence or mutual exclusivity of two signatures, two-sided Fisher’s exact tests were used, and p-values were reported, with p-values<0.05 considered statistically significant.

## DATA AVAILABILITY

Whole-genome sequencing data, somatic mutations, and patient metadata are deposited in the European Genome-phenome Archive (EGA) associated with study EGAS00001003774. All other data is provided in the accompanying Supplementary Tables.

## CODE AVAILABILITY

All algorithms used for data analysis are publicly available with repositories noted within the respective method sections. The code used for regression analysis and figures is available at https://github.com/AlexandrovLab/Mutographs_CRC.

## ACKNOWLEDGMENTS

The authors thank the IARC General Services, including the Laboratory Services and Biobank team led by Z. Kozlakidis and the Section of Support to Research overseen by C. Mehta under IARC regular budget funding for the support provided; Laura O’Neill, Kirsty Roberts, Katie Smith, Siobhan Austin-Guest, and the staff of Sequencing Operations at the Wellcome Sanger Institute for their contribution; Laura Rodríguez Porras for her help in designing and reviewing the figures; the work of all other collaborators in the Mutographs project who participated in the recruitment of patients in all centers; and all the patients involved in this study and their families. The computational analyses reported in this manuscript have utilized the Triton Shared Computing Cluster at the San Diego Supercomputer Center of UC San Diego. Where authors are identified as personnel of the International Agency for Research on Cancer / World Health Organization, the authors alone are responsible for the views expressed in this article and they do not necessarily represent the decisions, policy or views of the International Agency for Research on Cancer / World Health Organization.

## AUTHOR CONTRIBUTIONS

The study was conceived, designed and supervised by M.R.S., P.B., and L.B.A. Analysis of data was performed by M.D.-G., W.d.S., S. Moody, M.K., A.A., C.D.S, R.V., S. Senkin, J.W., S.F., E.N.B., A.K., B.O., T. Cattiaux, R.C.C.P., V.G., S.C., and J.W.T. Analysis and interpretation of the microbiomics data was performed by A.A. with assistance and advice from L.B.A. and T.D.L. Pathology review was carried out by B.A.-A. Sample manipulation was carried out by P.C., C.C., and C.L. Patient and sample recruitment was led or facilitated by D.Z., R.C., M.A., L.P., S.G., J.Y., R.M., A.N., M.M., K.E., S. Milosavljevic, S. Sangrajrang, M.P.C., S.A., R.M.R., M.T.R., L.G.R., D.P.G., I.H., J.K., C.A.V., T.A.P., B.Ś., J.L., K.R.P., A.H.-S., T.S., S. Shiba, S. Sangkhathat, T. Chitapanarux, G.R., P.A.-P., D.C.D., and F.H.d.O. Scientific project management was carried out by L.H., A.C.d.C., and S.P. M.D.-G., W.d.S., and S. Moody jointly contributed and were responsible for overall scientific coordination. The manuscript was written by M.D.-G., W.d.S., S. Moody, M.R.S., P.B., and L.B.A., with contributions from all other authors. All authors read and approved the final manuscript.

