## Extended Data Figures 1 - 10 for "Geographic and age-related variations in mutational processes in colorectal cancer"

### Extended Data Figure 1

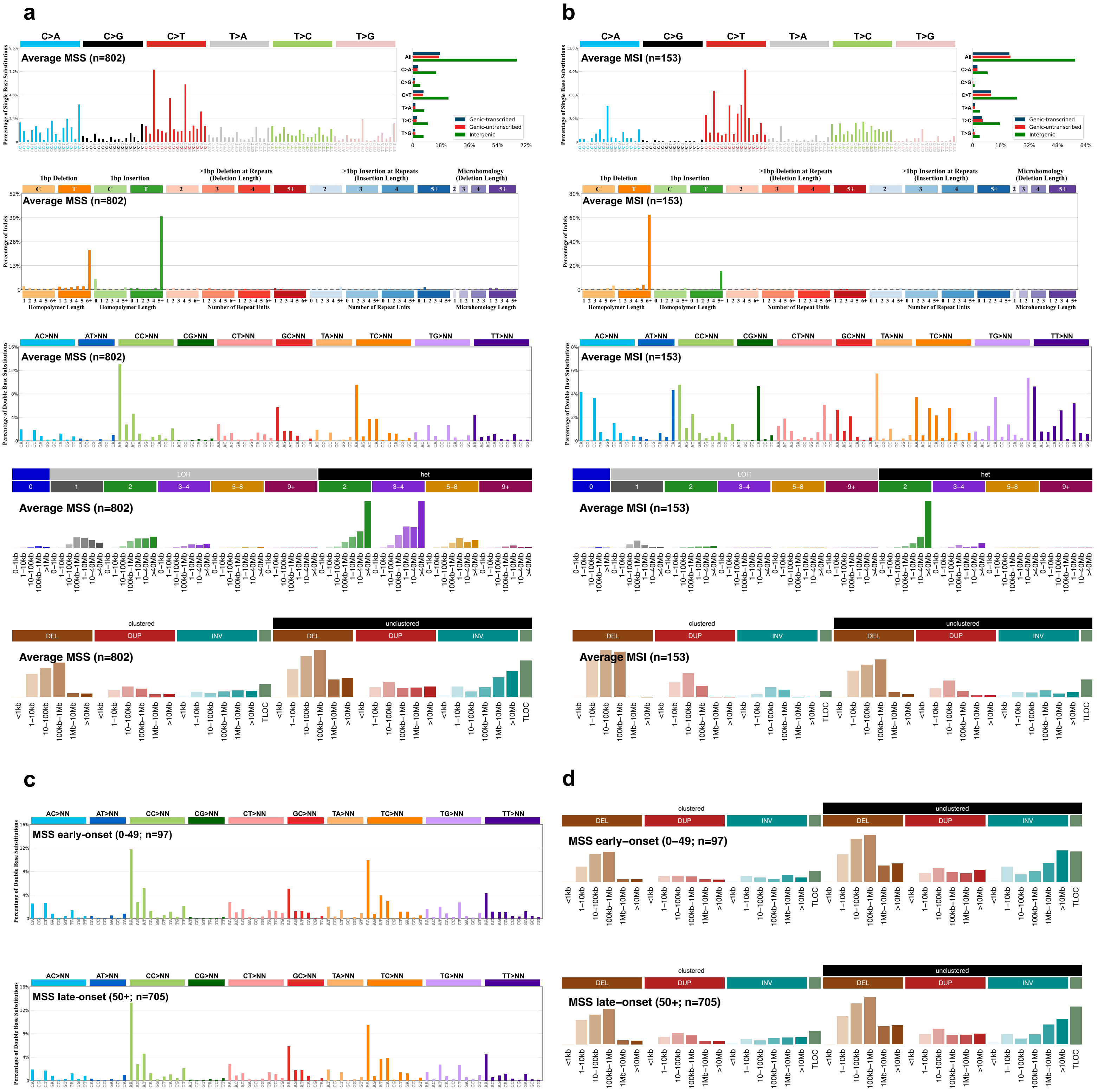

### Extended Data Figure 2

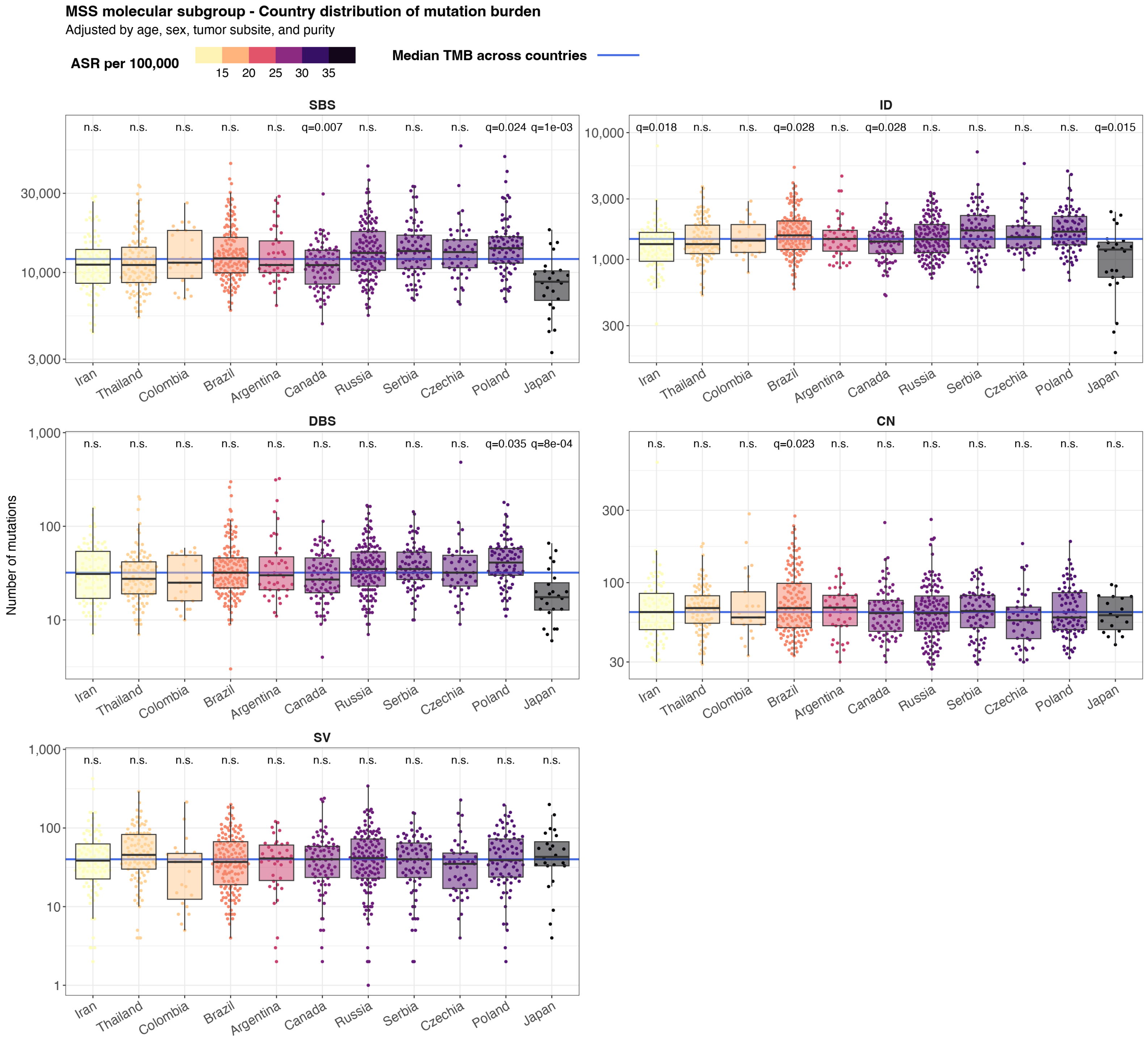

### Extended Data Figure 3

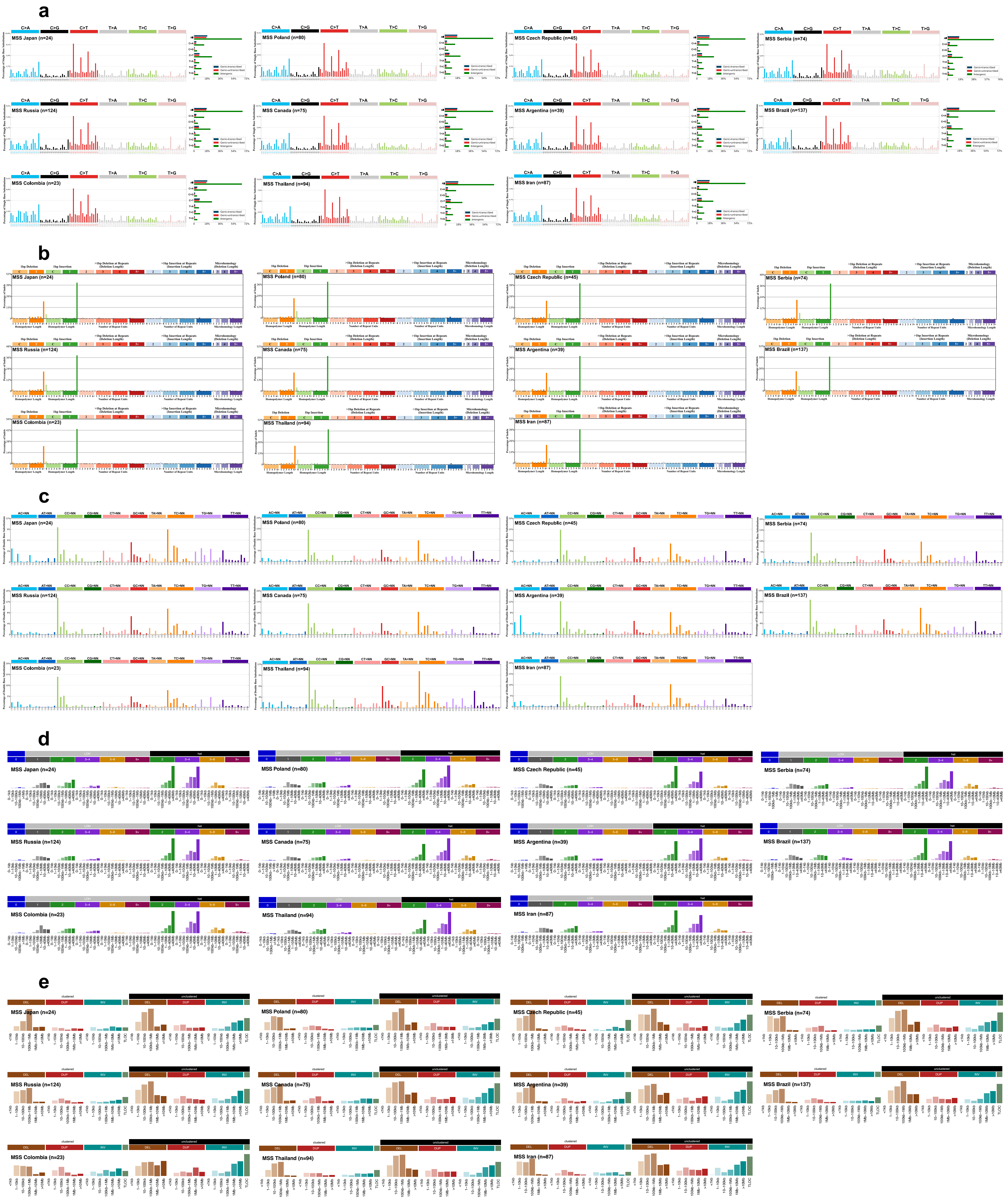

### Extended Data Figure 4

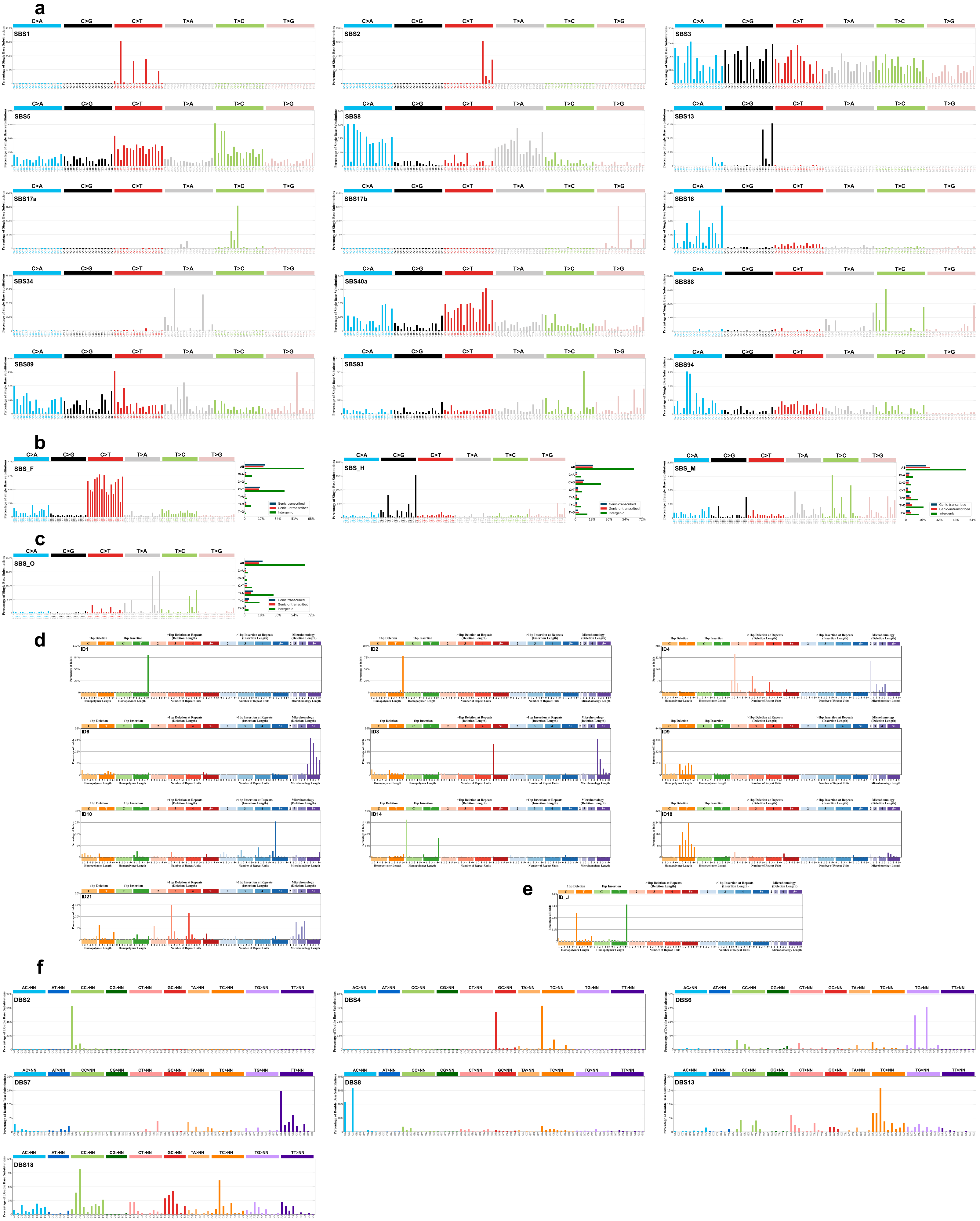

### Extended Data Figure 5

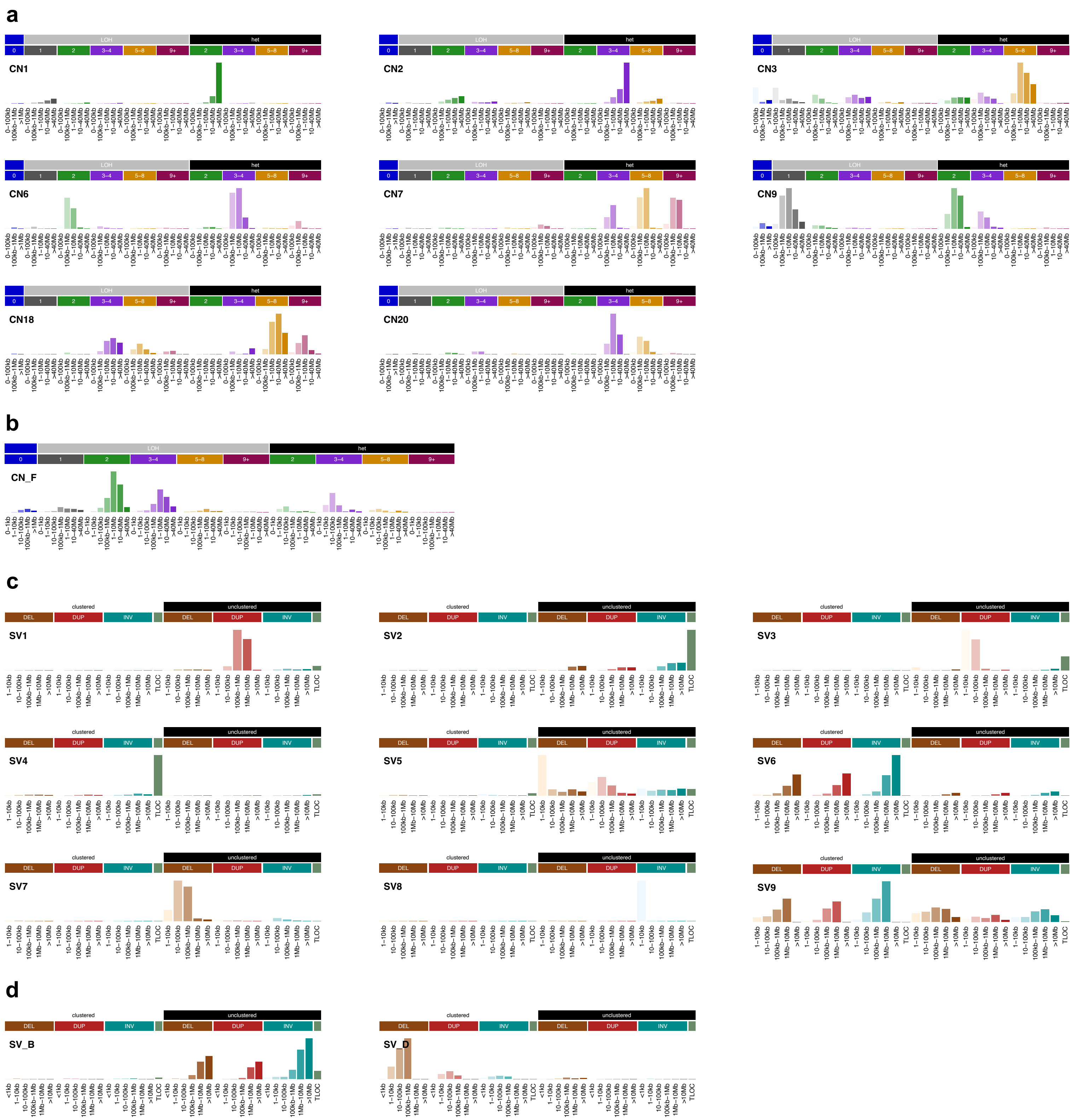

### Extended Data Figure 6

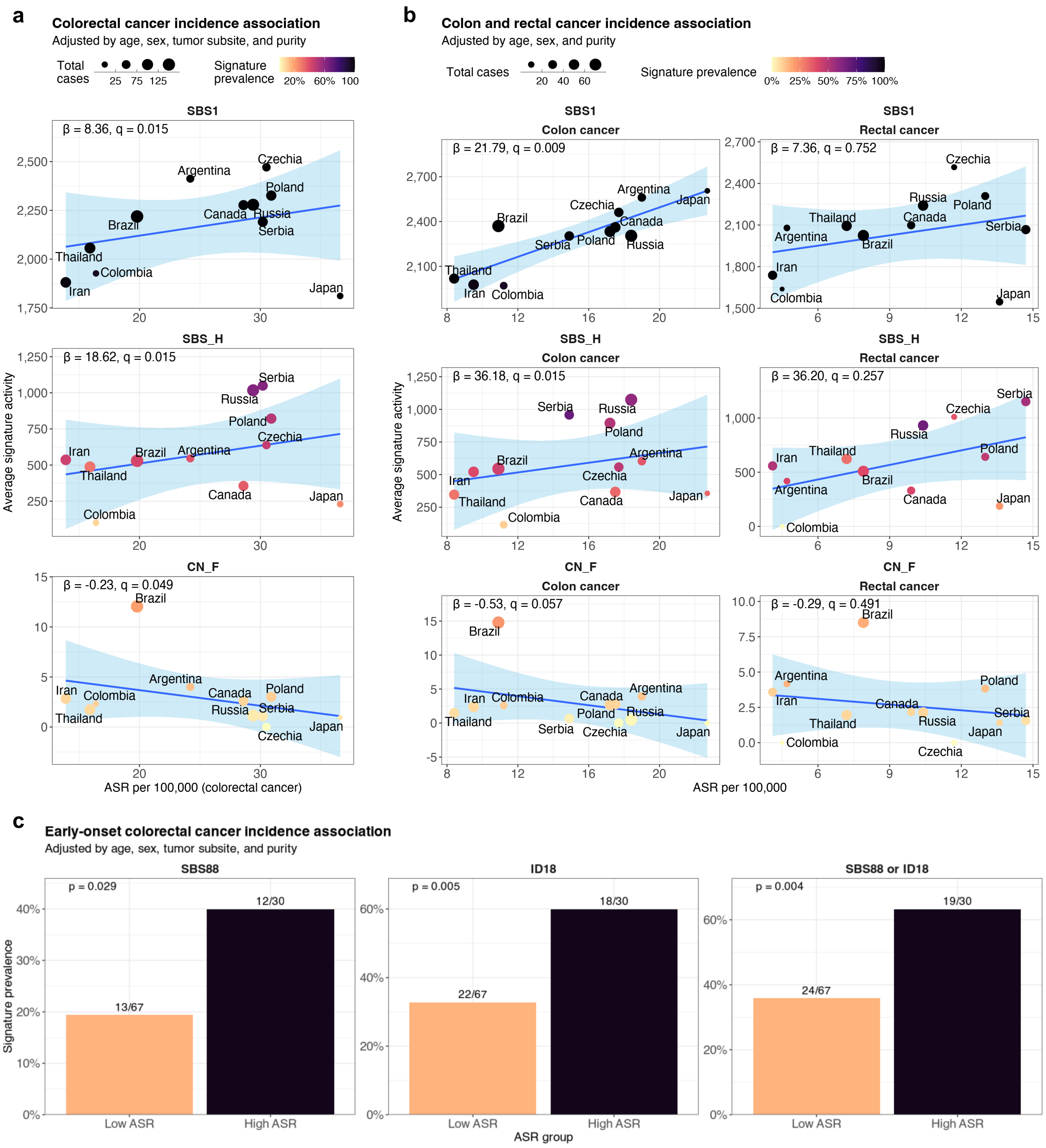

### Extended Data Figure 7

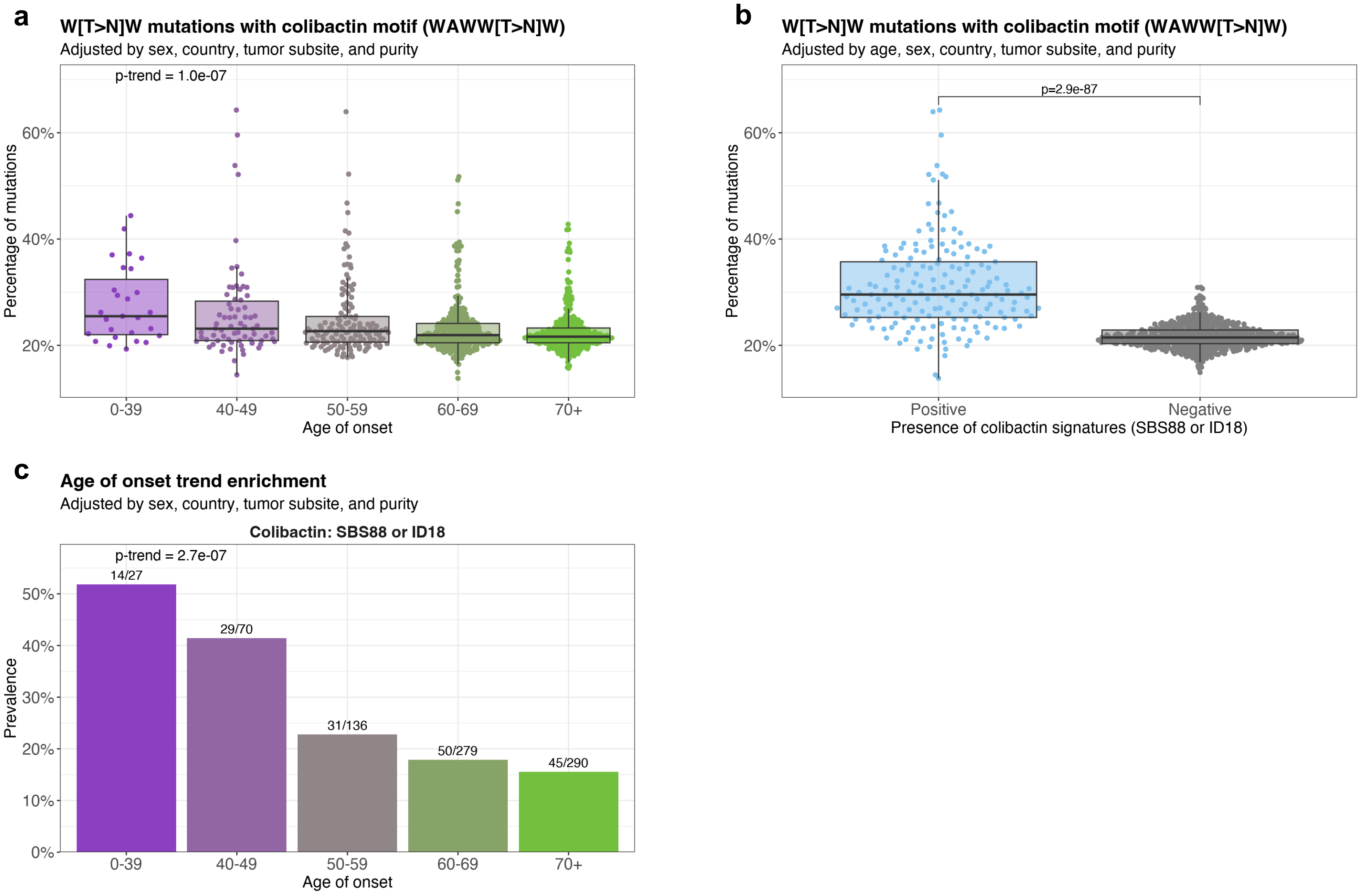

### Extended Data Figure 8

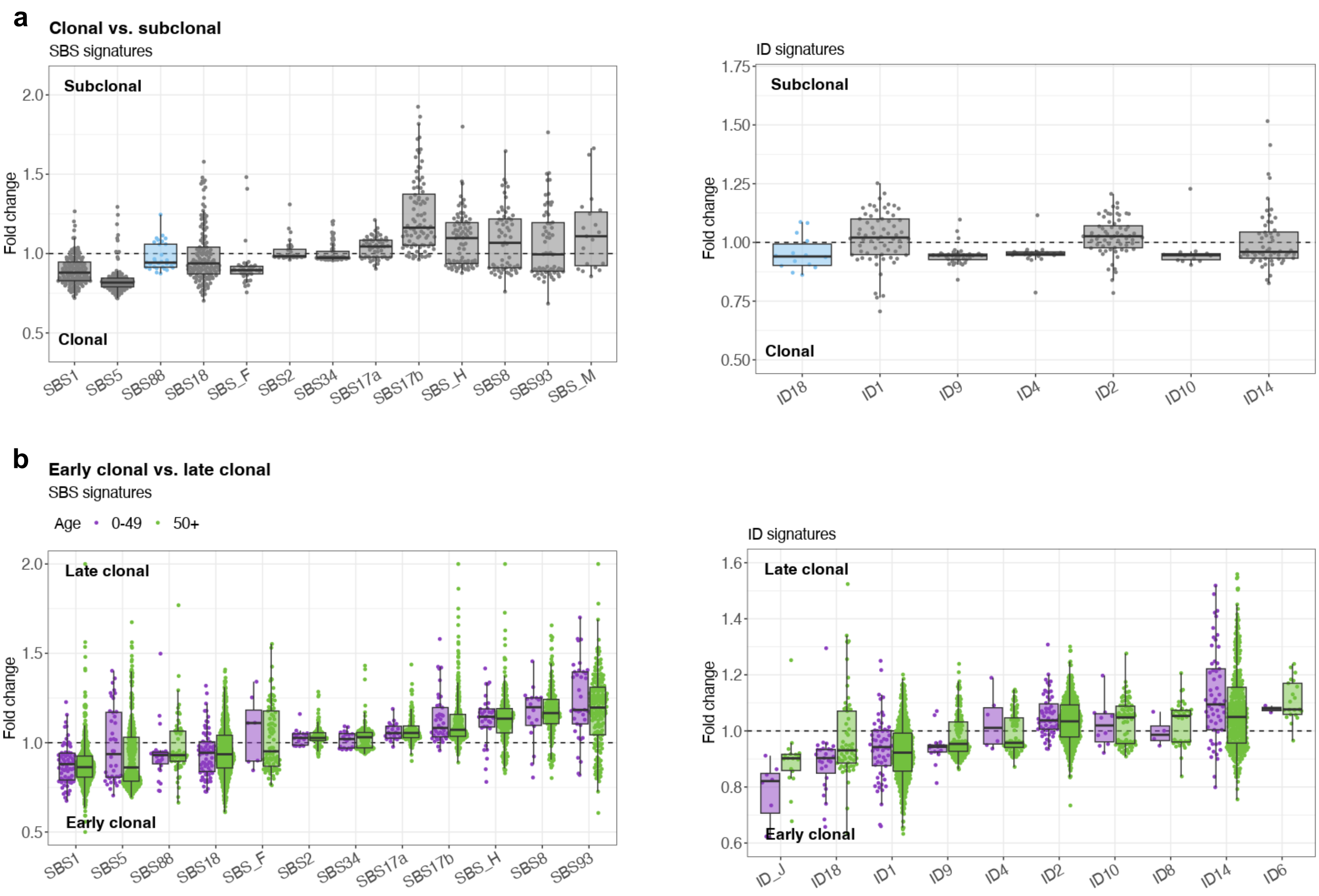

#### Extended Data Figure 9

**a**

Genomic+ and *pks*+

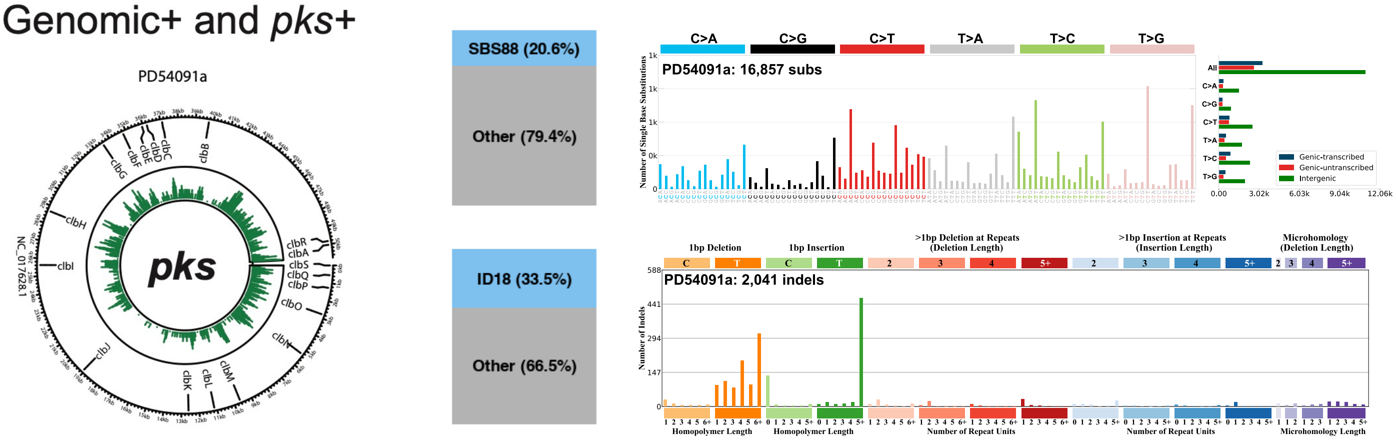


**b**

Genomic+ and *pks*-

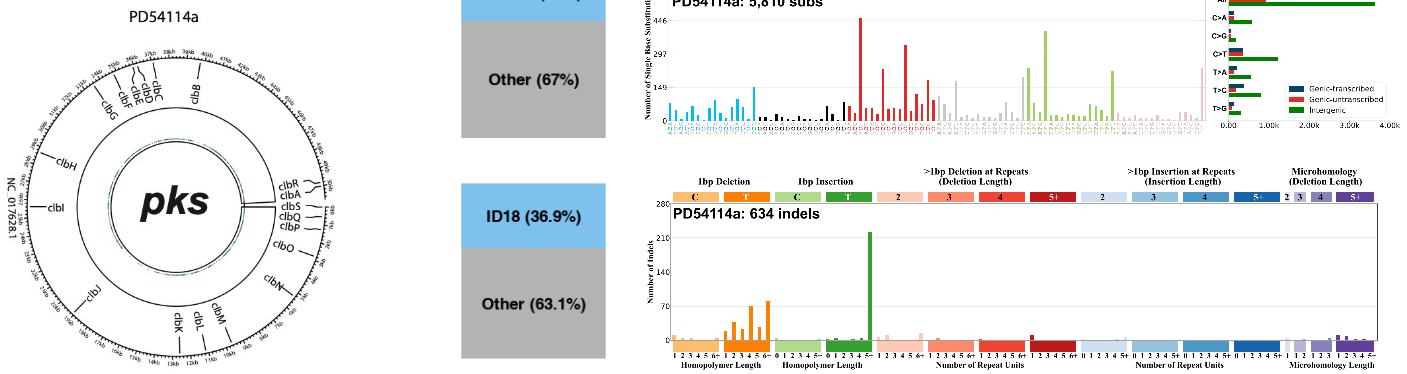


**c**

Genomic- and *pks*+

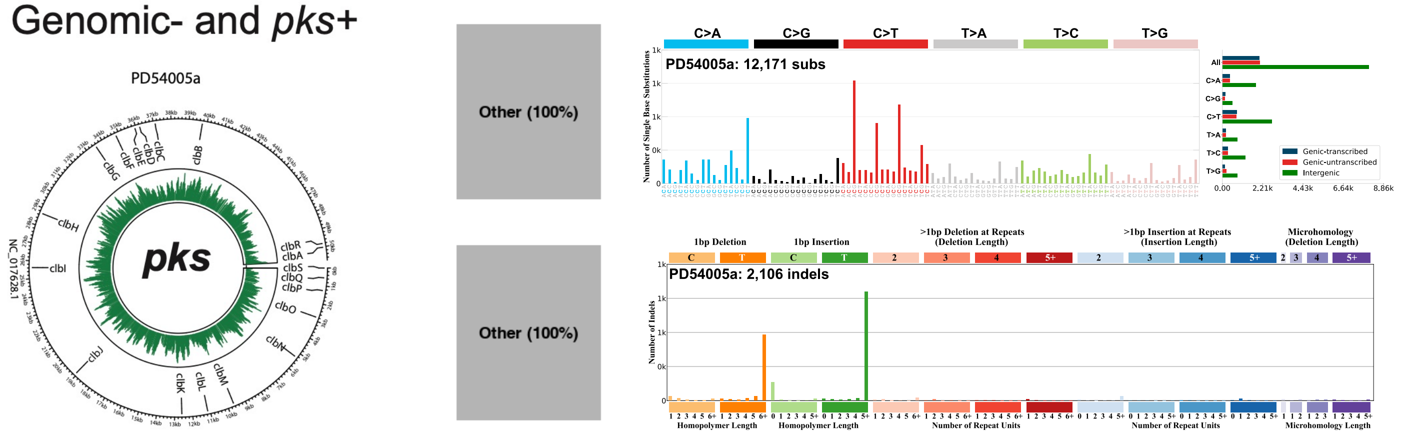


**d**

Genomic- and *pks*-

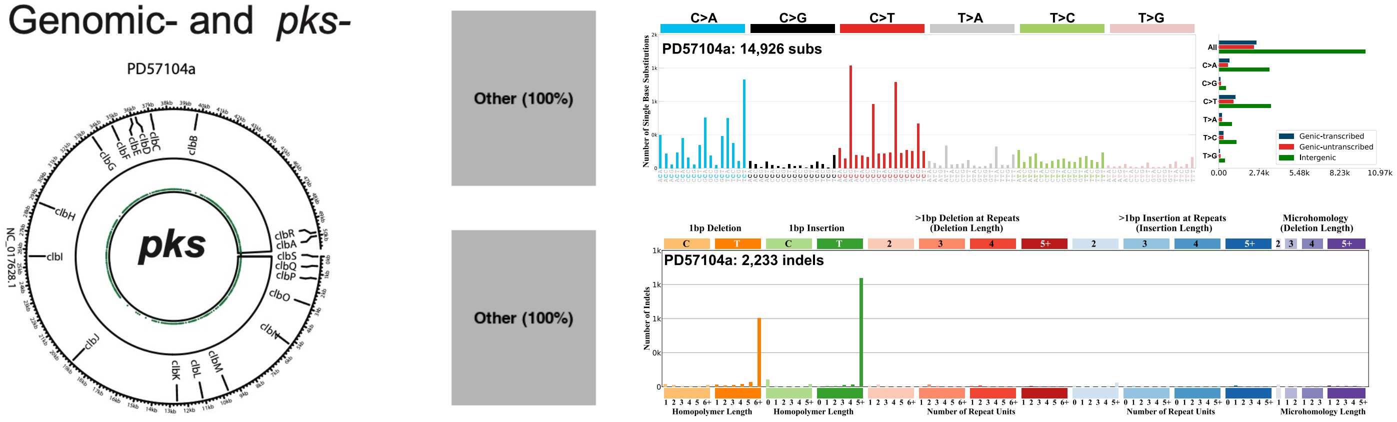

#### Extended Data Figure 10

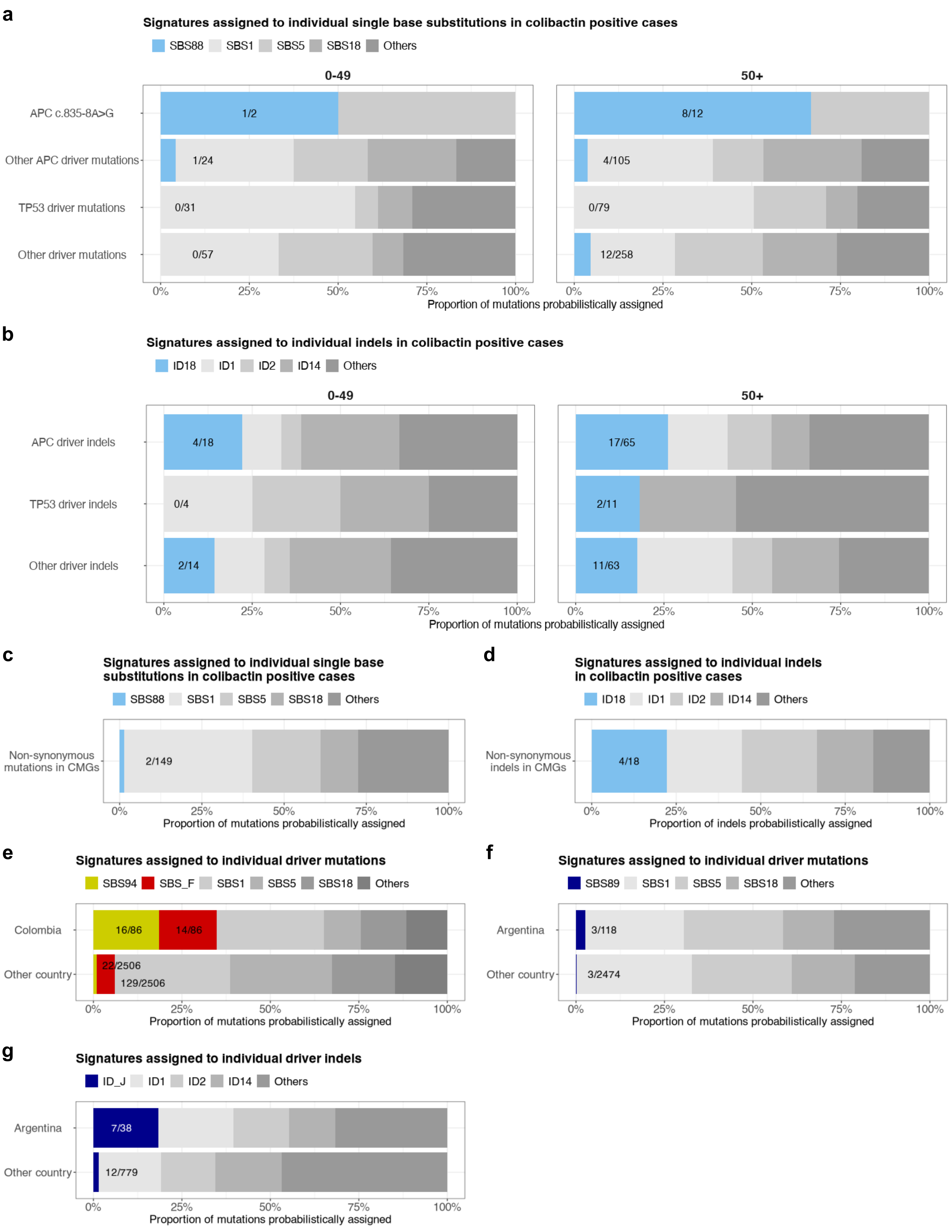
