## Supplementary Figures 1 - 19 for "Geographic and age-related variations in mutational processes in colorectal cancer"

### Supplementary Figure 1

**a**

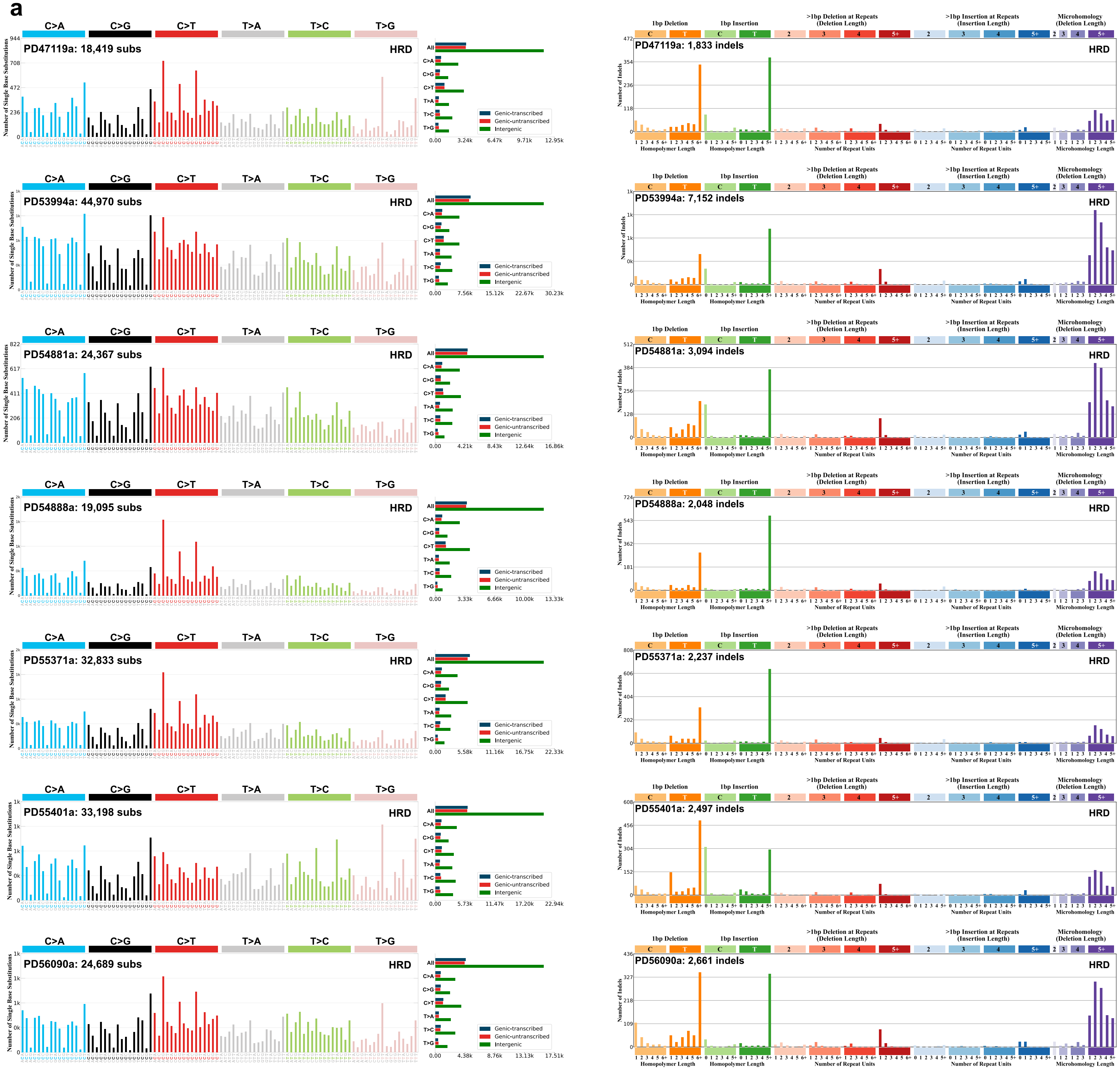


**b**

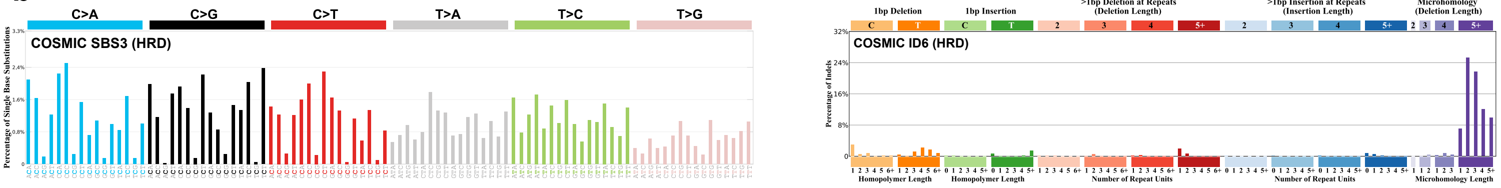

### Supplementary Figure 2

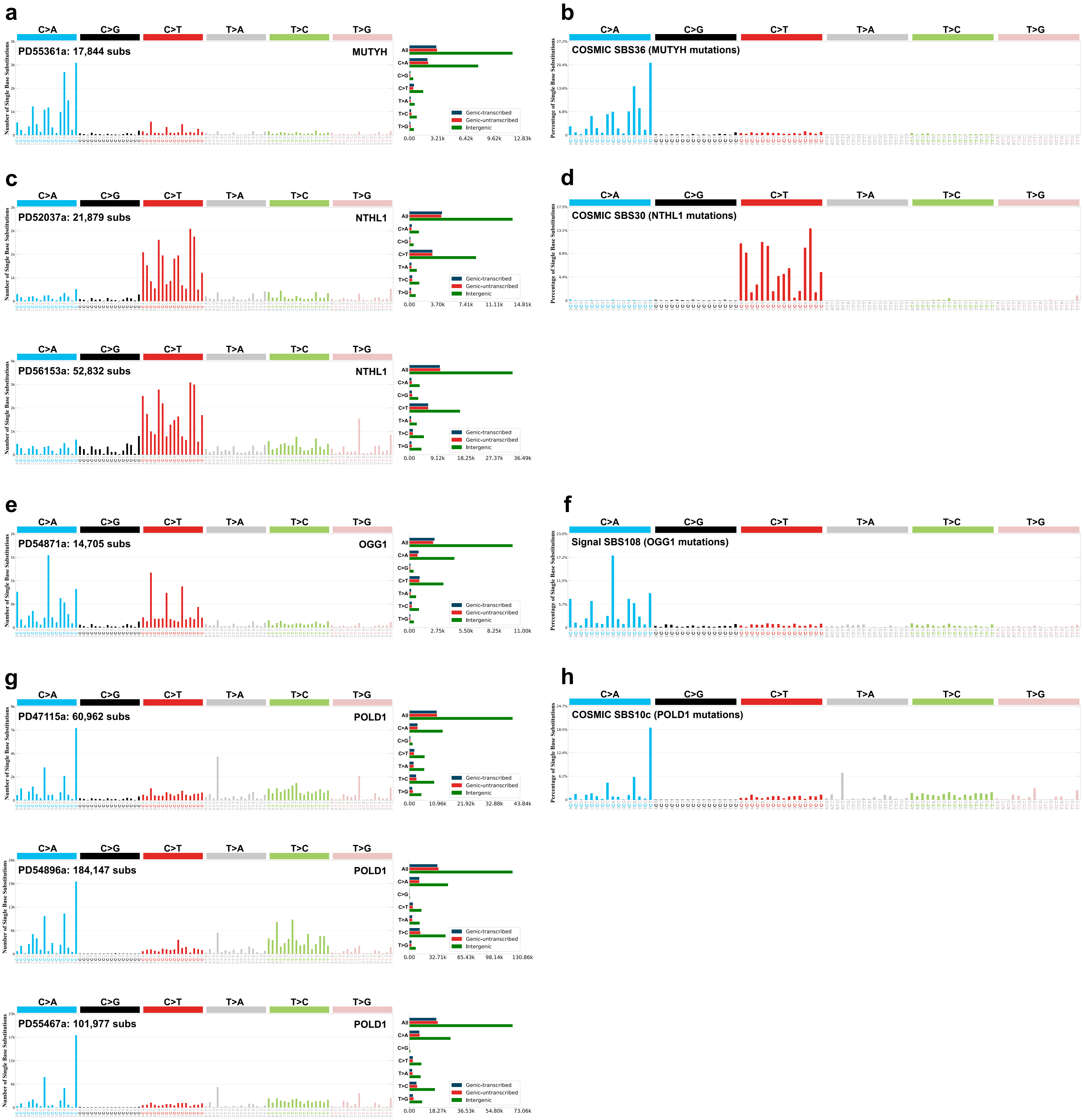

### Supplementary Figure 3

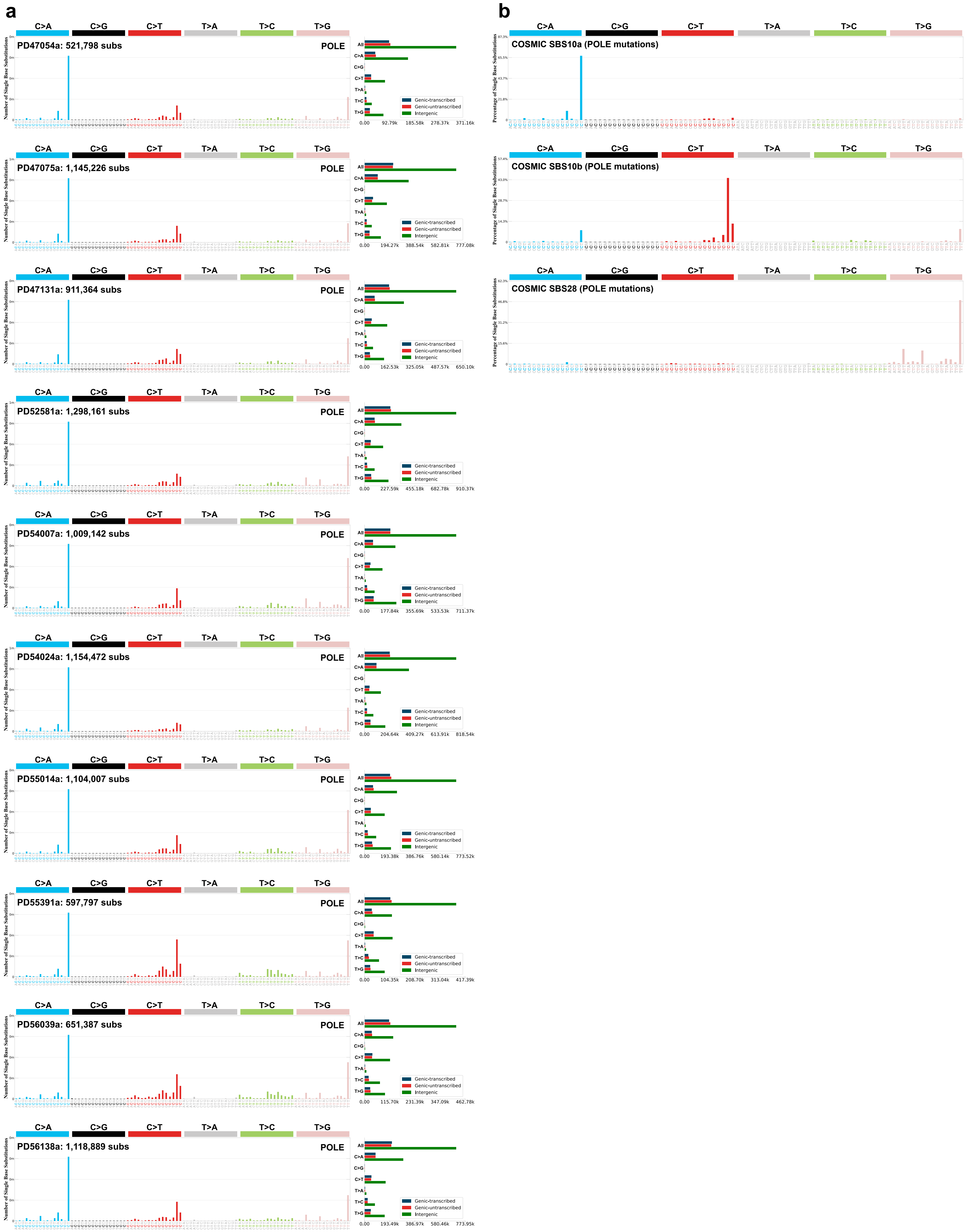

### Supplementary Figure 4

**a**

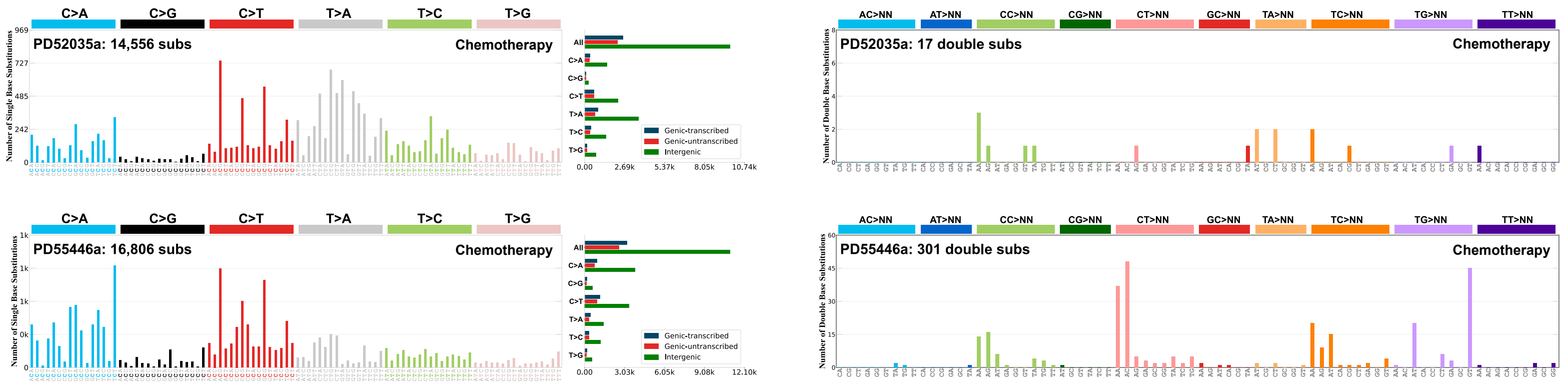


**b**

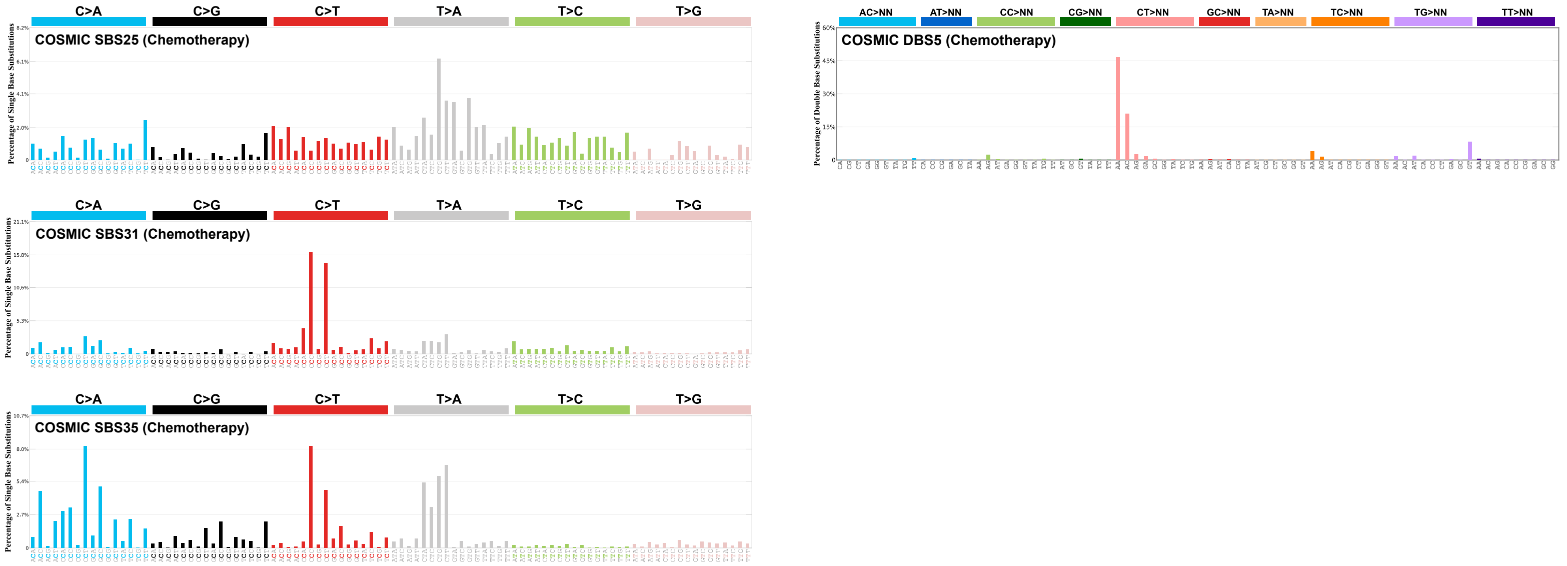

### Supplementary Figure 5

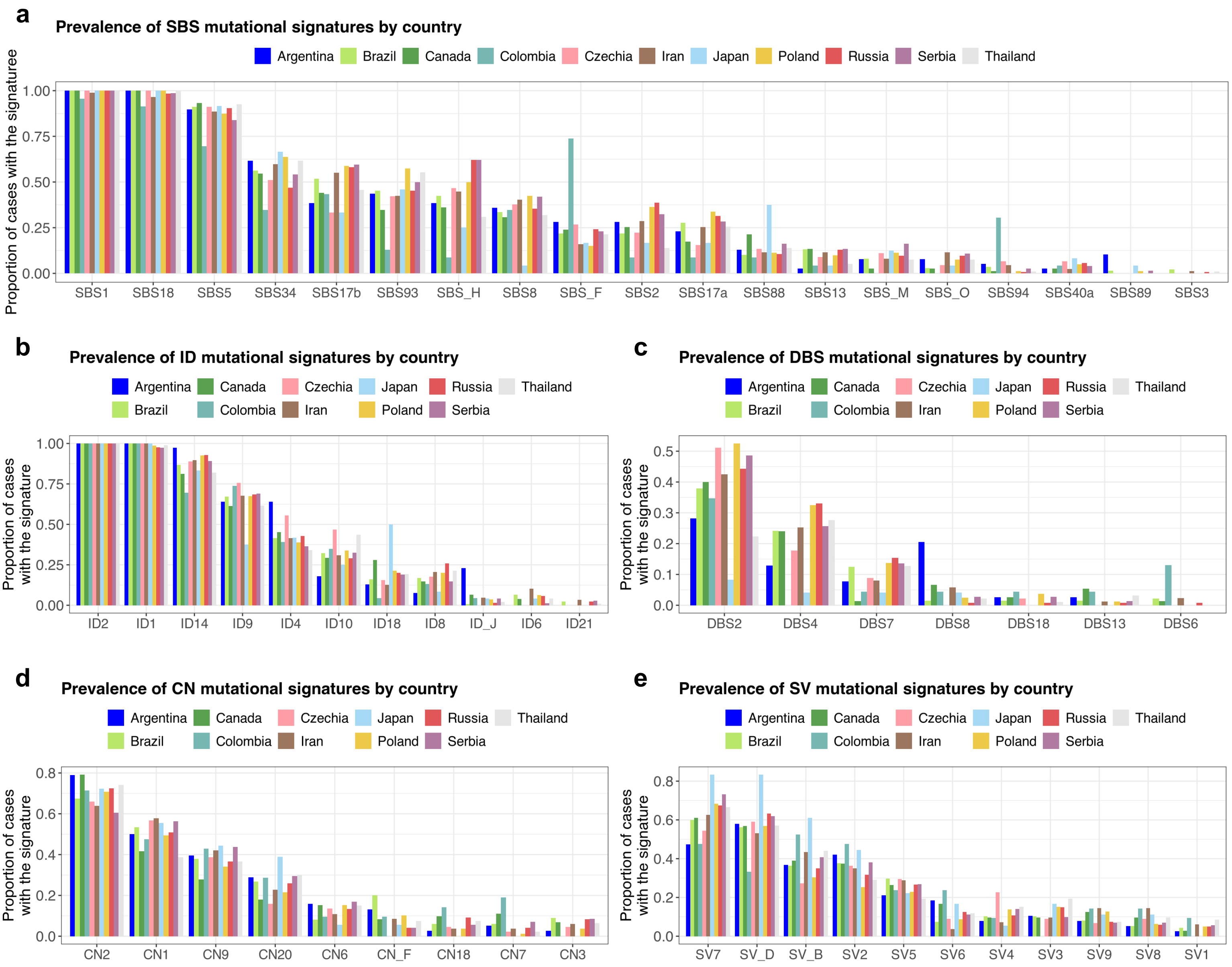

#### Supplementary Figure 6

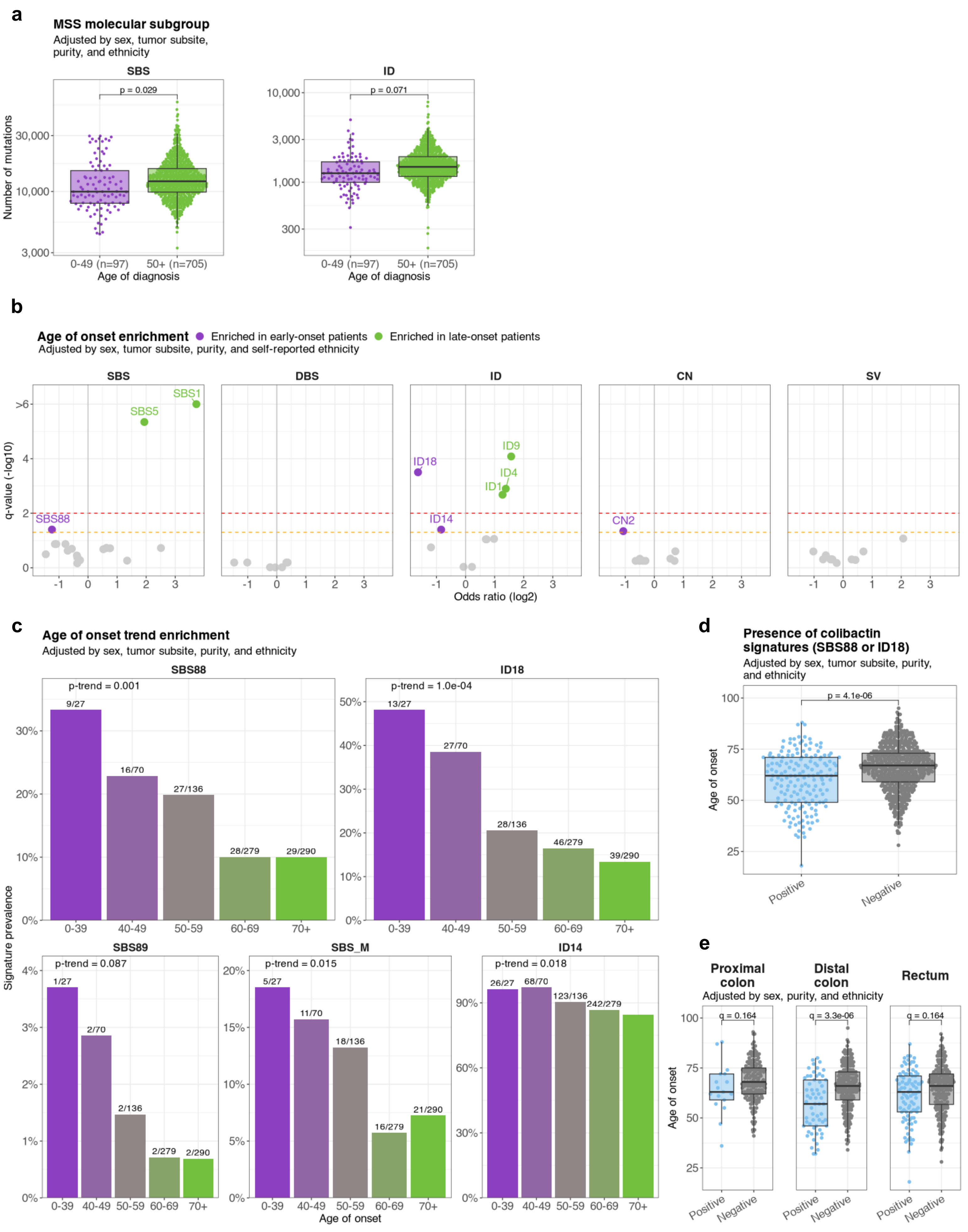

#### Supplementary Figure 7

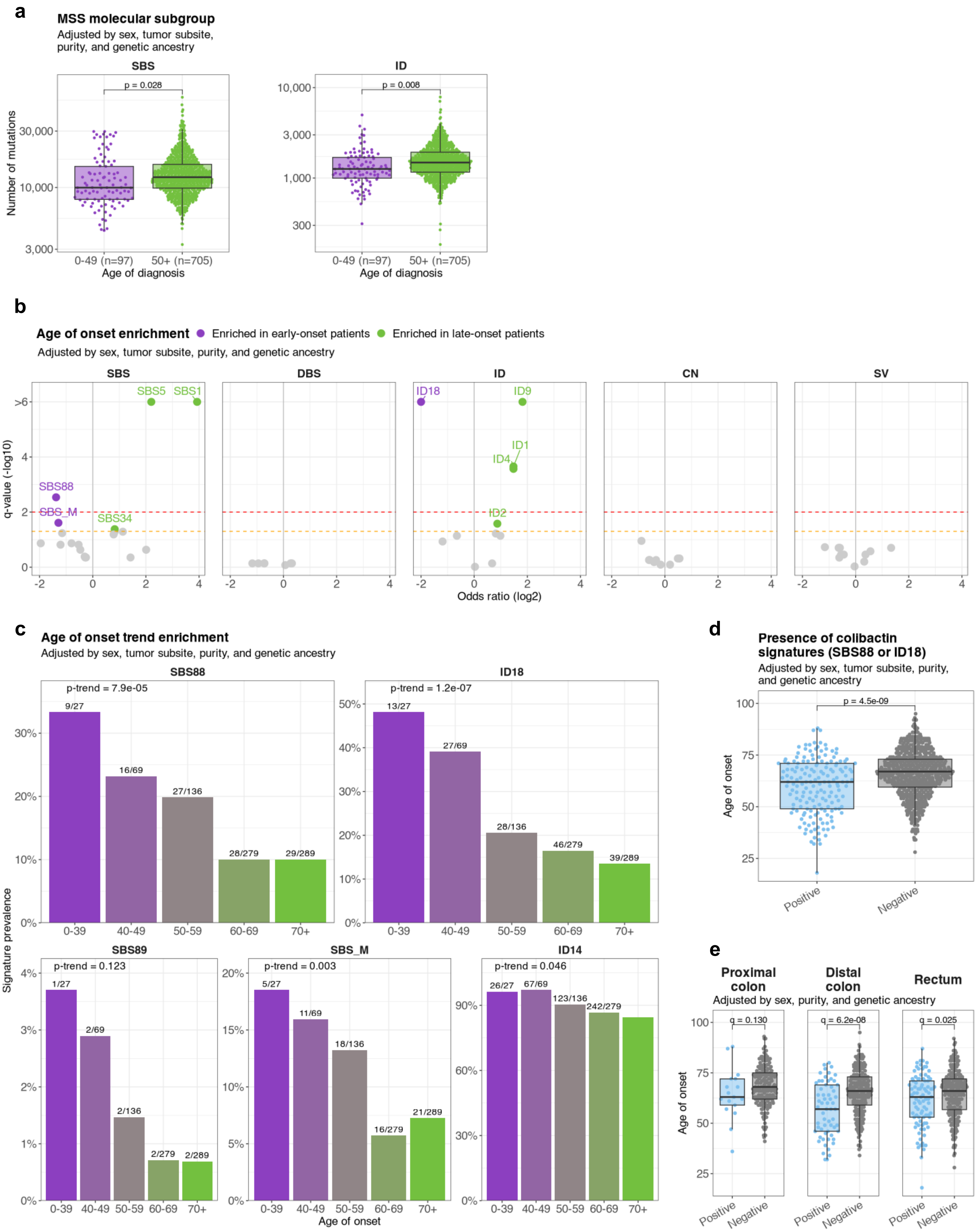

#### Supplementary Figure 8

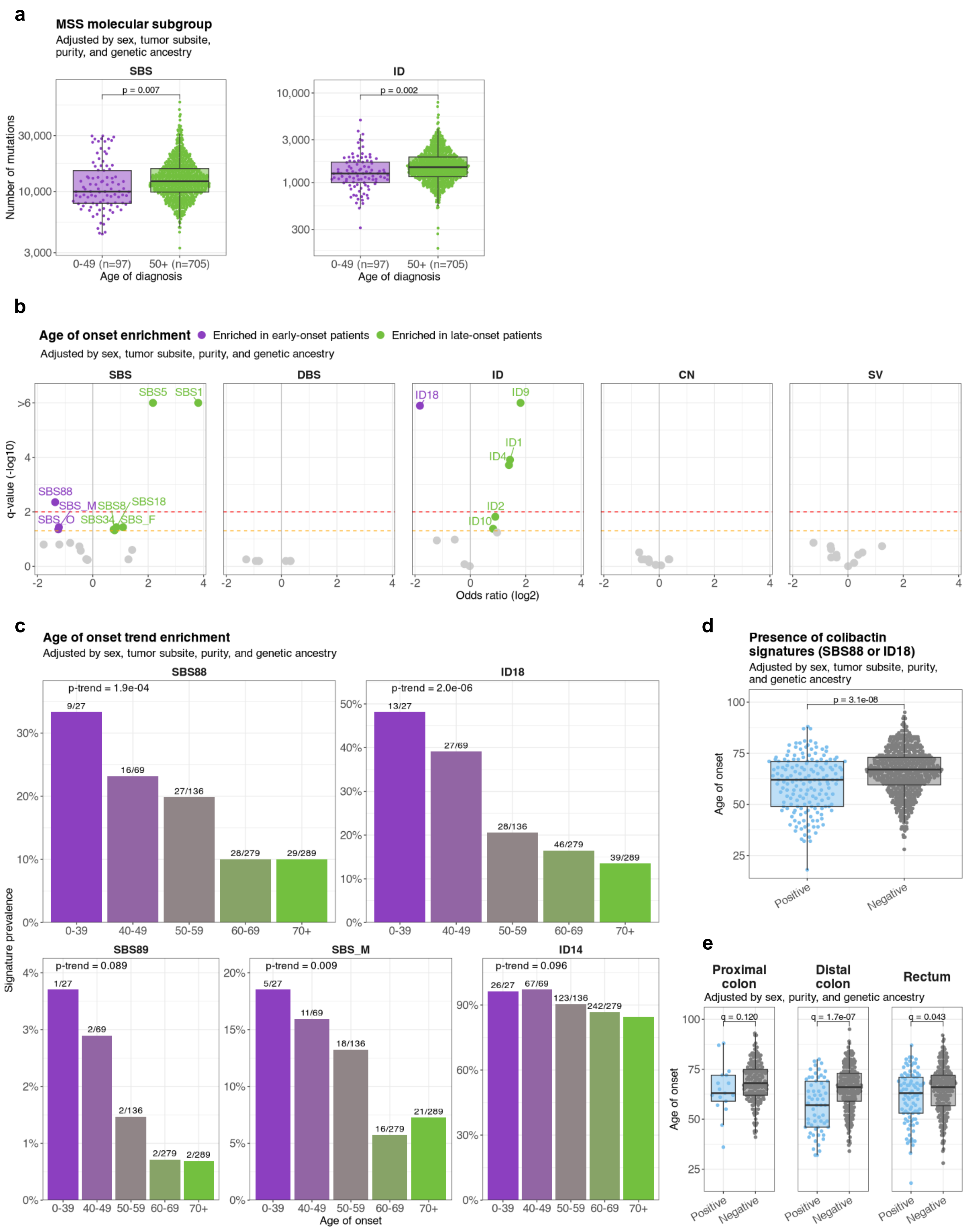

#### Supplementary Figure 9

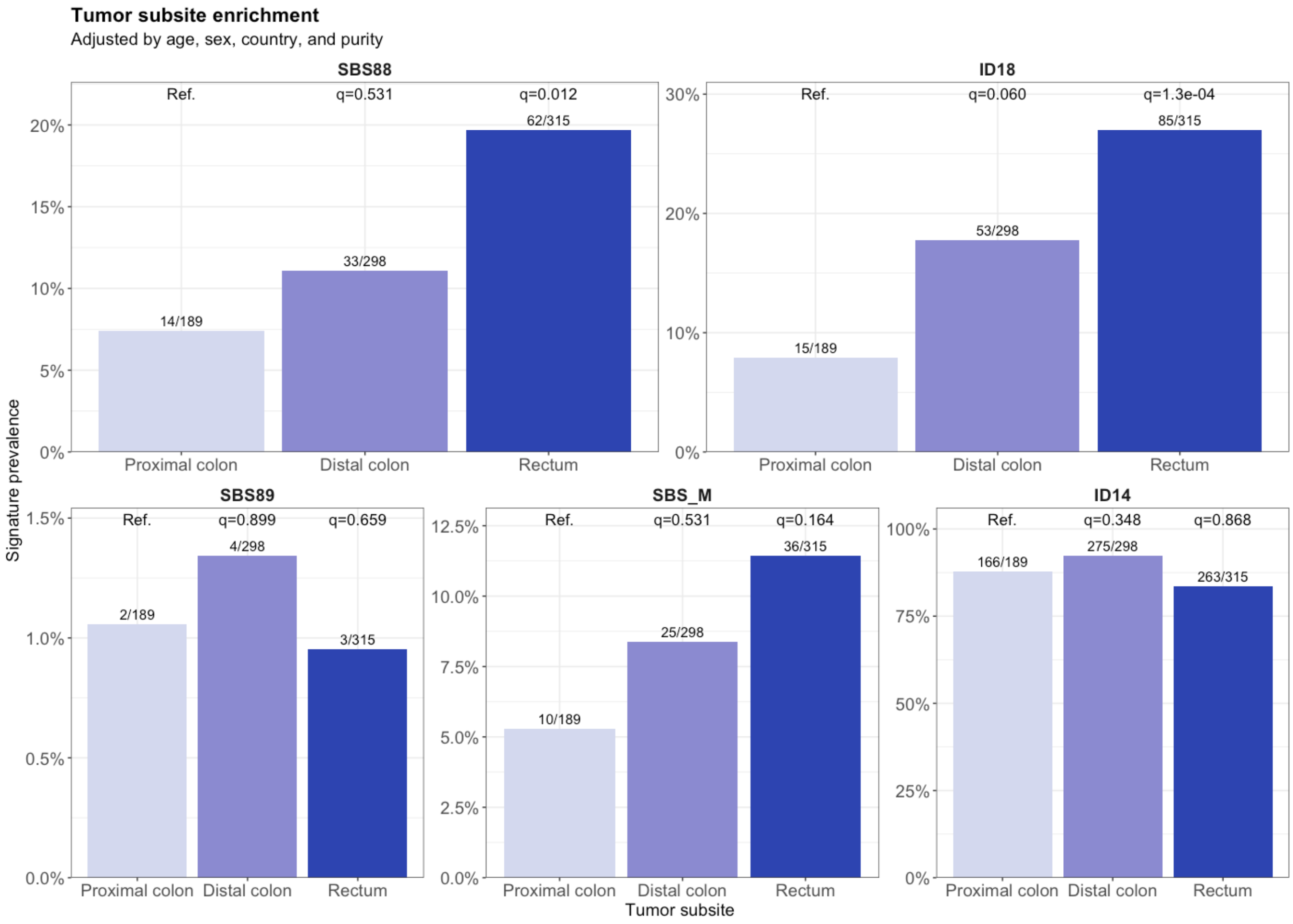

**Supplementary Figure 10**

**a**

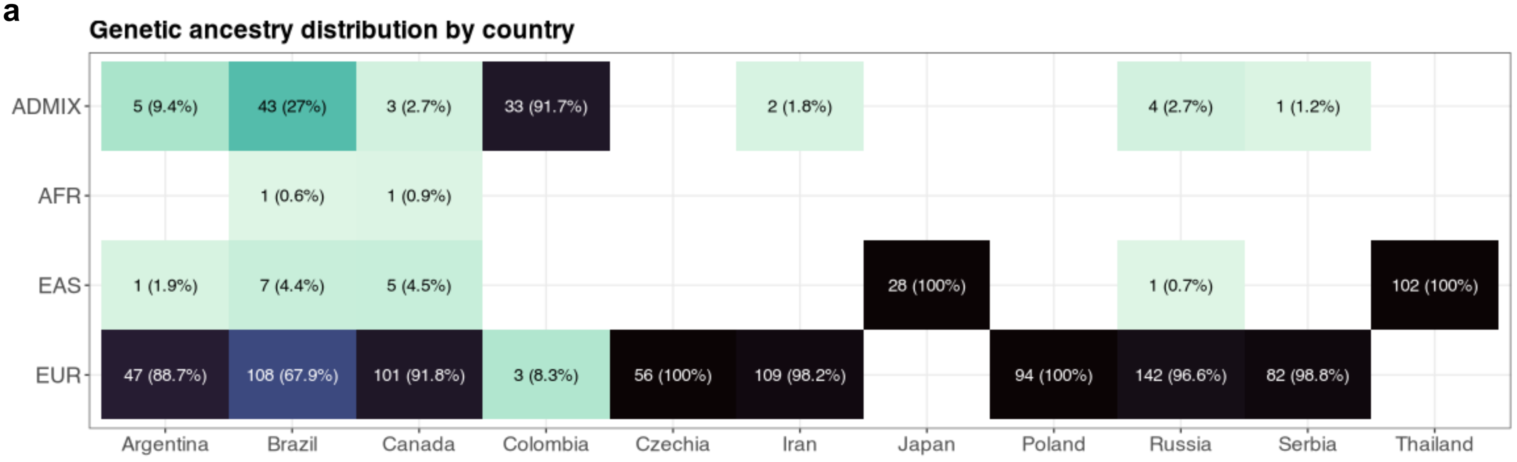


**b**

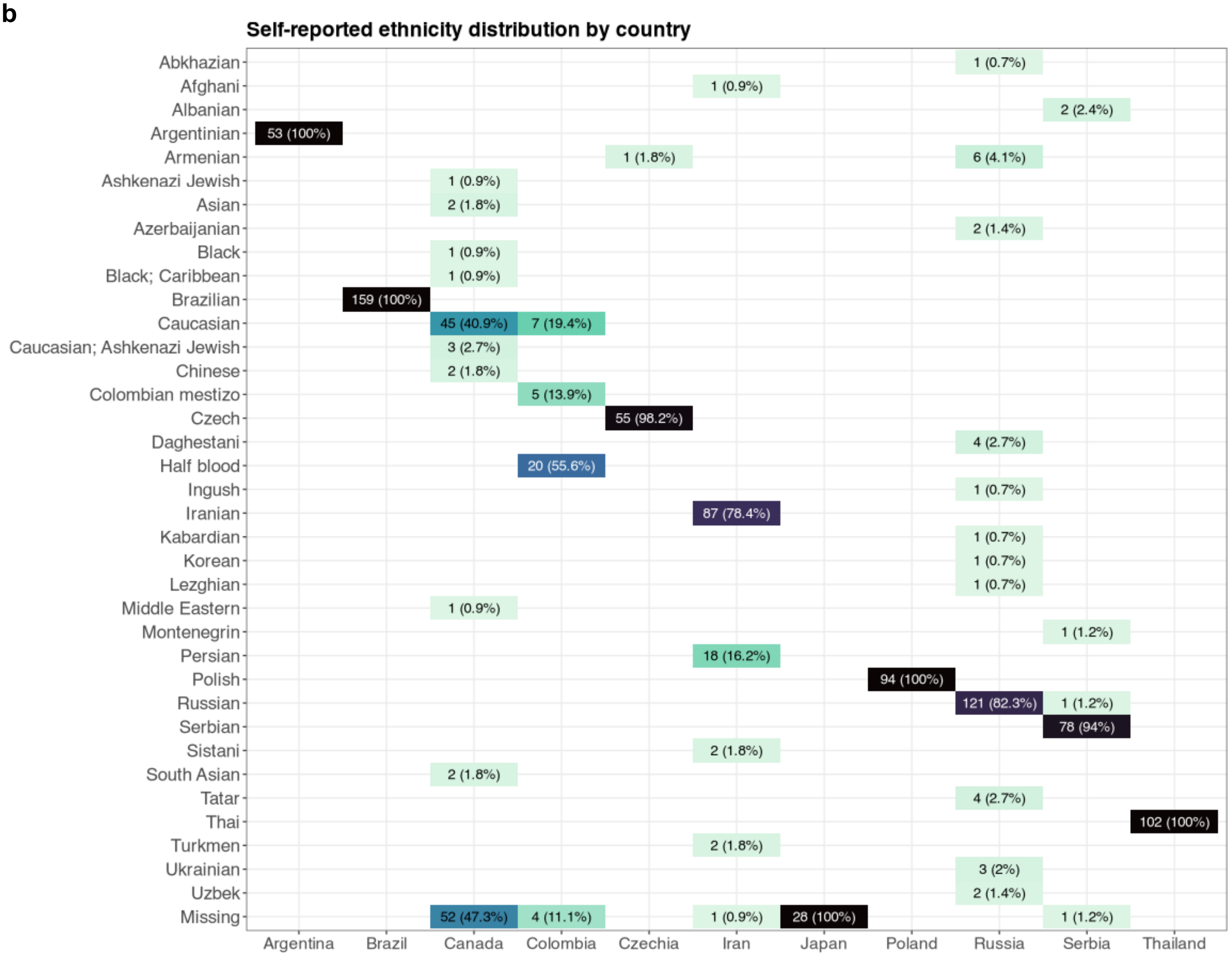

#### Supplementary Figure 11

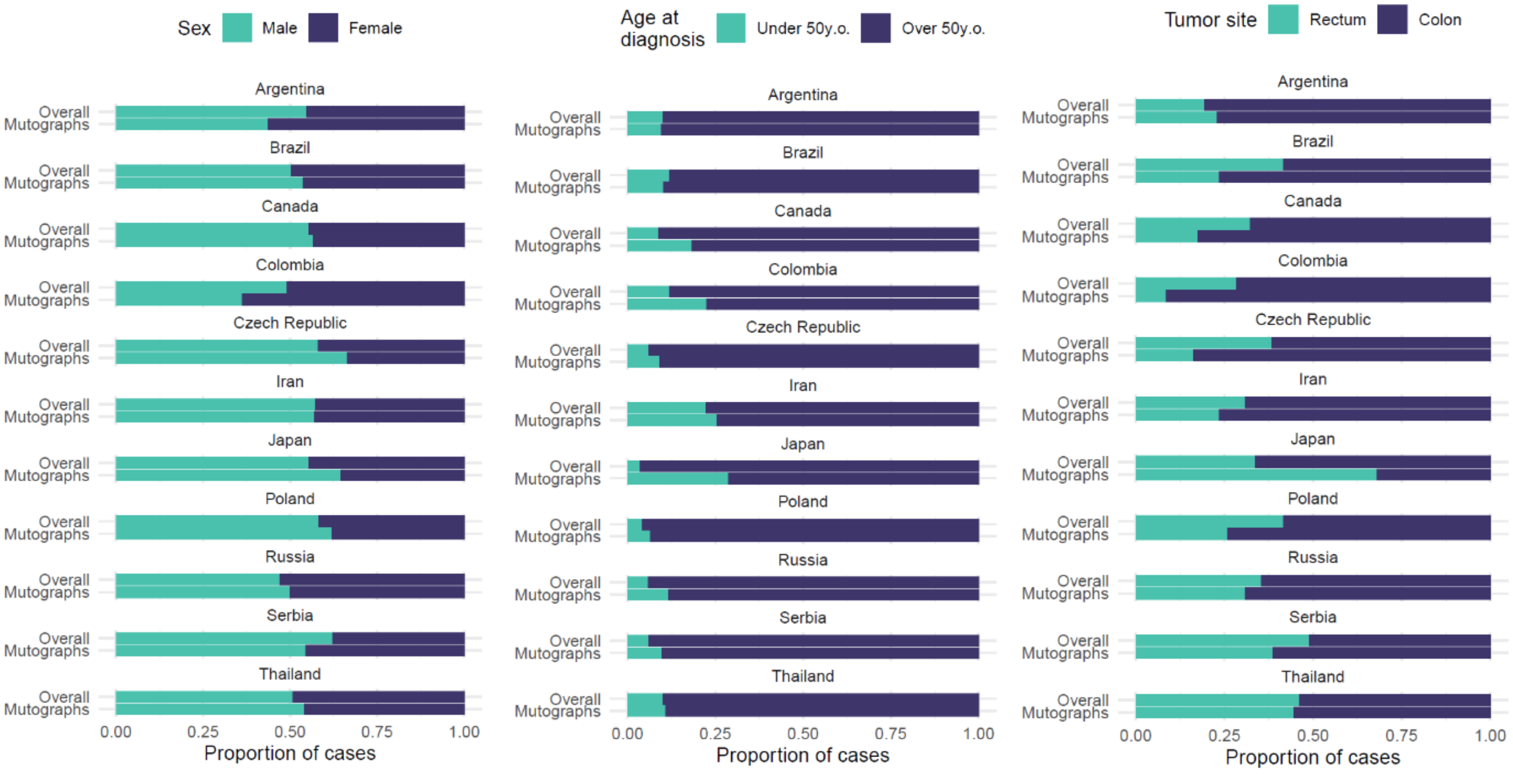

### Supplementary Figure 12

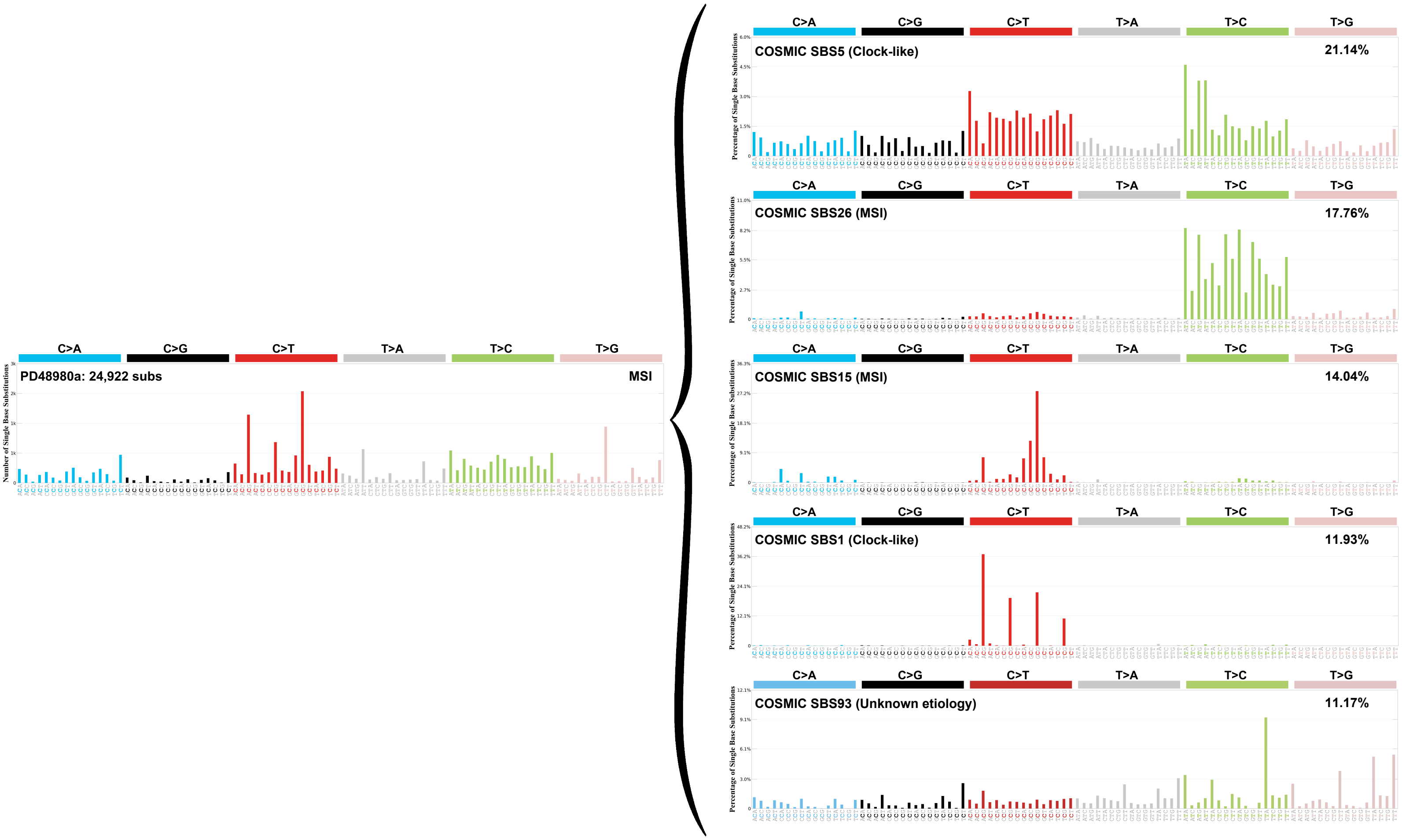


* COSMIC SBS18 (ROS; 9.99%), COSMIC SBS17b (Unknown etiology; 7.21%),
COSMIC SBS34 (Unknown etiology; 6.09%), Unknown (0.66%)

### Supplementary Figure 13

**a**

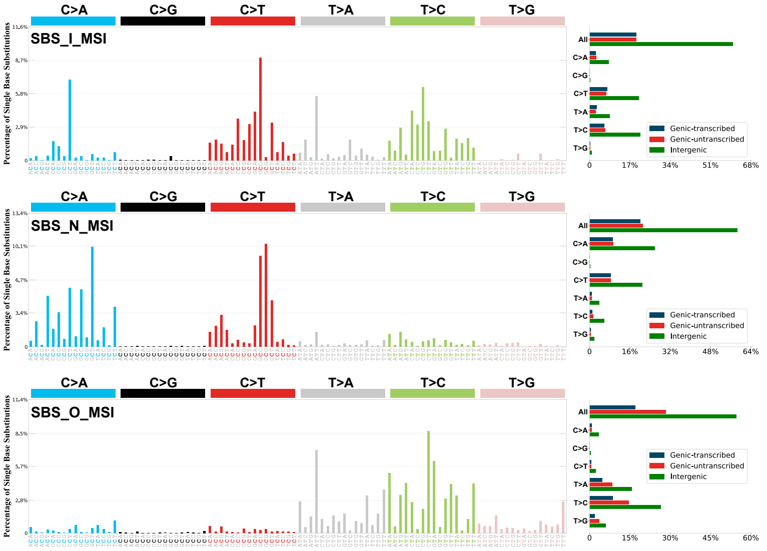


**b**

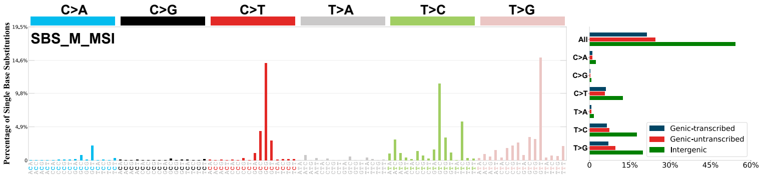


**c**

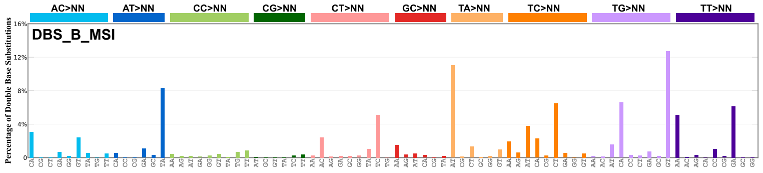


**d**

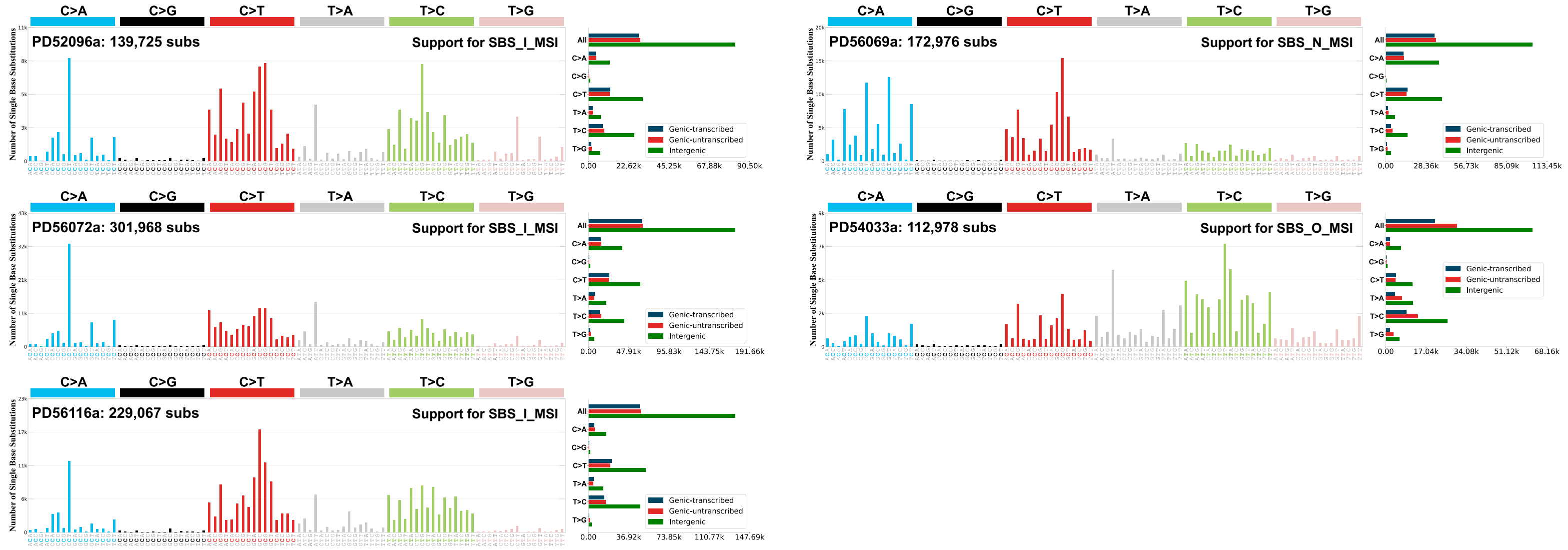


**e**

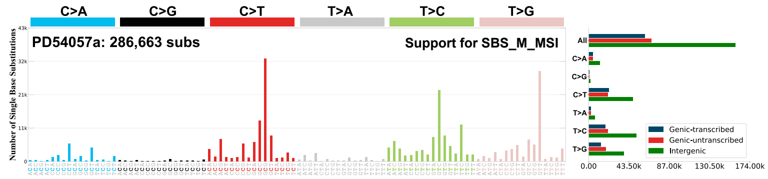

#### Supplementary Figure 14

**a**

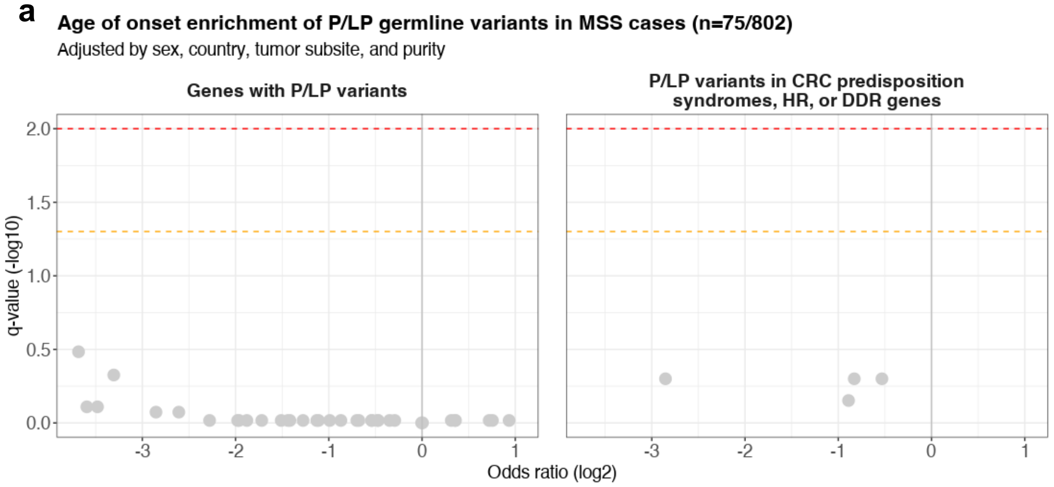


**b**

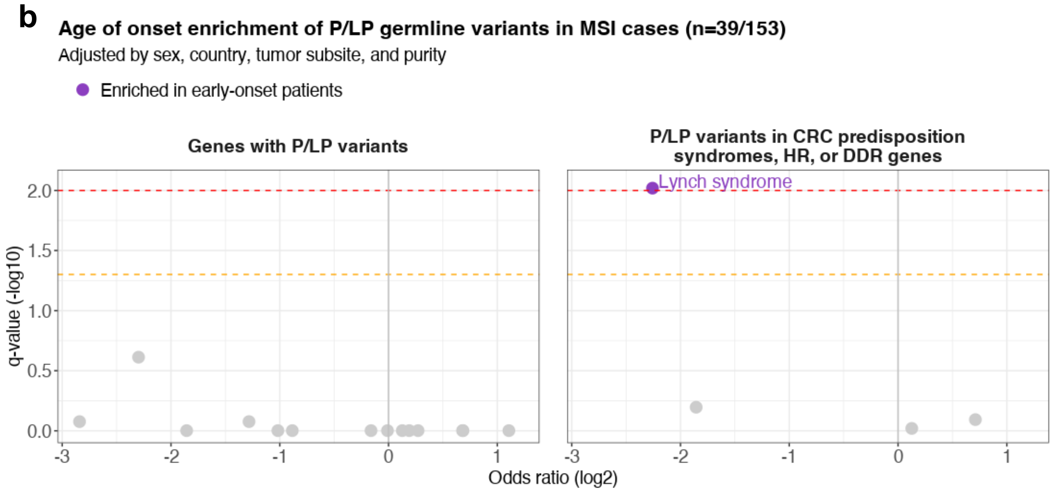

### Supplementary Figure 15

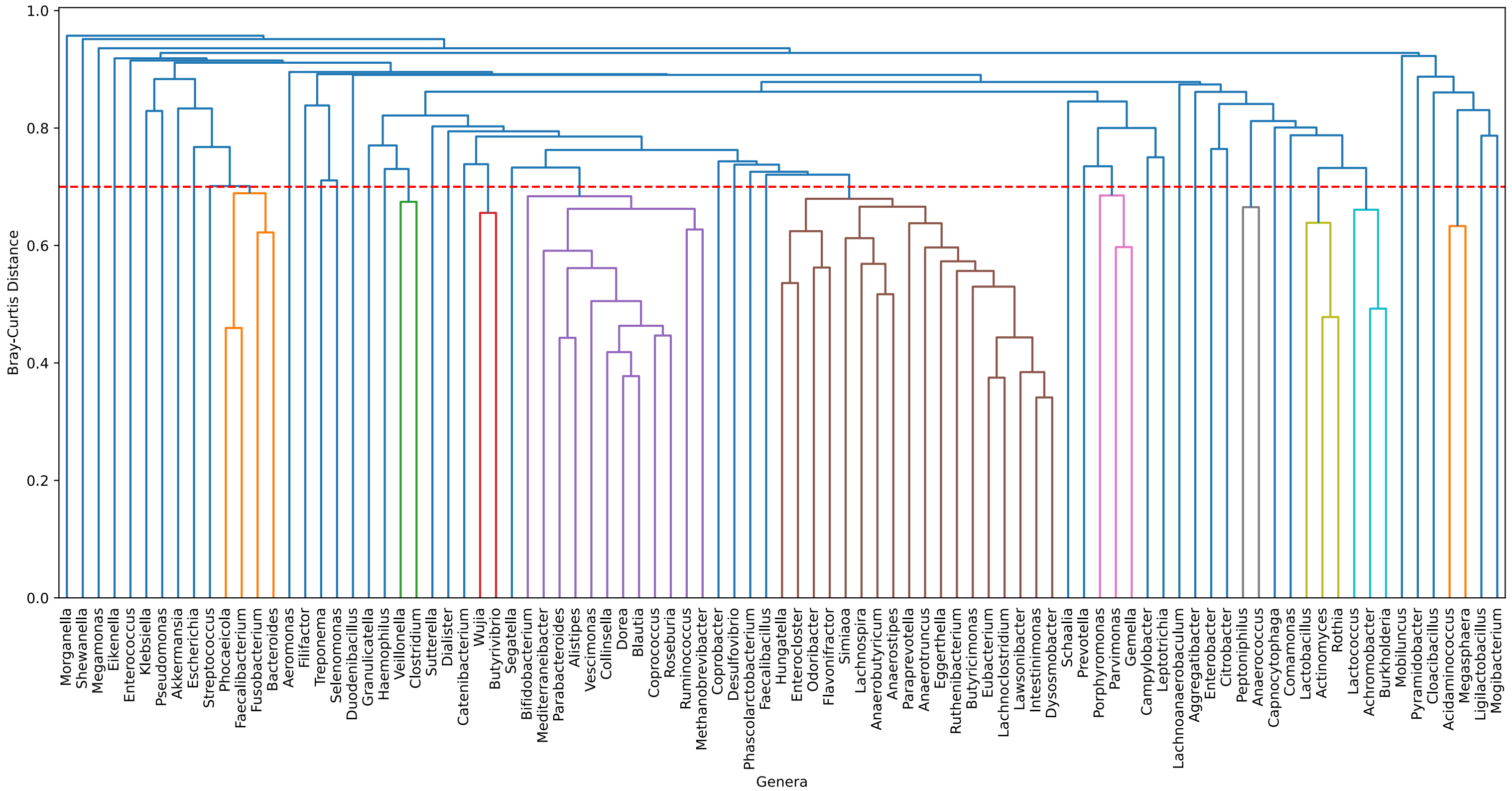

### Supplementary Figure 16

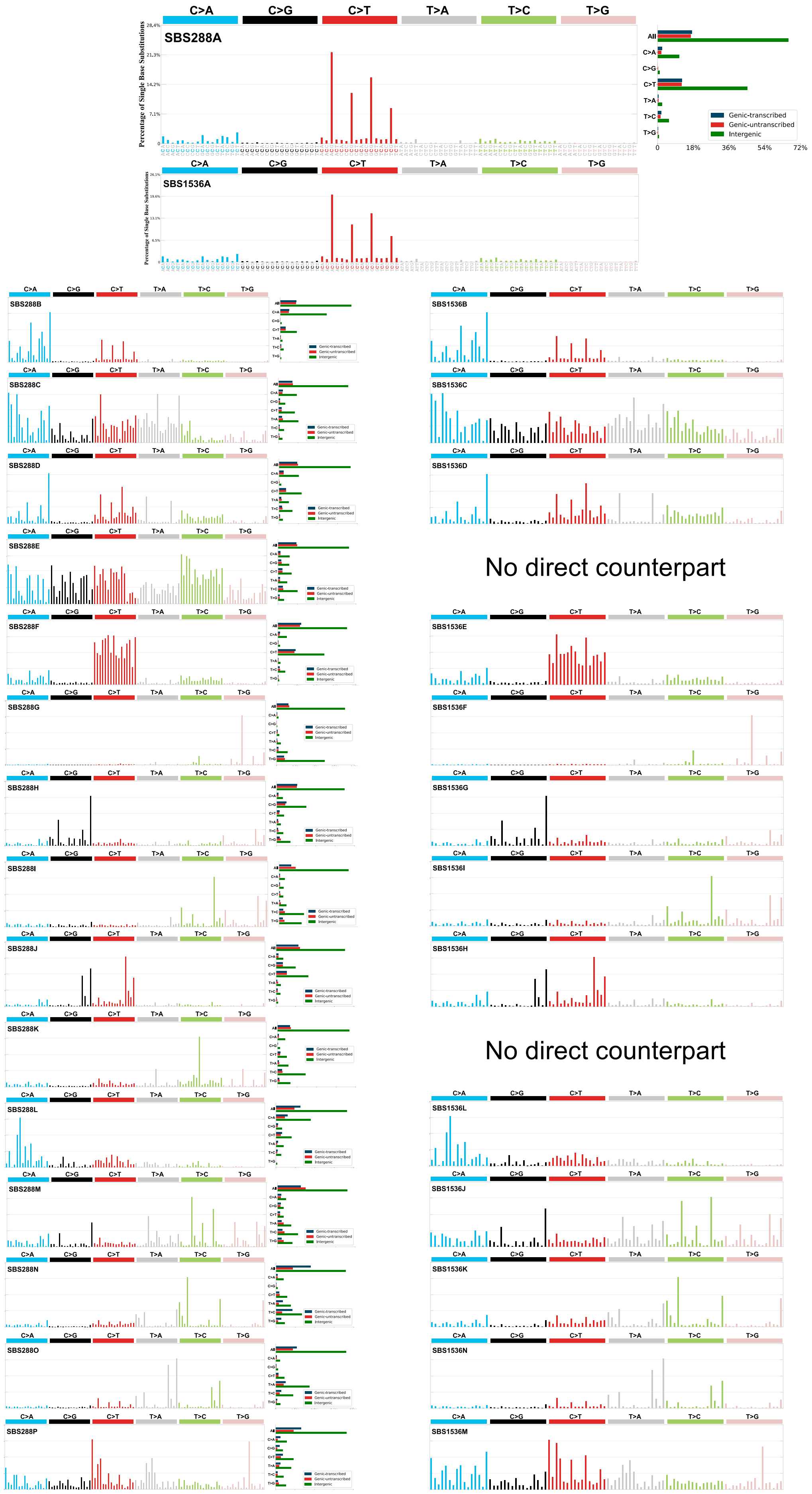

### Supplementary Figure 17

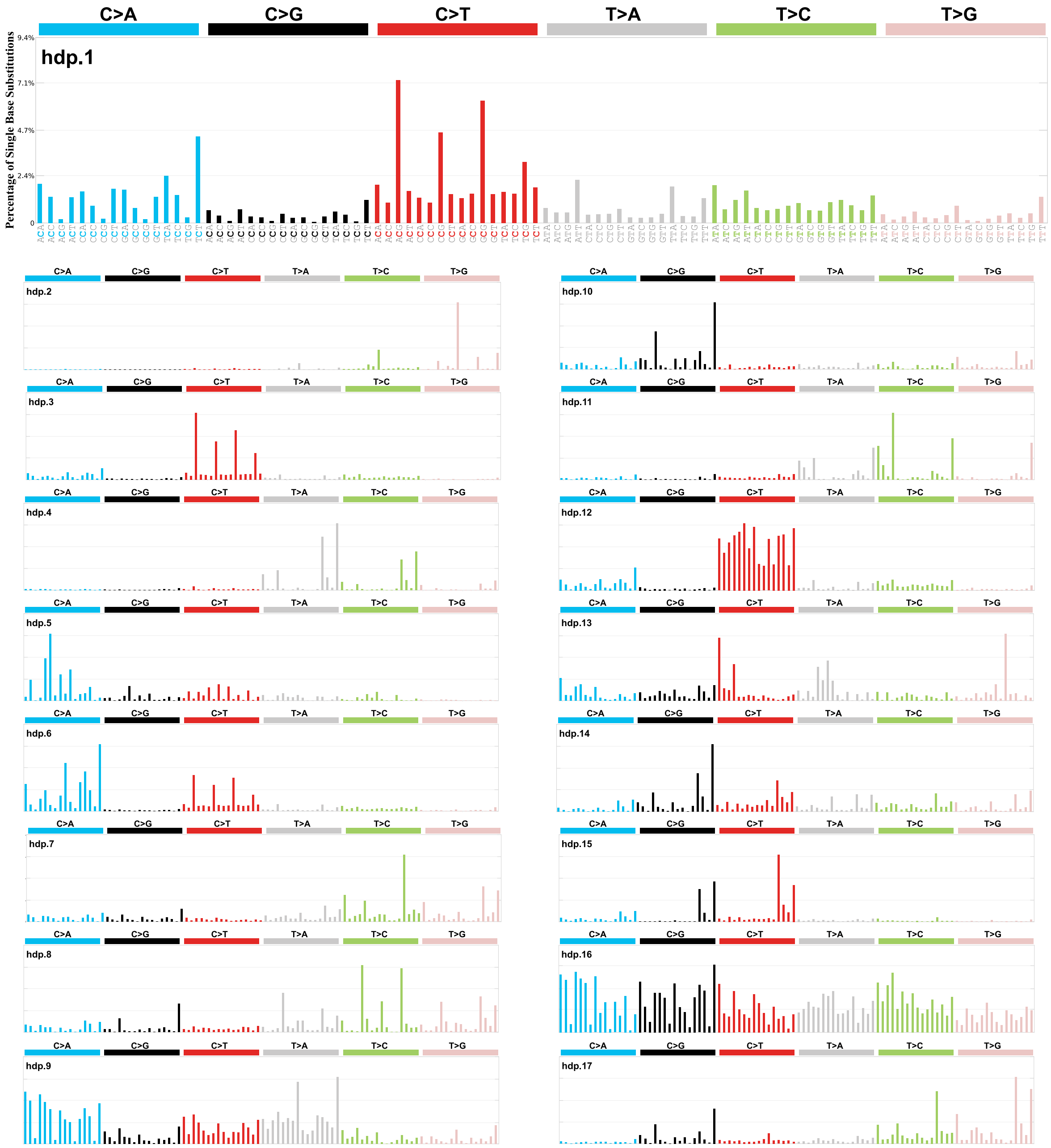

### Supplementary Figure 18

a

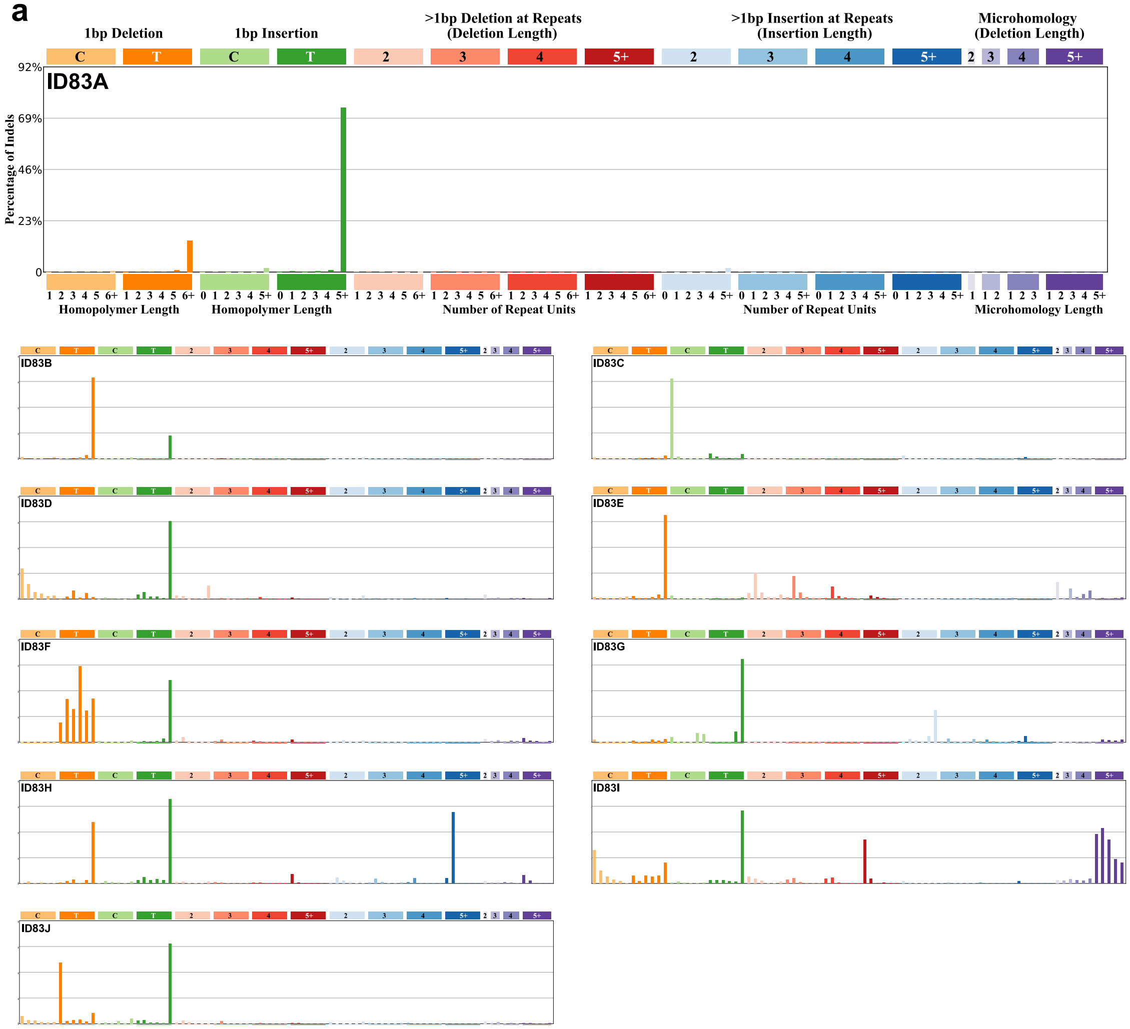


b

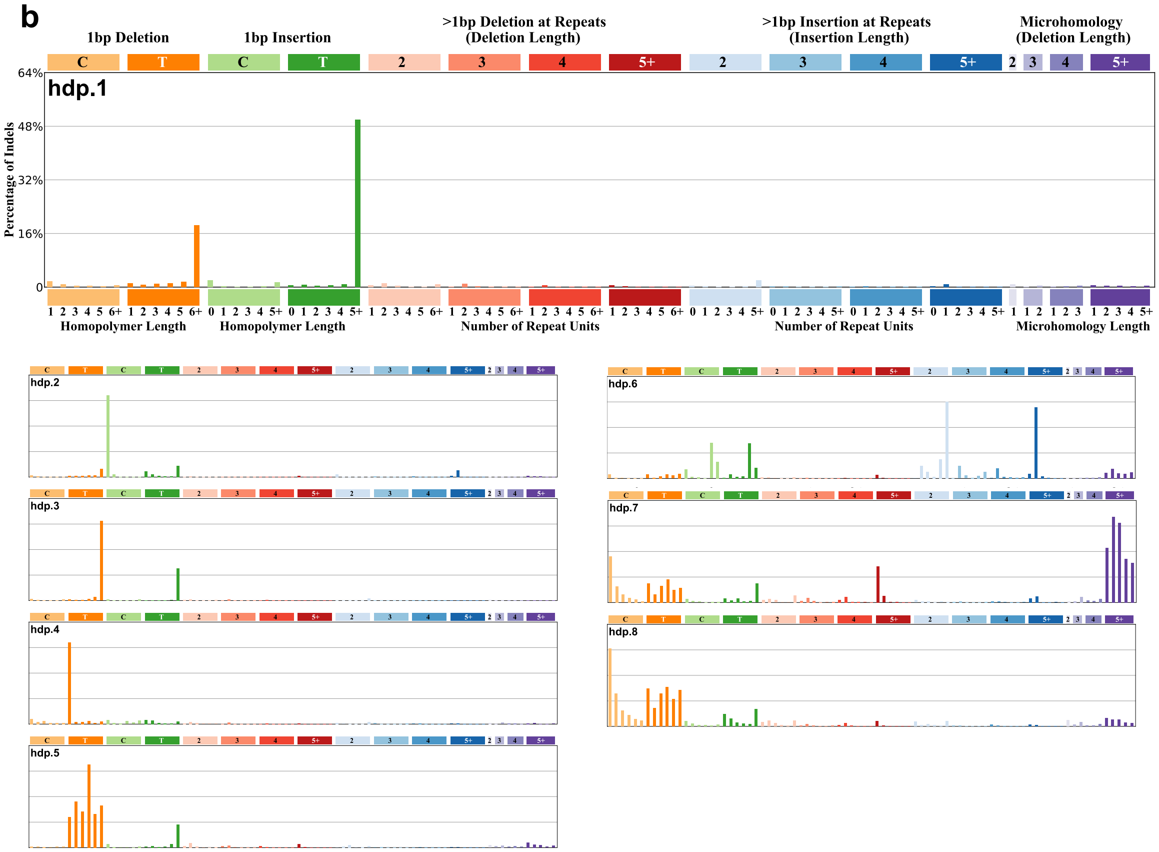

#### Supplementary Figure 19

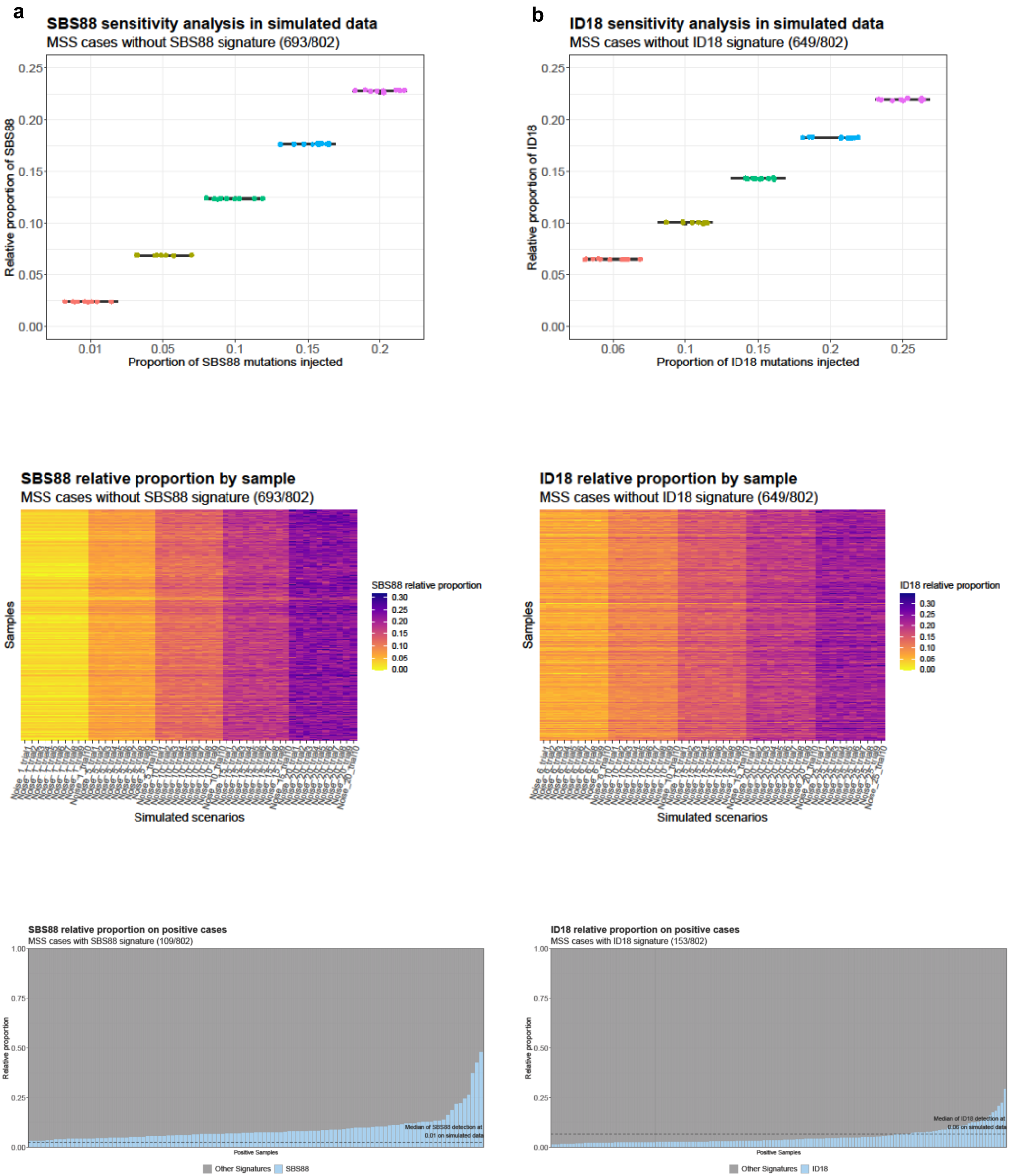
