## Supplementary Note for "Geographic and age-related variations in mutational processes in colorectal cancer"

**This file contains the following:**

Supplementary Results

Supplementary Methods

Supplementary Note Table Legends

Supplementary Note References

### SUPPLEMENTARY RESULTS

#### Identification of DNA repair-deficient cases

Microsatellite instability (MSI) was initially identified based on the observed mutational burdens and profiles (**Fig. 1*c***), as well as by identifying germline pathogenic mutations in the mismatch repair genes (*MLH1*, *MSH2*, *MSH6*, and *PMS2*) consistent with Lynch syndrome (**Supplementary Table 2**), classifying 153 colorectal cancers as MSI.

Apart from MSI, we also evaluated germline and somatic pathogenic mutations in specific genes (**Supplementary Table 3**), as well as the similarity of mutational profiles to previously known mutational signatures (**Supplementary Fig. 1-3**) to detect cases affected by other DNA repair deficiencies. This included the observation of specific mutations and signatures associated with defects in polymerase proofreading (*POLE* mutations and COSMICv3.4 SBS10a, SBS10b, and SBS28 signatures[1-3]; *POLD1* mutations and SBS10c[4]), homologous recombination repair (*BRCA1* and *BRCA2* mutations and SBS3 and ID6[5,6]); and base excision repair (*MUTYH* mutations and SBS36[7,8]; *NTHL1* mutations and SBS30[2,9]; *OGG1* mutations and Signal SBS108[10,11]). In addition, the Classifier of Homologous Recombination Deficiency (CHORD)[12] was run to refine the identification of homologous recombination repair deficient cases (**Supplementary Table 4**). In total, 10 cases were identified as harboring *POLE* mutations (with signatures associated with *POLE* mutations contributing on average 82.5% of mutations across all *POLE* mutated cases), 3 as harboring *POLD1* mutations (with *POLD1*-associated signatures contributing on average 61.1% of mutations), 7 as harboring homologous recombination deficiency (HRD; with HRD-associated signatures contributing on average 44.7% of indels), 2 as harboring *NTHL1* mutations (with *NTHL1*-associated signatures contributing on average 48.4% of mutations), 1 as harboring a

*MUTYH* mutation (with *MUTYH*-associated signatures contributing on average 83.2% of mutations), and 1 as harboring an *OGG1* mutation (with *OGG1*-associated signatures contributing on average 56.4% of mutations).

MSI tumors were enriched in the proximal colon (OR=12.2, $p=3.8\times10^{-27}$) and more common in early-onset patients (OR=2.6, $p=0.001$), although this latter association was not observed after excluding cases attributed to Lynch syndrome ($p>0.05$). Similar to MSI, homologous recombination repair deficient cases were also predominantly found in the proximal colon (OR=7.7, $p=0.009$), whereas no other DNA repair deficiency was significantly enriched early-onset or late-onset colorectal cancer patients ($p>0.05$).

**Validation of microsatellite instability status**

The MSI status for 131 of the 153 cases initially classified as MSI, where DNA was available post whole-genome sequencing (**Supplementary Note Table 1**), was independently validated using droplet digital PCR (ddPCR; **Methods**). Of these, 130 cases (99.2%) identified as MSI using mutational burdens and profiles were confirmed by ddPCR (**Supplementary Note Table 1**). Interestingly, the one case classified as microsatellite stable (MSS) by ddPCR showed loss of *PMS2* by immunohistochemistry. Although no germline or somatic mutations were found in known mismatch repair genes, this tumor displayed a mutational profile compatible with the presence of MSI (**Supplementary Fig. 12**) and was therefore labeled as MSI for subsequent analyses.

**Repertoire of mutational signatures in MSI colorectal cancers**

*De novo* extraction detected 15 single base substitution (SBS), 3 small insertion and deletion (ID), 6 double base substitution (DBS), 3 copy number (CN), and 2 structural variant (SV) mutational signatures in the 153 MSI colorectal tumors (**Supplementary Note Tables 2-12**). Upon decomposition to known COSMICv3.4 signatures, the MSI cohort predominantly displayed signatures associated with mismatch repair deficiencies. Specifically, the analysis identified all MSI-associated SBS, DBS, and CN signatures[1,3,5,13], including SBS6, SBS14, SBS15, SBS20, SBS21, SBS26, SBS44, DBS7, DBS10, and CN25 (**Supplementary Note Table 7**). Notably, three novel SBS signatures (SBS_I_MSI, SBS_N_MSI, and SBS_O_MSI) and one novel DBS signature (DBS_B_MSI) were also identified (**Supplementary Fig. 13*a-c***). Signature DBS_B_MSI was found in 114 of the 153 MSI colorectal tumors (74.5%), being the most prevalent DBS signature in 101 of these cases (**Supplementary Note Table 10**). In contrast, signatures SBS_I_MSI, SBS_N_MSI, and SBS_O_MSI were present in a small subset (<10% of all samples) of the MSI colorectal tumors, where they contributed many thousands of somatic mutations, being the most prevalent SBS signatures in a total of five cases (three cases for SBS_I_MSI, one for SBS_N_MSI, and one for SBS_O_MSI; **Supplementary Fig. 13*d***; **Supplementary Note Table 8**). SBS_M_MSI was not found in COSMICv3.4, but the signature was previously observed in a handful of breast, central nervous system, colorectal, and prostate tumors from Dutch and British patients, contributing a high amount of mutations and being associated with mismatch repair deficiency[10]. Similarly, in our cohort, SBS_M_MSI was only found present in 13 of the 153 MSI cases (8.5%), being the most prevalent signature in only one of them (**Supplementary Fig. 13*e***; **Supplementary Note Table 8**). Lastly, an MSI-associated artifactual signature DBS14, caused by the difficulty in aligning regions of long T or A repeats[10,13], was found in 7 of the 153 MSI colorectal cancers (4.6%).

In addition to MSI-associated signatures, our analysis identified other mutational signatures operative within the MSI cohort, including clock-like signatures SBS1, SBS5, ID1, and ID2[3,5], reactive oxygen species-linked signature SBS18[3], acetaldehyde-linked signature DBS2[5], ploidy-linked signatures CN1, CN2, and CN9[14], and homologous recombination deficiency associated signature SV3[2,13]. As previously reported[15], the number of mutations contributed by clock-like signatures was elevated in the MSI cases when compared to the MSS cases, with ID2 showing the highest elevation (median fold-change=273.7, $p$=$9.9\times10^{-86}$). Additionally, mutational signatures with unknown etiology were also detected within the MSI cohort, including SBS17a/b, SBS34, SBS93, DBS4, DBS8, DBS15, DBS16, DBS18, SV3, SV5, SV7, and SV_D, where SV_D was previously extracted in the MSS colorectal cancer cohort (**Methods**).

**Cancer driver genes in MSI colorectal cancers**

Consensus driver gene identification using the Intogen framework[16] identified a total of 31 cancer genes under positive selection in the MSI cases, 17 of them shared with the MSS cohort (**Supplementary Note Table 13**). Twenty-nine of these driver genes were previously reported in colorectal cancer[16,17] and two in other cancer types (*DDX3X* and *ACSL6*)[16]. We also identified *CDKN1B*, *TCF3*, and *TOP1* for the first time as driver genes for MSI colorectal cancer, after being reported recently as colorectal cancer drivers in UK patients exclusively for the MSS molecular subgroup[17].

### Geographic and age-related variations in mutational signatures and driver genes in MSI colorectal cancers

Despite exhibiting a significantly higher number of substitutions than MSS tumors (95,426 vs. 12,054; FC=7.92; $p=1.6\times10^{-85}$), MSI colorectal cancers display a lower diversity of mutational signatures due to the dominance of mismatch repair deficiency-associated signatures, which account for the majority of somatic mutations. Furthermore, limited geographic or age-related differences were observed in MSI cases, likely attributed to the smaller sample size and the predominance of mutations driven by defective DNA repair mechanisms. In particular, only mutational signature ID1 was found at significantly lower levels in the Czech Republic (OR=0.04, $q$=0.025), whereas no other statistically significant differences across countries were observed for either the prevalence of mutational signatures ($q$>0.05; **Supplementary Note Table 14**), or driver mutations affecting detected cancer driver genes ($q$>0.05; **Supplementary Note Table 15**), or hotspot driver mutations ($q$>0.05; **Supplementary Note Table 16**) across MSI cases, after adjusting by the effect of age, sex, tumor subsite, and tumor purity. Signature CN25, previously associated with mismatch repair deficiency[13], was found enriched in MSI patients older than 50 years (OR=20.0, $q$=0.012), while no other MSI signatures were found to be associated with the age of tumor onset ($q$>0.05; **Supplementary Note Table 17**). In addition, driver mutations in *APC* (OR=10.5, $q=3.0\times10^{-4}$) and *PIK3CA* (OR=8.1, $q$=0.004) were enriched in early-onset MSI patients, while *BRAF* (OR=25.5, $q$=0.002) and *RNF43* (OR=4.3, $q$=0.045) were enriched in late-onset MSI patients (**Supplementary Note Table 18**). In particular, the enrichment for *BRAF* mutations was driven by the p.V600E *BRAF* hotspot mutation (OR=21.9, $q$=0.005; **Supplementary Note Table 19**), which accounted for 55 out of the total 58 *BRAF* mutations, all of them present in late-onset MSI patients.

After excluding the 31 Lynch syndrome MSI cases, no additional associations were observed for mutational signatures, driver mutations, geographic locations, or ages of onset (**Supplementary Note Tables 20-25**; *q*>0.05), with ID1 burden still decreased in the Czech Republic (OR=0.03, *q*=0.020; **Supplementary Note Table 20**) and *APC* (OR=13.5, *q*=0.009; **Supplementary Note Table 24**) enriched in early-onset MSI cases, and *BRAF* (OR=20.1, *q*=0.029; **Supplementary Note Table 24**) and the p.V600E *BRAF* hotspot mutation (OR=17.9, *q*=0.042; **Supplementary Note Table 25**) enriched in late-onset MSI cases.

**Germline variants in MSS and MSI colorectal cancers**

To access the impact of pathogenic or likely pathogenic germline variants, we access the ClinVar status of germline variants in 292 colorectal cancer predisposition syndromes genes[18], homologous recombination genes (HR)[19], and DNA damage repair genes (DDR)[20]. We identified a total of 127 pathogenic or likely pathogenic germline variants in 41 of the genes analyzed **(Supplementary Note Table 28**). The MSS cases harbored 78 variants in 75 cases, including one variant in a gene related to Cowden syndrome (*SEC23B*), two variants on mismatch repair genes related to Lynch syndrome (*MLH1* and *MSH6*), 52 variants on DDR-genes, 23 on HR-genes. None of these variants showed enrichment in early-onset MSS cases (**Supplementary Fig. 14*a***; **Supplementary Note Table 29**). In the MSI cases, in addition to the 31 variants in DNA mismatch repair genes associated with Lynch syndrome, we also identified 10 pathogenic or likely pathogenic variants on DDR-genes, *BRCA1*, and *SEC23B* (**Supplementary Note Table 28**). Cases harboring pathogenic or likely pathogenic variants in DNA mismatch repair genes consistent with Lynch syndrome showed enrichment in early-onset patients (OR=4.8, *q*=0.010; **Supplementary Fig. 14*b***; **Supplementary Note Table 30**).

**Microbiome analysis of signatures enriched in early-onset MSS colorectal cancers**

We selected genera with a minimum abundance of 1,000 reads across all samples and filtered out those with low overall variance in the dataset. This process resulted in 91 genera for association analysis with mutational signatures. To ensure an unbiased analysis, we stratified the mutational signatures into binary classes and applied a binomial logistic regression model. For each mutational signature, we conducted a separate regression model for each of the 91 genera. We included the genus as a covariate alongside all other covariates previously included in our regression models (age of diagnosis, sex, tumor subsite, country, and tumor purity). In addition, we also included as a covariate the total number of reads, as a proxy for sequencing depth, considering its potential impact on the microbiome analysis. After applying false discovery rate (FDR) p-value correction, we did not identify any significant association between specific genera and SBS89, SBS_M, or ID14.

As an alternative analysis, to address potential multicollinearity among the genera and reduce the number of genus-specific models, we also performed a hierarchical clustering using the Bray-Curtis distance metric. This approach grouped the 91 genera identified in the initial analysis into 50 clusters, using a distance threshold of 0.70 (**Supplementary Fig. 15**). For each mutational signature, we conducted a separate regression model for each of the 50 clusters, replacing the individual genera used previously. Each model included the cluster as a covariate alongside the same base-level covariates specified previously. By incorporating clusters instead of individual genera, we reduced the dimensionality of the data and accounted for correlated features. This clustering-based approach allowed us to test associations between microbial community structures and mutational signatures. However, even with this adjustment, we did not find significant associations for SBS89, SBS_M, or ID14. Further exploration of microbial data in future studies, potentially integrating additional cases harboring these signatures, more detailed metadata, or functional microbial pathways, could provide insights into the underlying causes of SBS89, SBS_M, and ID14, and their effect on early colorectal mutagenesis.

### SUPPLEMENTARY METHODS

#### Extraction of *de novo* mutational signatures with SigProfilerExtractor in microsatellite stable tumors

Single base substitutions (SBS) extractions were performed using SigProfilerExtractor[21] with both SBS-288 and SBS-1536 contexts. SBS-288 extends the SBS-96 contexts by classifying mutations into transcribed, untranscribed, or intergenic non-transcribed regions, whereas SBS-1536 considers the two flanking bases on either side of the mutated base to form a pentanucleotide context[22]. These extractions extracted 16 and 14 signatures, respectively (**Supplementary Fig. 16)**. In order to calculate the cosine similarity, the SBS-288 and SBS-1536 signatures were both collapsed into the SBS-96 mutational context. 14 signatures were extracted in both formats with cosine similarity >0.9 (**Supplementary Fig. 16**; **Supplementary Note Table 26**). The two signatures extracted only in the SBS-288 format were SBS_E and SBS_K. The former is a flat signature that decomposed to SBS1, SBS3 and SBS5, and the latter represents an incomplete separation of SBS17a and SBS17b (**Supplementary Table 10**). The SBS-288 results were used for the final analysis as this format allowed the extraction of these additional signatures. Notably, all four signatures where decomposition was rejected (SBS_F, SBS_H, SBS_M, and SBS_O) could be reproduced in both SBS-288 and SBS-1536 formats with a cosine similarity >0.95 (**Supplementary Note Table 26**).

#### Extraction of *de novo* mutational signatures with mSigHdp in microsatellite stable tumors

In order to validate the mutational signatures obtained using SigProfilerExtractor, extractions were also performed with a second algorithm, mSigHdp[23]. SBS extraction was performed using the SBS-96 mutational context only, as mSigHdp has not been benchmarked for extended contexts. In total, mSigHdp extracted 17 signatures (**Supplementary Fig. 17**), of which 15 were very close matches (cosine similarity >0.90) to the SBS-288 results, whereas signatures hdp.14 and hdp.17 were unique to mSigHdp (**Supplementary Note Table 26**). The first unique signature, hdp.14, was an additional APOBEC containing signature, whereas hdp.17 was a combination of the new signature hdp.10 (SBS_H) and SBS93 (hdp.7 / SBS_I). The latter was confirmed by performing a decomposition using the panel of COSMICv3.4 and the new colorectal cancer signatures as the signature database. Again, all four signatures where decomposition was rejected (SBS_F, SBS_H, SBS_M, and SBS_O) can be reproduced in mSigHdp with a cosine similarity >0.95 (**Supplementary Note Table 26**), indicating that these signatures are highly reproducible using independent methodology.

mSigHdp extracted 8 indel signatures in comparison to the 10 extracted from SigProfilerExtractor (**Supplementary Fig. 18**). Of these, 6 mSigHdp signatures could be matched directly to those from SigProfilerExtractor (**Supplementary Note Table 27**). Of the remaining 2 mSigHdp signatures, hdp.6 appears to be a combination of the SigProfilerExtractor *de novo* signatures ID_G and ID_H (but missing the ID1 component of both of those *de novo* signatures), whereas hdp.8 is a combination of ID2, ID5, and ID9. ID5 is not included in the final panel of signatures decomposed from the SigProfilerExtractor (**Supplementary Table 10**; **Extended Data Fig. 4*d-e***) and was not included in the final panel of signatures used for signature assignment (**Supplementary Table 12**). Notably, the novel signature ID_J (**Extended Data Fig. 4*e***) was reproducible in the mSigHdp but with low cosine similarity (0.64; **Supplementary Note Table 27**). However, the low cosine similarity is explained by the lack of ID1 contamination in the signature extracted using mSigHdp.

**Decomposition to reference signatures**

The extracted *de novo* mutational signatures were decomposed into the COSMICv3.4 reference signatures using SigProfilerAssignment[24]. As the COSMICv3.4 reference signatures are only available for the SBS-96 mutational context, the *de novo* SBS-288 signatures were collapsed into the SBS-96 context. As a result of the loss of information from extended contexts and also due to the large number of COSMICv3.4 reference mutational signatures, it is likely that the decompositions on default settings will include signatures that are implausible given the cancer type.

For the MSS signatures using the default decomposition settings, only SBS_M was not decomposed. In order to optimize the decomposition, the following signature subgroups were excluded (using the *exclude_signature_subgroups* parameter of SigProfilerAssignment): artifact signatures, ultraviolet signatures, lymphoid signatures, mismatch repair deficiency signatures, polymerase deficiency signatures, base excision repair deficiency signatures, and treatment signatures. In addition, the *new_signature_threshold* was set to 0.90. Using these settings, SBS_H, SBS_O, and ID_J, in addition to SBS_M were not decomposed. SBS_F was still decomposed into SBS1, SBS5, SBS19 and SBS40a in the optimized decomposition. However, this was rejected on the basis that SBS_F had been previously extracted in an independent cohort[10] and the lack of individual spectra that supported the presence of SBS19 (the other reference signatures all appeared in other decompositions). In contrast, SBS_D, despite being a borderline signature, with a cosine similarity of 0.90, had not previously been extracted in independent cohorts. Therefore, we chose to be conservative and not consider SBS_D as a novel signature. As such, SBS_D was decomposed into SBS1, SBS5, SBS18, and SBS34 (**Supplementary Table 10**).

For the MSI extractions, using default settings for SigProfilerAssignment, only SBS_M_MSI and DBS_B_MSI were not decomposed. In order to optimize the decomposition, the following signature subgroups were excluded (using the *exclude_signature_subgroups* parameter): artifact signatures, ultraviolet signatures, lymphoid signatures, polymerase deficiency signatures, base excision repair deficiency signatures, homologous repair deficiency signatures, and treatment signatures. Optimizing did not change the number of signatures that were not decomposed. However, the decompositions for SBS_I_MSI, SBS_N_MSI, and SBS_O_MSI were subsequently rejected on the basis that individual spectra existed that strongly support these signatures being the result of distinct mutational processes (**Supplementary Fig. 13*d***).

**Quantification of DNA repair deficiency-associated mutational signatures in DNA repair deficient cases**

To quantify the number of mutations contributed by signatures associated with DNA repair deficiencies in the 24 cases classified as DNA repair deficient, we assigned directly all SBS and ID COSMICv3.4 signatures[25], as well as Signal SBS108[10] (related to *OGG1* deficiency) to these samples using SigProfilerAssignment[24]. The following SBS signatures were considered to quantify the mutations contributed by *MUTYH* mutations (COSMIC SBS36; **Supplementary Fig. 2*b***), *NTHL1* mutations (COSMIC SBS30; **Supplementary Fig. 2*d***), *OGG1* mutations (Signal SBS108; **Supplementary Fig. 2*f***), *POLD1* mutations (COSMIC SBS10c; **Supplementary Fig. 2*h***), and *POLE* mutations (COSMIC SBS10a, SBS10b, SBS28; **Supplementary Fig. 3*b***), whereas

COSMIC ID signature ID6 was considered to characterize the indels contributed by homologous recombination deficiency (**Supplementary Fig. 1*b***).

**Sensitivity analysis for the detection of colibactin signatures in microsatellite stable tumors**

To access the resolution for detecting colibactin signatures in MSS cases, we performed simulations for both SBSs and IDs. Specifically, synthetic SBS88 and ID18 mutations were injected at different average levels in each sample (scenarios 1%, 5%, 10%, 15% and 20% for SBS88 and 6%, 10%, 15%, 20% and 25% for ID18) for all MSS cases where the two colibactin-associated signatures were not originally detected. Mutations were injected according to a Gaussian distribution where the mean was equal to a percentage of a sample's total mutational burden, and the standard deviation was equal to 10% of the mean. Importantly, the overall mutational burden for each sample was kept the same by randomly subtracting the same number of mutations that were injected into the sample, while ensuring all mutation counts were still non-negative. Mutational signatures were re-extracted as done for the original data, and MSA attributions were performed using the same penalties applied for the original data. In the SBS context, our analysis indicates that among the non-SBS88 positive cases in the original data (693/802), SBS88 was attributed to samples if it contributes at least 2.5% of mutations (median of 0.025 relative proportion at the level of 1% injection; **Supplementary Fig. 19*a***). Indeed, MSA attributed SBS88 in about 90.6% of simulation trials at the 1% injection level, approximately 99.7% at the 5% injection level, and 100% at the 10%, 15%, and 20% injection levels. For the ID context, the results indicate that among the non-ID18 positive cases in the original data (649/802), ID18 was attributed to samples if it contributed at least 6.6% of mutations (**Supplementary Fig. 19*b***). MSA attributed ID18 in about 99.8% of simulation trials at the 6% injection level, and 100%

at 10%, 15%, 20%, and 25% injection levels. These results suggest that our analyses are unlikely to have overlooked SBS88 and ID18 in the examined set of MSS colorectal cancers, assuming they contribute at least 1% and 6% of mutations per sample, respectively.

**SUPPLEMENTARY NOTE TABLES**

**Supplementary Note Table 1. Validation of MSI status using droplet digital PCR.**

**Supplementary Note Table 2. Mutational profiles of de novo SBS signatures extracted in MSI colorectal cancer cases.**

**Supplementary Note Table 3. Mutational profiles of de novo ID signatures extracted in MSI colorectal cancer cases.**

**Supplementary Note Table 4. Mutational profiles of de novo DBS signatures extracted in MSI colorectal cancer cases.**

**Supplementary Note Table 5. Mutational profiles of de novo CN signatures extracted in MSI colorectal cancer cases.**

**Supplementary Note Table 6. Mutational profiles of de novo SV signatures extracted in MSI colorectal cancer cases.**

**Supplementary Note Table 7. Decomposition of de novo MSI colorectal cancer signatures into previously reported signatures.**

**Supplementary Note Table 8. Sample attributions of decomposed SBS signatures in MSI colorectal cancer cases.**

**Supplementary Note Table 9. Sample attributions of decomposed ID signatures in MSI colorectal cancer cases.**

**Supplementary Note Table 10. Sample attributions of decomposed DBS signatures in MSI colorectal cancer cases.**

**Supplementary Note Table 11. Sample attributions of decomposed CN signatures in MSI colorectal cancer cases.**

**Supplementary Note Table 12. Sample attributions of decomposed SV signatures in MSI colorectal cancer cases.**

**Supplementary Note Table 13. Driver genes detected in MSI colorectal cancer cases.**

**Supplementary Note Table 14. Enrichment of mutational signature prevalence in specific countries compared to all others in MSI colorectal cancer cases.**

**Supplementary Note Table 15. Enrichment of driver mutations in cancer driver genes in specific countries compared to all others in MSI colorectal cancer cases.**

**Supplementary Note Table 16. Enrichment of hotspot driver mutations in specific countries compared to all others in MSI colorectal cancer cases.**

**Supplementary Note Table 17. Enrichment of mutational signature prevalence in late-onset compared to early-onset MSI colorectal cancer cases.**

**Supplementary Note Table 18. Enrichment of driver mutations in cancer driver genes in late-onset compared to early-onset MSI colorectal cancer cases.**

**Supplementary Note Table 19. Enrichment of hotspot driver mutations in late-onset compared to early-onset MSI colorectal cancer cases.**

**Supplementary Note Table 20. Enrichment of mutational signature prevalence in specific countries compared to all others in MSI colorectal cancer cases, excluding Lynch syndrome cases.**

**Supplementary Note Table 21. Enrichment of driver mutations in cancer driver genes in specific countries compared to all others in MSI colorectal cancer cases, excluding Lynch syndrome cases.**

**Supplementary Note Table 22. Enrichment of hotspot driver mutations in specific countries compared to all others in MSI colorectal cancer cases, excluding Lynch syndrome cases.**

**Supplementary Note Table 23. Enrichment of mutational signature prevalence in late-onset compared to early-onset MSI colorectal cancer cases, excluding Lynch syndrome cases.**

**Supplementary Note Table 24. Enrichment of driver mutations in cancer driver genes in late-onset compared to early-onset MSI colorectal cancer cases, excluding Lynch syndrome cases.**

**Supplementary Note Table 25. Enrichment of hotspot driver mutations in late-onset compared to early-onset MSI colorectal cancer cases, excluding Lynch syndrome cases.**

**Supplementary Note Table 26. Comparison of single base substitution signatures extracted by SigProfilerExtractor and mSigHdp in MSS colorectal cancer cases.**

**Supplementary Note Table 27. Comparison of indel signatures extracted by SigProfilerExtractor and mSigHdp in MSS colorectal cancer cases.**

**Supplementary Note Table 28. Pathogenic variants in genes associated with colorectal cancer syndromes, homologous recombination, and DNA damage repair.**

**Supplementary Note Table 29. Enrichment of pathogenic or likely pathogenic germline variants in late-onset compared to early-onset MSS colorectal cancer cases.**

**Supplementary Note Table 30. Enrichment of pathogenic or likely pathogenic germline variants in late-onset compared to early-onset MSI colorectal cancer cases.**

### SUPPLEMENTARY NOTE REFERENCES


1 Alexandrov, L. B. *et al.* Clock-like mutational processes in human somatic cells. *Nat Genet* **47**, 1402-1407 (2015). https://doi.org/10.1038/ng.3441

2 Nik-Zainal, S. *et al.* Landscape of somatic mutations in 560 breast cancer whole-genome sequences. *Nature* **534**, 47-54 (2016). https://doi.org/10.1038/nature17676

3 Alexandrov, L. B. *et al.* Signatures of mutational processes in human cancer. *Nature* **500**, 415-421 (2013). https://doi.org/10.1038/nature12477

4 Robinson, P. S. *et al.* Increased somatic mutation burdens in normal human cells due to defective DNA polymerases. *Nature Genetics* **53**, 1434-1442 (2021). https://doi.org/10.1038/s41588-021-00930-y

5 Alexandrov, L. B. *et al.* The repertoire of mutational signatures in human cancer. *Nature* **578**, 94-101 (2020). https://doi.org/10.1038/s41586-020-1943-3

6 Nik-Zainal, S. *et al.* Mutational processes molding the genomes of 21 breast cancers. *Cell* **149**, 979-993 (2012). https://doi.org/10.1016/j.cell.2012.04.024

7 Pilati, C. *et al.* Mutational signature analysis identifies MUTYH deficiency in colorectal cancers and adrenocortical carcinomas. *J Pathol* **242**, 10-15 (2017). https://doi.org/10.1002/path.4880

8 Viel, A. *et al.* A Specific Mutational Signature Associated with DNA 8-Oxoguanine Persistence in MUTYH-defective Colorectal Cancer. *EBioMedicine* **20**, 39-49 (2017). https://doi.org/10.1016/j.ebiom.2017.04.022

9 Grolleman, J. E. *et al.* Mutational Signature Analysis Reveals NTHL1 Deficiency to Cause a Multi-tumor Phenotype. *Cancer Cell* **35**, 256-266 e255 (2019). https://doi.org/10.1016/j.ccell.2018.12.011

10 Degasperi, A. *et al.* Substitution mutational signatures in whole-genome-sequenced cancers in the UK population. *Science* **376**, abl9283 (2022). https://doi.org/10.1126/science.abl9283

11 Zou, X. *et al.* A systematic CRISPR screen defines mutational mechanisms underpinning signatures caused by replication errors and endogenous DNA damage. *Nature Cancer* **2**, 643-657 (2021). https://doi.org/10.1038/s43018-021-00200-0

12 Nguyen, L., W. M. Martens, J., Van Hoeck, A. & Cuppen, E. Pan-cancer landscape of homologous recombination deficiency. *Nature Communications* **11**, 5584 (2020). https://doi.org/10.1038/s41467-020-19406-4

13 Everall, A. *et al.* Comprehensive repertoire of the chromosomal alteration and mutational signatures across 16 cancer types from 10,983 cancer patients. *medRxiv*, 2023.2006.2007.23290970 (2023). https://doi.org/10.1101/2023.06.07.23290970

14 Steele, C. D. *et al.* Signatures of copy number alterations in human cancer. *Nature* **606**, 984-991 (2022). https://doi.org/10.1038/s41586-022-04738-6

15 Díaz-Gay, M. & Alexandrov, L. B. in *Advances in Cancer Research* Vol. 151 (eds Franklin G. Berger & C. Richard Boland) 385-424 (Academic Press, 2021).

16 Martinez-Jimenez, F. *et al.* A compendium of mutational cancer driver genes. *Nat Rev Cancer* **20**, 555-572 (2020). https://doi.org/10.1038/s41568-020-0290-x

17 Cornish, A. J. *et al.* The genomic landscape of 2,023 colorectal cancers. *Nature* (2024). https://doi.org/10.1038/s41586-024-07747-9

18 Board, W. C. o. T. E. *WHO classification of Tumours: Digestive System Tumours*. 5th edn, (World Health Organization, 2019).

19 Riaz, N. *et al.* Pan-cancer analysis of bi-allelic alterations in homologous recombination DNA repair genes. *Nature Communications* **8**, 857 (2017). https://doi.org/10.1038/s41467-017-00921-w

20 Knijnenburg, T. A. *et al.* Genomic and Molecular Landscape of DNA Damage Repair Deficiency across The Cancer Genome Atlas. *Cell Reports* **23**, 239-254.e236 (2018). https://doi.org/10.1016/j.celrep.2018.03.076

21 Islam, S. M. A. *et al.* Uncovering novel mutational signatures by de novo extraction with SigProfilerExtractor. *Cell Genom* **2**, 100179 (2022). https://doi.org/10.1016/j.xgen.2022.100179

22 Bergstrom, E. N. *et al.* SigProfilerMatrixGenerator: a tool for visualizing and exploring patterns of small mutational events. *BMC Genomics* **20**, 685 (2019). https://doi.org/10.1186/s12864-019-6041-2

23 Liu, M., Wu, Y., Jiang, N., Boot, A. & Rozen, S. G. mSigHdp: hierarchical Dirichlet process mixture modeling for mutational signature discovery. *NAR Genom Bioinform* **5**, lqad005 (2023). https://doi.org/10.1093/nargab/lqad005

24 Díaz-Gay, M. *et al.* Assigning mutational signatures to individual samples and individual somatic mutations with SigProfilerAssignment. *Bioinformatics* **39**, btad756 (2023). https://doi.org/10.1093/bioinformatics/btad756

25 Sondka, Z. *et al.* COSMIC: a curated database of somatic variants and clinical data for cancer. *Nucleic Acids Res* **52**, D1210-D1217 (2024). https://doi.org/10.1093/nar/gkad986
